# Pre-diagnostic plasma metabolomics and the risk of exfoliation glaucoma

**DOI:** 10.1101/2021.08.02.21261514

**Authors:** Jae H. Kang, Oana Zeleznik, Lisa Frueh, Jessica Lasky-Su, A. Heather Eliassen, Clary Clish, Bernard A. Rosner, Louis R. Pasquale, Janey L. Wiggs

**Author notes:** Correspondence and reprints: Jae Hee Kang, ScD, Channing Division of Network Medicine, 181 Longwood Avenue, Boston, MA 02114. contributed equally (Jae H. Kang, Oana Zeleznik).

## Abstract

**Objective:** To identify pre-diagnostic plasma metabolomic biomarkers associated with risk of exfoliation glaucoma (XFG).

**Methods:** We conducted a metabolomic study using a 1:1 matched nested case-control study design within the Nurses’ Health Study (NHS) and Health Professionals Follow-up Study (HPFS). Participants provided blood samples in 1989-’90 (NHS) and 1993-’95 (HPFS); we identified 205 participants who newly developed XFG during follow-up to 2018 (average time to diagnosis from blood draw =11.8 years); XFG was confirmed with medical record review. We profiled plasma metabolites using liquid chromatography-mass spectrometry and identified 379 known metabolites that passed quality control checks. Metabolites were transformed using probit scores for normality. We used multivariable-adjusted logistic regression adjusting for matching factors (such as age, residential latitude, season and time of blood draw), glaucoma family history and other covariates. Metabolite Set Enrichment Analysis was used to identify metabolite classes associated with risk of XFG. Number of effective tests (NEF) and False Discovery Rate (FDR) were used to adjust for multiple comparisons.

**Results:** Mean age of cases (n=205) at diagnosis was 71 years; 84% were women and >99% were Caucasian; matched controls (n=205) all reported eye exams as of the matched cases’ index date. A total of 33 metabolites were nominally significantly associated with XFG risk (p<0.05) and 4 metabolite classes were significantly associated (FDR<0.05). Overall, adverse associations were observed for the classes of lysophosphatidylcholines (FDR=0.02) and phosphatidylethanolamine plasmalogens (FDR=0.004). Inverse associations were observed for triglycerides (FDR<0.001) and steroid and steroid derivatives (FDR=0.03); in particular, the multivariable-adjusted odds ratio for XFG risk associated with each 1 standard deviation increase in plasma cortisone levels was 0.49 (95% CI=0.32-0.74; NEF=0.05). Results did not differ materially by time between blood draw and diagnosis, latitude of residence (< or ≥41^°^N latitude), age (< or ≥60 years), sex or glaucoma family history.

**Conclusions:** Four broad classes of metabolites (including steroids such as cortisone and 3 lipid classes) in pre-diagnostic plasma collected almost a decade before diagnosis were associated with XFG risk; these results should be confirmed in future studies.

## INTRODUCTION

Exfoliation syndrome (XFS) is a systemic age-related disorder of the extracellular matrix that primarily manifests as accumulation of white or grey flaky deposits in the ocular anterior segment.^1^ XFS is the most common identifiable cause of glaucoma (XFG), an irreversible blinding disease, and this systemic disorder has been associated with greater risks of cataract and several systemic diseases.^2,3^

XFS pathogenesis may involve a stress-induced elastosis characterized by excessive production and abnormal cross-linking of microfibrillar components into fibrillar aggregates (XFM: exfoliation material) in the ocular anterior segment and connective tissues in other organs. ^1,4^ XFM contains glycoproteins, proteoglycans, and several elastic fiber components and the cross-linking enzyme, lysyl oxidase-like 1 (LOXL1).^5^ Genome-wide association studies have identified several genetic associations with XFS, including *LOXL1*, a principal genetic risk factor for XFS.^6^ While *LOXL1* variants are very strongly associated with XFS, the same variants are also prevalent in unaffected populations, making this a complex disease involving genetic factors, environmental factors and their interactions .^7^

Given that XFS etiology is poorly understood, we conducted an untargeted metabolomics study to identify potential metabolomic markers for XFS. Metabolomics, which is the profiling of small molecules (metabolites) in the body, is a powerful tool for biomarker discovery. To date, there have been only two other metabolomic studies in XFS/XFG;^8,9^ however, these studies were both relatively small (<35 XFS cases) and cross-sectional in design. In this study, we included 205 cases and 205 matched controls in a nested case-control study of pre-diagnostic circulating plasma metabolites from ∼10 years before XFS/XFG diagnosis.

## METHODS

### Study Population

We conducted a nested case-control study in the Nurses’ Health Study (NHS) and Health Professional Follow-up Study (HPFS). In 1976, the NHS began with 121,700 female nurses in the US aged 30–55 years, and in 1986, the HPFS was initiated with 51,529 male US health professionals aged 40–75 years. Participants have responded to biennial questionnaires that updated information on lifestyle and medical conditions, such as glaucoma. Blood samples were collected in 1989–’90 among 32,826 NHS participants and in 1993–’95 among 18,159 HPFS participants. In both cohorts, each participant arranged to have a blood sample drawn and shipped by overnight courier to the laboratory. Samples were processed and aliquots of white blood cell, red blood cell, and plasma were archived in liquid nitrogen freezers (≤-130°C).

XFG cases were diagnosed after blood draw until June 1, 2016 (NHS), or January 1, 2016 (HPFS). Cases of XFG (205 total; 174 women and 31 men) were first identified by self-report of glaucoma and then confirmed by medical record review. Controls were 1:1 matched to cases on: age, sex, ancestry (Scandinavian, Southern-European Caucasian, other Caucasian, others), month and year of blood collection, time of day of blood draw, fasting status (>8 hours or ≤8 hours), latitude and longitude of residence as of blood draw, and among women: additional matching on menopausal status and hormone therapy use at blood draw (premenopausal, postmenopausal and hormone therapy use, postmenopausal and no hormone therapy use, missing/ unknown) as well as at diagnosis. For matching factors and additional covariate data adjusted for in statistical models, we used questionnaire data collected as of the blood draw, and if not available, we used biennial questionnaire data prior to the blood sample. The study protocol was approved by the institutional review board of the Brigham and Women’s Hospital and Harvard T.H. Chan School of Public Health. The return of the self-administered questionnaire and blood sample was considered as implied consent.

### Metabolite profiling

Plasma metabolites were profiled using liquid chromatography tandem mass spectrometry (LC-MS) to measure endogenous, polar metabolites and lipids as described previously^10-12^ (Broad Institute of MIT and Harvard University (Cambridge, MA)). Of the 398 unique known metabolites evaluated, we excluded 19 metabolites that did not pass quality control checks (10 that were adversely impacted by delay in blood processing inherent in the archived study samples and 9 metabolites that had coefficients of variation (CVs) >25%),^13^ leaving 379 metabolites for analyses. For metabolites with <10% missing across participant samples, missing values were imputed with 1/2 of the minimum value measured for that metabolite. All metabolites included in the analysis exhibited good within person reproducibility over a 1- year period, indicating that one blood sample provides a reasonable measure of longer-term exposure. The 379 metabolites were assigned a metabolite class based on chemical taxonomy; we evaluated 17 metabolite classes: steroids and steroid derivatives; carnitines; diglycerides; triglycerides; cholesteryl esters; lysophosphatidylethanolamines (LPEs); phosphatidylethanolamines (PEs); lysophosphatidylcholines (LPCs); phosphatidylcholines (PCs); phosphatidylethanolamine plasmalogens (PEPs); phosphatidylcholine plasmalogens (PCPs); organoheterocyclic compounds; ceramides; carboxylic acids and derivatives; organic acids and derivatives; nucleosides, nucleotides, and analogues; sphingomyelins. Metabolite values were transformed to probit scores to scale the metabolite levels to the same range and minimize the influence of skewed distributions on the results.

For the metabolite of homocysteine, which has been previously associated with XFS/XFG, ^14-16^ we determined the concentration of total homocysteine at the Boston Children’s Hospital using an enzymatic assay on the Roche P Modular system (Roche Diagnostics - Indianapolis, IN), using reagents and calibrators from Catch Inc. (Seattle, WA), as homocysteine levels assessed using the metabolomics platform described above did not pass quality control checks. The CVs for both NHS and HPFS samples were <10%.

### Statistical analysis

We used conditional logistic regression in Model 1 to evaluate metabolite associations with risk of XFG. For both the individual metabolite level analyses as well as metabolite class analyses, in successive models, we used nested multivariable-adjusted conditional logistic regression adjusting for the matching factors as well as additional important covariates. In Model 2, we adjusted for factors as of blood draw that have a major influence on metabolites: age (years; matching factor), sex, month of blood draw (matching factor), year of blood draw (matching factor), time of blood draw (matching factor), fasting status (>8 hours or less), body mass index (kg/m^2^), smoking status (never, past, current), physical activity (metabolic equivalents of task / week). In Model 3, we expanded Model 2 by adding established risk factors for XFG: glaucoma family history, ancestry (Scandinavian ancestry, other Caucasian, others), time spent outdoors in the summer during youth,^17^ history of non-melanoma skin cancer,^18^ latitude of residence^19^ and residential population density.^20^ In Model 4, we added additional XFG risk factors, namely the intakes of dietary folate,^21^ caffeine,^22^ alcohol and total caloric intake. In Model 5, we further added co-morbidities that have been associated with XFG previously: stroke, heart disease, hypertension, high cholesterol, diabetes, hearing loss and sleep duration. ^23-25^ Finally, in Model 6, we added use of oral or inhaled steroid use, to minimize confounding by a commonly used drug that has been associated with glaucoma.

For analyses of individual metabolites, metabolite values were used as continuous variables (per 1 standard deviation increase) to calculate linear trend p-values. We estimated the odds ratios (OR) and 95% confidence intervals (95% CI) per 1 standard deviation (SD) increase in metabolite levels. For analyses of metabolite classes, Metabolite Set Enrichment Analysis was used to identify metabolite classes associated with risk of XFG. For the evaluation of individual metabolites, “number of effective tests” (NEF)^26^ was used to adjust for multiple comparisons given the high correlation structure of the metabolomics data; NEF<0.05 was considered statistically significant while metabolites with NEF<0.2 were considered as candidates worthy of further study given the exploratory nature of the analyses. For the evaluation of metabolite classes, False Discovery Rate (FDR) ^27^ was used; FDR<0.05 was considered statistically significant. All analyses were performed with SAS 9.4 and R 3.4.1.

## RESULTS

### Study population

Of the 205 cases, 84.9% were female, with a mean age (SD) at blood draw of 59.3 (SD=6.5) years and at diagnosis of 71.1 (SD=7.4) years. Mean follow-up time was 11.8 years. Controls were similar to cases in matching factors. Distributions of XFG risk factors were generally in the expected directions for cases and controls (**Table 1**).

**Table 1.**
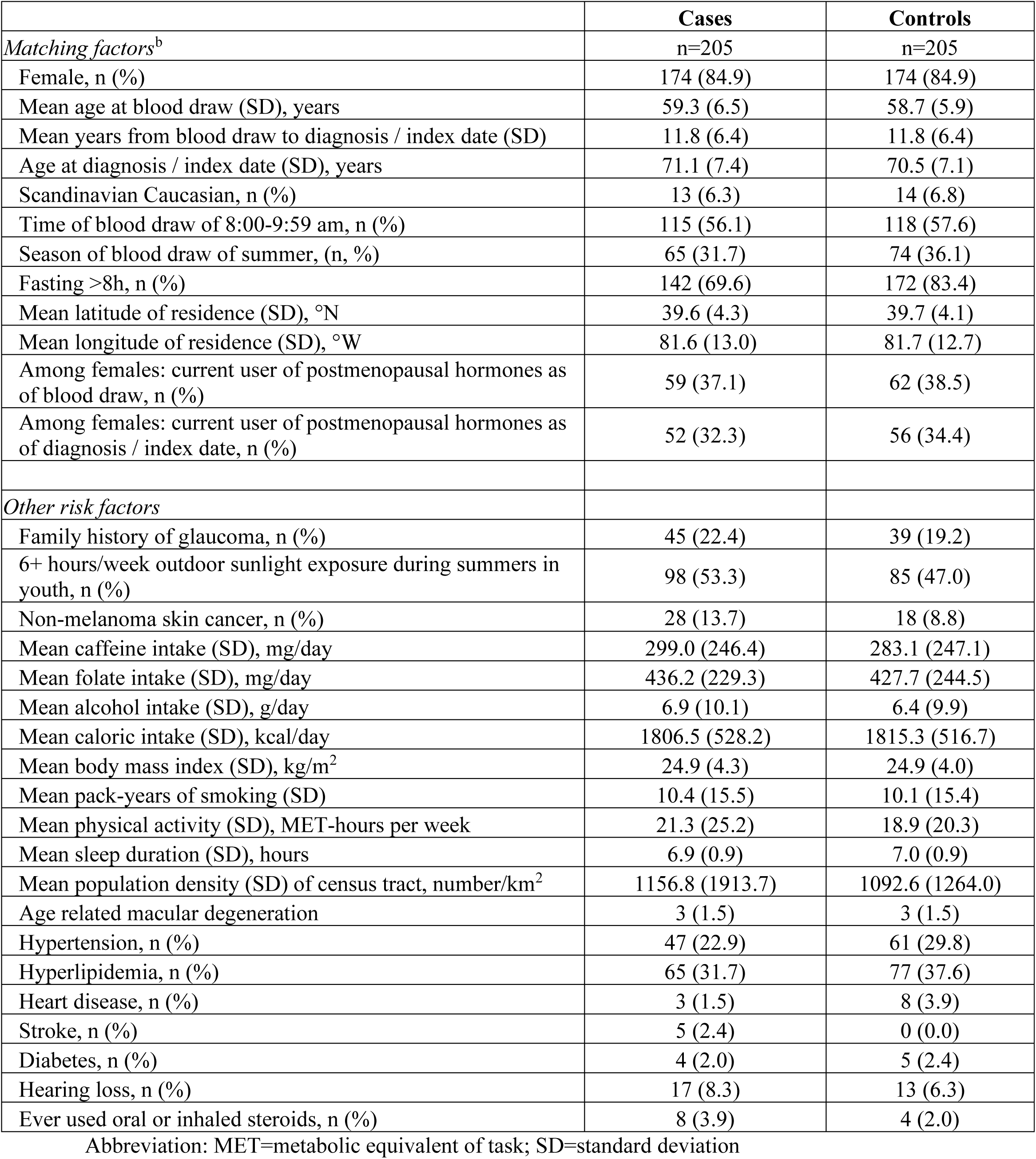
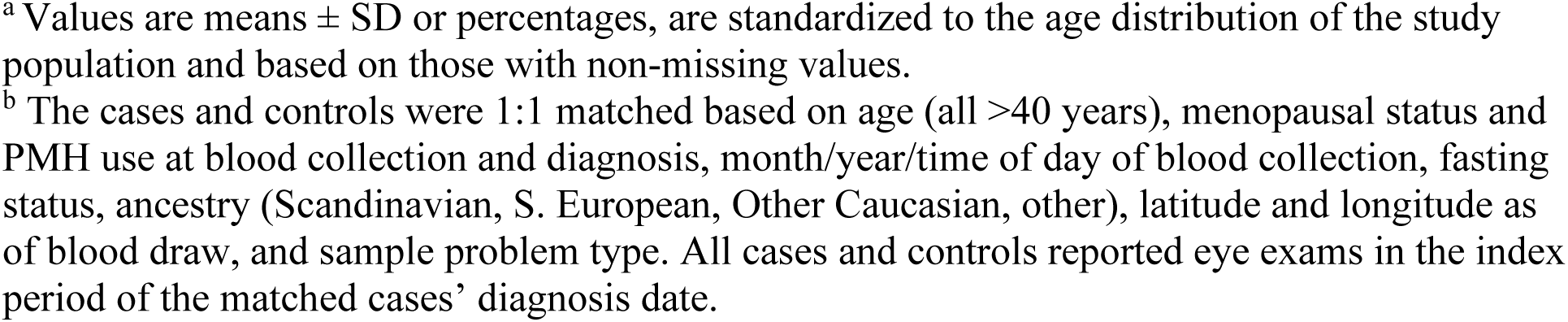
Age and age-standardized characteristics of exfoliation glaucoma cases and their matched controls ^a^

|  | **Cases** | **Controls** |
| --- | --- | --- |
| *Matching factors<sup>b</sup>* | n=205 | n=205 |
| Female, n (%) | 174 (84.9) | 174 (84.9) |
| Mean age at blood draw (SD), years | 59.3 (6.5) | 58.7 (5.9) |
| Mean years from blood draw to diagnosis / index date (SD) | 11.8 (6.4) | 11.8 (6.4) |
| Age at diagnosis / index date (SD), years | 71.1 (7.4) | 70.5 (7.1) |
| Scandinavian Caucasian, n (%) | 13 (6.3) | 14 (6.8) |
| Time of blood draw of 8:00-9:59 am, n (%) | 115 (56.1) | 118 (57.6) |
| Season of blood draw of summer, (n, %) | 65 (31.7) | 74 (36.1) |
| Fasting >8h, n (%) | 142 (69.6) | 172 (83.4) |
| Mean latitude of residence (SD), °N | 39.6 (4.3) | 39.7 (4.1) |
| Mean longitude of residence (SD), °W | 81.6 (13.0) | 81.7 (12.7) |
| Among females: current user of postmenopausal hormones as of blood draw, n (%) | 59 (37.1) | 62 (38.5) |
| Among females: current user of postmenopausal hormones as of diagnosis / index date, n (%) | 52 (32.3) | 56 (34.4) |
| *Other risk factors* |  |  |
| Family history of glaucoma, n (%) | 45 (22.4) | 39 (19.2) |
| 6+ hours/week outdoor sunlight exposure during summers in youth, n (%) | 98 (53.3) | 85 (47.0) |
| Non-melanoma skin cancer, n (%) | 28 (13.7) | 18 (8.8) |
| Mean caffeine intake (SD), mg/day | 299.0 (246.4) | 283.1 (247.1) |
| Mean folate intake (SD), mg/day | 436.2 (229.3) | 427.7 (244.5) |
| Mean alcohol intake (SD), g/day | 6.9 (10.1) | 6.4 (9.9) |
| Mean caloric intake (SD), kcal/day | 1806.5 (528.2) | 1815.3 (516.7) |
| Mean body mass index (SD), kg/m<sup>2</sup> | 24.9 (4.3) | 24.9 (4.0) |
| Mean pack-years of smoking (SD) | 10.4 (15.5) | 10.1 (15.4) |
| Mean physical activity (SD), MET-hours per week | 21.3 (25.2) | 18.9 (20.3) |
| Mean sleep duration (SD), hours | 6.9 (0.9) | 7.0 (0.9) |
| Mean population density (SD) of census tract, number/km<sup>2</sup> | 1156.8 (1913.7) | 1092.6 (1264.0) |
| Age related macular degeneration | 3 (1.5) | 3 (1.5) |
| Hypertension, n (%) | 47 (22.9) | 61 (29.8) |
| Hyperlipidemia, n (%) | 65 (31.7) | 77 (37.6) |
| Heart disease, n (%) | 3 (1.5) | 8 (3.9) |
| Stroke, n (%) | 5 (2.4) | 0 (0.0) |
| Diabetes, n (%) | 4 (2.0) | 5 (2.4) |
| Hearing loss, n (%) | 17 (8.3) | 13 (6.3) |
| Ever used oral or inhaled steroids, n (%) | 8 (3.9) | 4 (2.0) |

Abbreviation: MET=metabolic equivalent of task; SD=standard deviation
<sup>a</sup> Values are means ± SD or percentages, are standardized to the age distribution of the study population and based on those with non-missing values.
<sup>b</sup> The cases and controls were 1:1 matched based on age (all >40 years), menopausal status and PMH use at blood collection and diagnosis, month/year/time of day of blood collection, fasting status, ancestry (Scandinavian, S. European, Other Caucasian, other), latitude and longitude as of blood draw, and sample problem type. All cases and controls reported eye exams in the index period of the matched cases' diagnosis date.

### Relation between individual metabolites and XFG

**Supplementary eTable 1** presents the association with homocysteine (per 1 SD increase) and XFG; no significant associations were observed across the various models (p>0.18; Model 6 OR=1.22 (95%CI=0.90, 1.65)). **Supplementary eTable 2** presents the associations between all of the 379 individual metabolites and XFG, and **Figure 1** shows the main results of the individual metabolite analyses; 33 metabolites among 379 metabolites that were at least nominally significant at p<0.05 in any of the models (Model 1 through Model 6) were plotted. In the results from Model 1 (crude model adjusting for matching factors only), 20 metabolites were nominally significant, including acetaminophen, methionine sulfoxide, C16:0 LPE, which were all adversely associated (NEF<0.2); in Model 2, where we more finely adjusted for matching factors (e.g., linear age, fasting status, time of day and season of blood draw and other major determinants of variability in metabolites such as BMI), only 9 metabolites were nominally significant. However, in Model 3 and Model 4, with the addition of XFG established and suspected risk factors related to greater time spent outdoors and factors that may raise serum homocysteine levels as covariates, we observed that cortisone emerged as an NEF significant metabolite that was strongly inversely associated with XFG risk: Model 4 results for each 1SD difference in cortisone level was OR= 0.51 (95%CI=0.36- 0.73; NEF=0.01). Also, in Model 4, gabapentin, an anticonvulsant and nerve pain medication, was adversely associated with XFG at OR=1.90 (95%CI=1.26-2.87; NEF=0.16). With Model 5 to further adjust for co-morbidities and Model 6 with the additional adjustment for the use of oral/inhaled corticosteroids, we observed that the effect estimates were similar for cortisone (OR=0.49 (95%CI=0.32-0.74; NEF=0.05)) and gabapentin (OR=2.21 (95% CI=1.31-3.73; NEF=0.21)).

**Figure 1.**
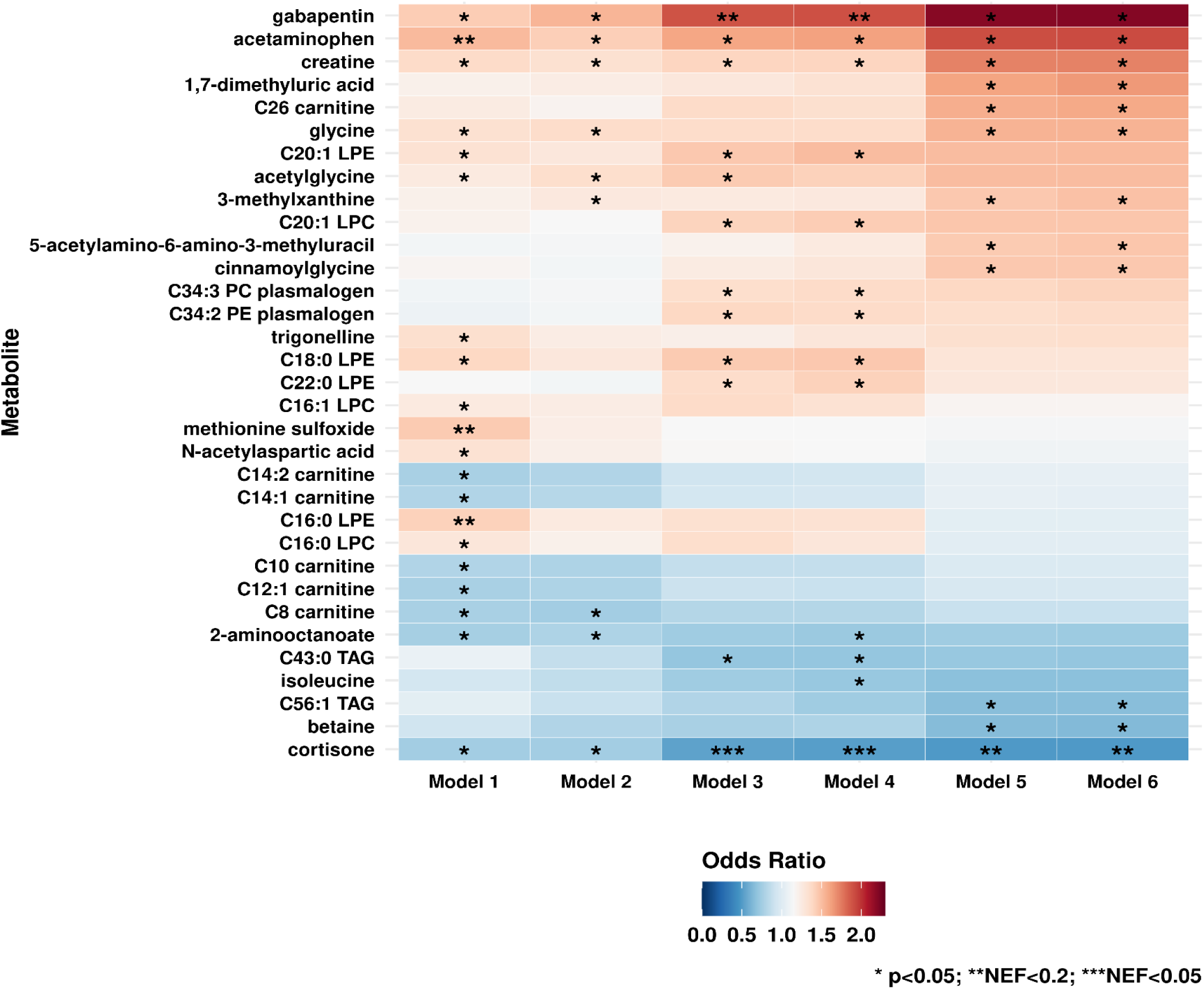
Individual metabolites among the n=379 metabolites evaluated that were significant across the various nested multiple conditional logistic regression models of exfoliation glaucoma (205 cases and 205 controls). **Model 1**: basic model, adjusting for matching factors only (see Table 1); **Model 2** (factors that affect metabolite levels): Model 1 + age, sex, smoking status, BMI, physical activity, time of day of blood draw, month of blood draw, fasting status ; **Model 3** (presumed exfoliation syndrome risk factors): Model 2 + type of Caucasian, family history of glaucoma, time spent outdoors in sunlight in the summer in youth, non-melanoma skin cancer, latitude, population density; **Model 4** (factors that may raise homocysteine levels): Model 3 + folate intake, caffeine intake, alcohol intake, caloric intake; **Model 5** (systemic comorbidities suggested to be associated with XFS in some studies): Model 4 + heart disease, stroke, diabetes, hypertension, high cholesterol, hearing loss, sleep duration; **Model 6** (use of drugs associated with glaucoma): Model 5 + steroid use. * p<0.05; * * Number of effective tests (NEF)<0.2; * * * NEF<0.05

### Relation between metabolite classes and XFG

Figure 2 shows the results of evaluating metabolite classes and XFG; results for all 17 metabolite classes are plotted. Model 1 and Model 2 showed generally similar results with FDR-significant positive associations for LPEs and in Model 2, LPCs, nucleosides, nucleotides and analogues newly emerged as significantly adversely associated metabolite classes while carnitines, and triglycerides were significantly inversely associated. In Model 3 and Model 4, with the addition of XFG risk factors, the adverse associations with LPE and LPCs and inverse associations with triglycerides were robust; cholesteryl esters, PEPs and PCPs newly emerged as NEF significantly adversely associated while steroids and steroid derivatives were NEF significantly inversely associated. In Model 5 and Model 6 to additionally adjust for co-morbidities and oral/inhaled corticosteroids, associations with cholesteryl esters, LPEs and PCPs were no longer significant. In the final model, Model 6, we observed 4 classes that were NEF significant: steroids and steroid derivatives and triglycerides were inversely associated (both NEF=0.02), while various lipid classes of LPCs (NEF= 0.03), and PEPs (NEF= 0.03) were positively associated.

**Figure 2.**
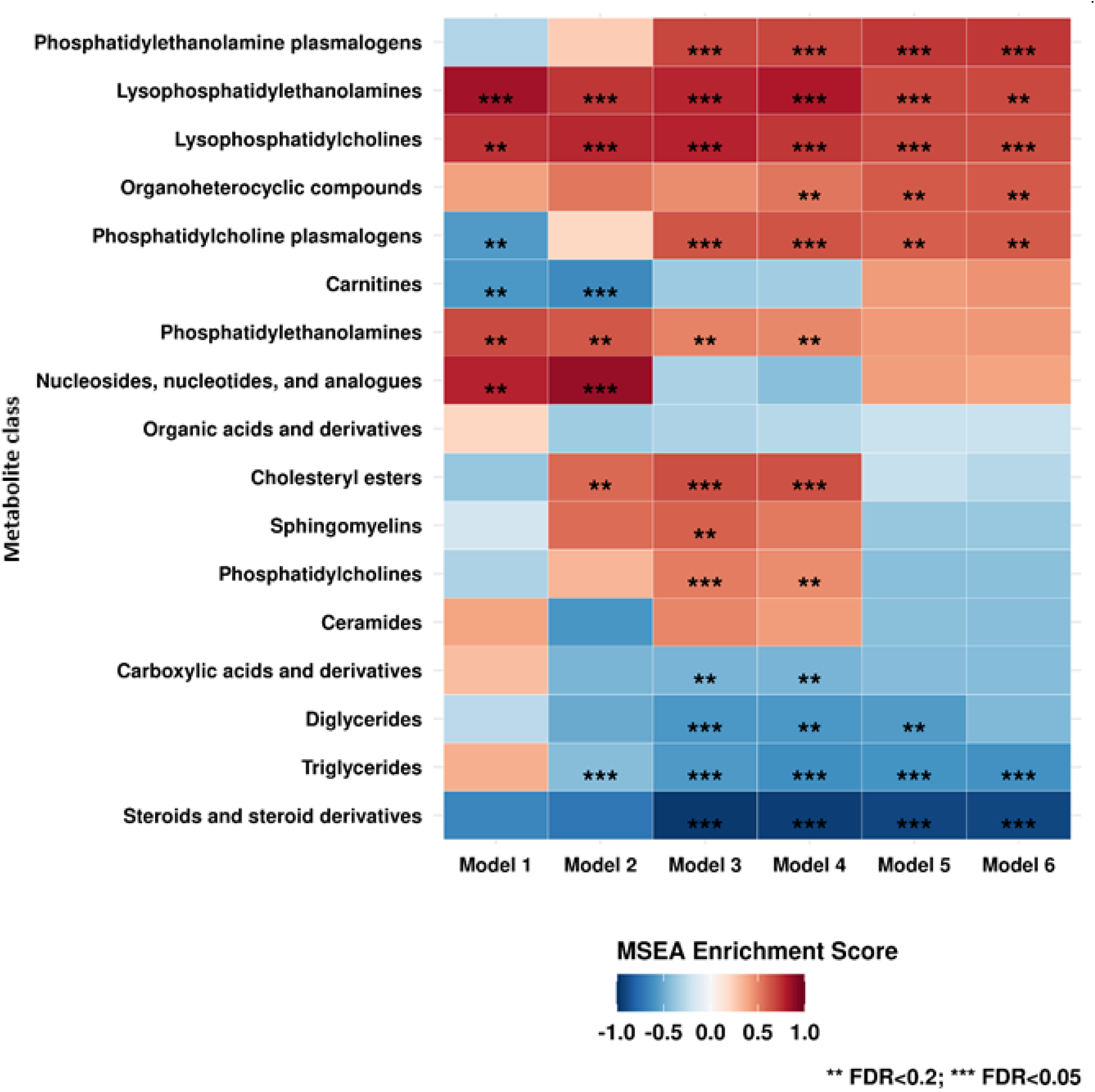
Metabolite classes (n=17) evaluated in various nested multiple conditional logistic regression models of exfoliation glaucoma (205 cases and 205 controls). **Model 1**: basic model, adjusting for matching factors only (see Table 1); **Model 2** (factors that affect metabolite levels): Model 1 + age, sex, smoking status, BMI, physical activity, time of day of blood draw, month of blood draw, fasting status; **Model 3** (presumed exfoliation syndrome risk factors): Model 2 + type of Caucasian, family history of glaucoma, time spent outdoors in sunlight in the summer in youth, non-melanoma skin cancer, latitude, population density; **Model 4** (factors that may raise homocysteine levels): Model 3 + folate intake, caffeine intake, alcohol intake, caloric intake; **Model 5** (systemic comorbidities suggested to be associated with XFS in some studies): Model 4 + heart disease, stroke, diabetes, hypertension, high cholesterol, hearing loss, sleep duration; **Model 6** (use of drugs associated with glaucoma): Model 5 + steroid use. * * False Discovery Rate (FDR)<0.2; * * * FDR<0.05.

### Secondary analyses

To evaluate whether associations differed by subgroups, we assessed whether results varied by age (**eFigure 1**), sex (**eFigure 2**), latitude (**eFigure 3**), time to diagnosis (**eFigure 4**), and history of glaucoma (**eFigure 5**). Results, particularly for cortisone, did not differ by subgroups; however, there was a trend for a stronger inverse association among women (which may be due to the much larger sample size among women) and those living in <41°N in latitude for cortisone. Gabapentin tended to be more strongly associated among those with a family history of glaucoma (OR=3.68 (95%CI=1.50-8.99)) than in those without (OR=1.08 (95%CI=0.85-1.39); **eFigure 5**). In sensitivity analyses excluding those who were diagnosed within 5 years of diagnosis in an attempt to remove those who already may have subclinical disease (**eFigure 6**), we observed that the association with cortisone was similar in magnitude (although it was no longer NEF significant). In sensitivity analyses, 1) we excluded those with another age-related ocular condition, age-related macular degeneration, or those who have ever reported using oral / inhaled corticosteroids (**eFigure 7a**) and 2) we further excluded those with conditions that would indicate treatment with corticosteroids (**eFigure 7b**); we observed that the inverse association results for cortisone were robust (OR=0.50 (95%CI=0.32-0.78) and OR=0.55 (95%CI=0.28-1.08), respectively), indicating that the lower cortisone levels in XFG cases than controls was not explained by corticosteroid use.

## DISCUSSION

In this nested case-control study of pre-diagnostic plasma metabolites in relation to XFG (n=410), with a mean 11.8 years between blood draw and diagnosis, we observed that the metabolite cortisone and the metabolite class of triglycerides was inversely associated with XFG while the lipid classes of LPCs, and PEPs were adversely associated with XFG. Because this study was the first to evaluate the relation between pre-diagnostic plasma metabolites, these findings should be confirmed in future studies.

Two previous studies have investigated the metabolic signature of XFS. Leruez, et al. ^8^ found that compared to controls with cataracts (n=18), XFS cases (n=16) showed elevated levels of amino acids and two carnitines (C8 and C10) but lower levels of neuroprotective spermine and spermidine polyamines, diacyl PC phospholipids (C38:0 and C38:1) and sphingomyelin (C26:1). Myer et al., evaluated metabolites in samples of aqueous humor from those with XFS (n=31; 18 women and 13 men; 29 with cataracts) versus controls (n=25; all men with cataracts) and observed that while creatine and 5-hydroxypentanoate were positively associated, propylene glycol, N(6)-acetonyllysine, and 5 amino acids were inversely associated with XFS. Our study results are not directly comparable in that prior studies were cross-sectional studies and samples were collected after diagnosis in XFS cases versus mainly those with cataracts as controls; in contrast, our larger study evaluated pre-diagnostic plasma and those with XFG were matched to those without XFG (regardless of cataract status). Therefore, our results are notable in addressing a major gap by providing data from pre-diagnostic plasma that may reflect metabolic changes occurring early in the XFG disease process.

### Cortisone and cortisol

The strongly inverse association between steroids and steroid derivatives, especially cortisone, and XFG was a notable finding. Relatively low levels of cortisone may reflect lower systemic endogenous anti-inflammatory corticosteroid levels that may prevent the disruption of the uveal tract blood-ocular barrier integrity leading to the subsequent formation of XFM. ^28^ Although absolute cortisone levels are unknown, relatively lower levels may also reflect adrenal insufficiency; this may be plausible given that body aches and joint pain are common symptoms,^29^ and gabapentin and acetaminophen were metabolites that were nominally significantly associated with XFG. Adrenal insufficiency has multiple causes;^29^ one cause may be the adrenal suppression brought on by exogenous corticosteroid use.^30^ Indeed, XFG cases had somewhat higher prevalence of corticosteroid use than controls (3.9% versus 2.0%), although the overall prevalence was low. To address this possibility, we conducted sensitivity analyses where we excluded participants who self-reported ever having taken oral or inhaled corticosteroids or had conditions that are treated with them, and we observed similar inverse associations. Furthermore, those with corticosteroid-induced secondary glaucoma were not included in our cases, making it unlikely that differences in corticosteroid use may be underlying all of the inverse associations with cortisone. The link between adrenal insufficiency and XFG is unclear; however, some speculative reasons for why cortisone may be inversely associated is that some studies^31-33^ though not all,^31,34^ have observed that low cortisone/cortisol may be a marker of greater UV exposure, which has been strongly implicated in the pathogenesis of XFS/XFG.^35^ Indeed, inverse associations with cortisone appeared to be stronger in the subgroup residing in ≤41° N latitude, where cortisone levels may be a stronger marker of greater UV exposure. Finally, cortisone/cortisol are glucocorticoids, and higher levels in controls versus cases may indicate that controls may have had more frequent hyperglycemia, and several studies^36-43^ have reported inverse associations between diabetes and XFG. The mechanism underlying this trend is unknown; however, it has been hypothesized that greater glycation of basement membrane components may prevent XFM aggregation. ^39,44^ This association warrants further study in relation to XFG.

### Lysophosphatidylcholines (LPCs)

LPCs are pro-inflammatory and may be involved with glaucoma via LPCs’ role in the autotaxin-lysophophatidic acid pathway in intraocular pressure regulation, although the data has been inconsistent.^8,45-49^ Our results are consistent with other studies of targeted lipidomics that have revealed higher total LPCs in the aqueous humor of eyes with glaucoma, including XFG;^45,46^ however, two POAG metabolomics studies of the aqueous humor observed null associations with LPCs,^47,48^ and Leruez et al.^8^ and another study of LPCs in the optic nerve tissue observed inverse associations.^49^ These discrepancies may be related to LPCs with inflammatory properties being limited to those with shorter and unsaturated fatty acids^50,51^ while LPCs with longer and more saturated fatty acids have been inversely associated with type 2 diabetes^52^ and heart disease.^53^ In our study, we had stronger associations with longer chain length (C20 and C22) LPCs; thus, it is also possible that higher levels of these long-chain LPCs may be markers of a healthier cardiometabolic profile in cases than controls and thus less frequent hyperglycemia that may be possibly associated with less XFM aggregation.^39,44^

### Phosphatidylethanolamine plasmalogens (PEPs)

Although higher PEPs have been associated with lower risk of dementia and cardiovascular disease via antioxidant and neuroprotective mechanisms, higher PEPs have also been observed with greater physical activity,^54^ which may be a marker for greater UV exposure that in turn would increase XFG risk. Also, higher PEPs have been associated with a lower quality diet, ^55-58^ characterized by low whole grains and vegetable intake; such a diet with fewer folate sources would be associated with higher homocysteine levels, which has been frequently observed in XFS.^15,16,59^ Although we observed that modest adverse associations with homocysteine itself was not significantly associated, given that other metabolites such as lower phosphatidylcholines, ^60^ lower betaine^61^ and higher levels of coffee related metabolites (i.e., 3-methylxanthine and trigonelline)^62,63^ that increase homocysteine were near significant, the dysregulation in the homocysteine pathway may still be important early on in the disease process.

### Triglycerides

Higher triglycerides as a metabolite class were significantly inversely associated with XFG, which is in contrast to findings from multiple cross-sectional studies that observed positive associations with XFG. ^64-68^ Although we adjusted for caffeine intake and recreational physical activity, lower triglycerides in cases may be capturing residual information about greater coffee consumption^69^ or lower sedentary behavior and greater physical activity, ^70,71^ which are likely associated with higher homocysteine levels and greater UV exposure,^72^ respectively, and higher XFG risk. Also, as higher triglyceride levels are a component of metabolic syndrome, it could also be a proxy for greater hyperglycemia, which may have protective effects against the accumulation of XFM. ^39,44^

A limitation of our data is that our study population was relatively homogenous, with mostly White health professionals; therefore, our findings may not be generalizable to other populations with different race and ethnicity composition. Also, there may have been residual confounding by other unmeasured factors in these results. Our blood samples were collected at one timepoint; however, we have only included metabolites with good correlations for within person stability over at least 1 year. Our study had several strengths. To our knowledge, this is the first study assessing the associations of pre-diagnostic metabolites and XFG risk. Our study has a relatively large sample size with 205 cases and 205 matched controls. Additional strengths include the detailed covariate information and long follow-up time (mean=11.8 years).

Overall, these results provide new and promising candidates for risk detection biomarkers. Additional population studies evaluating metabolites in pre-diagnostic blood are needed to validate these findings. If confirmed, these markers may be used to identify high-risk participants who can benefit from more frequent eye exams.

## Data Availability

Information including the procedures to obtain and access data from the Nurses' Health Studies and Health Professionals Follow-up Study is described at https://www.nurseshealthstudy.org/researchers (contact) and https://sites.sph.harvard.edu/hpfs/for-collaborators/

## ACKNOWLEDGMENTS

This work was supported by NEI R01 EY020928, R01 EY015473 (LRP), NCI UM1 CA186107, U01 CA167552, and an unrestricted challenge grant to Icahn School of Medicine at Mount Sinai, Department of Ophthalmology from Research to Prevent Blindness (LRP). The content is solely the responsibility of the authors and does not necessarily represent the official views of the National Institutes of Health.

