## Supplemental material for "Pre-diagnostic plasma metabolomics and the risk of exfoliation glaucoma"

### Table of Contents

eFigure 1. Secondary analysis by age (< vs. ≥60 years; n=214 vs. n=196). 3

eFigure 2. Secondary analysis by sex (Nurses Health Study (NHS; n=348) participants are all women and Health Professionals Follow-up Study (HPFS; n=62) participants are all men). 4

eFigure 3. Secondary analysis by latitude (<41°N in latitude vs. ≥41°N in latitude; n=182 vs. n=228). 5

eFigure 4. Secondary analysis by time to diagnosis (<11.75 vs. ≥11.75 years; n=204 vs. 206). 6

eFigure 5. Secondary analysis by self-reported glaucoma family history (yes vs. no; n=84 vs n=320). 7

eFigure 6. Sensitivity analyses excluding those who were diagnosed within 5 years after blood draw (n=336). 8

eFigure 7a. Sensitivity analyses excluding those who had a history of age-related macular degeneration or history of use of oral / inhaled corticosteroids (n=391). 9

eFigure 7b. Sensitivity analyses excluding those who had a history of age-related macular degeneration or an ever history of use of oral / inhaled corticosteroids or cumulative history of any conditions (that may be indications for oral / inhaled corticosteroids (n=220)) 10

eTable 1. Associations (odds ratio (95%CI)) between plasma homocysteine and XFG risk in a nested case-control study* 12

eTable 2. Associations (odds ratio (95%CI)) between 379 metabolites and XFG risk in a nested case-control study 13

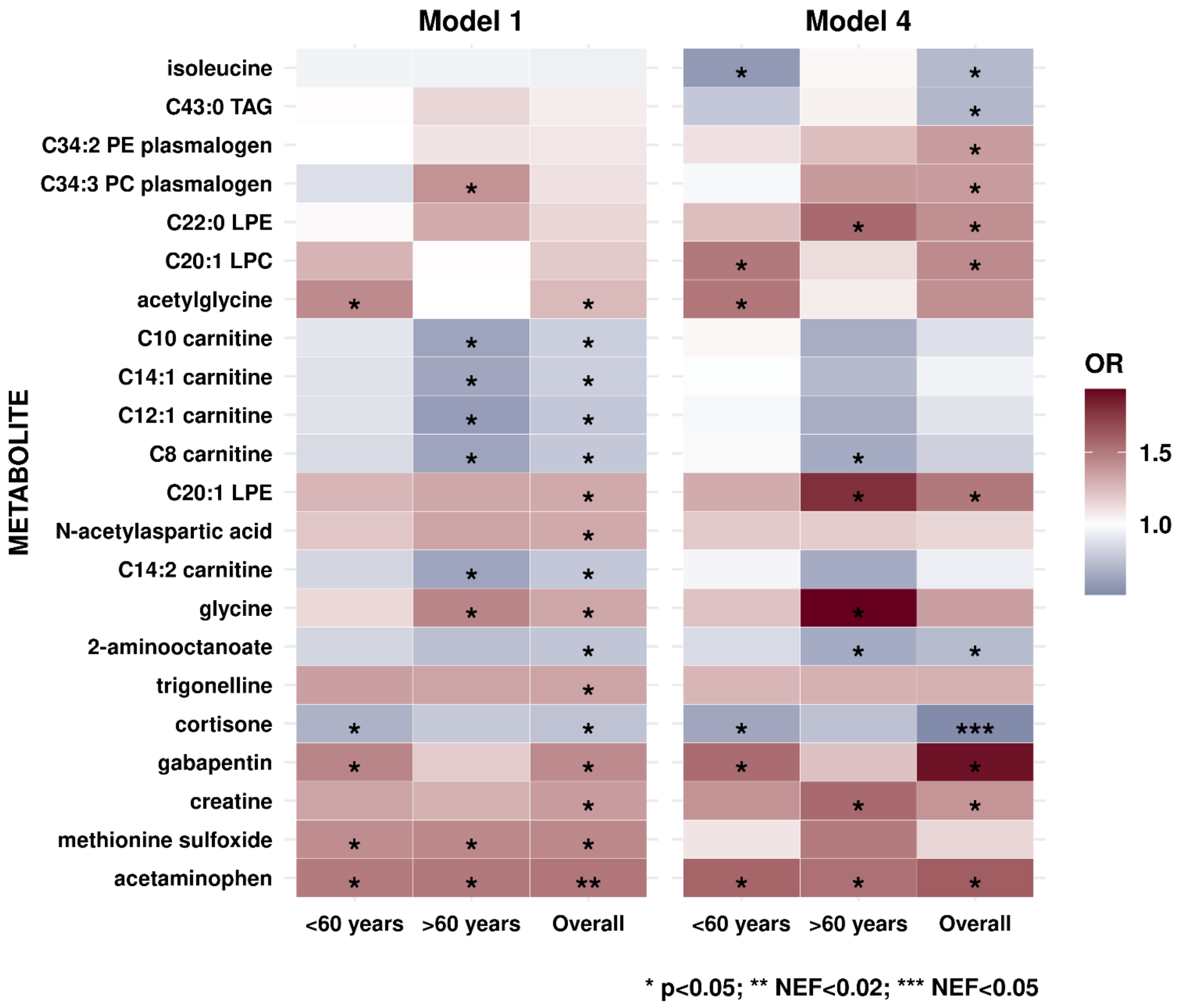


#### eFigure 1. Secondary analysis by age (< vs. ≥60 years; n=214 vs. n=196).

Metabolites that are nominally significant in either Model 1 or Model 4 are plotted (Model 5 and Model 6 did not converge due to smaller sample sizes for stratified analyses). **Model 1**: basic model, adjusting for matching factors only (see Table 1); **Model 4**: Model 1 + age, sex, smoking status, BMI, physical activity, fasting status, time of day of blood draw, month of blood draw, family history of glaucoma, type of Caucasian, time out in sunlight in the summer in youth, non-melanoma skin cancer, latitude, population density, folate intake, caffeine intake, alcohol intake, caloric intake. Models 5 and 6 did not converge due to smaller sample sizes in subgroup analysis. *p<0.05. ** Number of effective tests (NEF)<0.2; ** NEF<0.05.

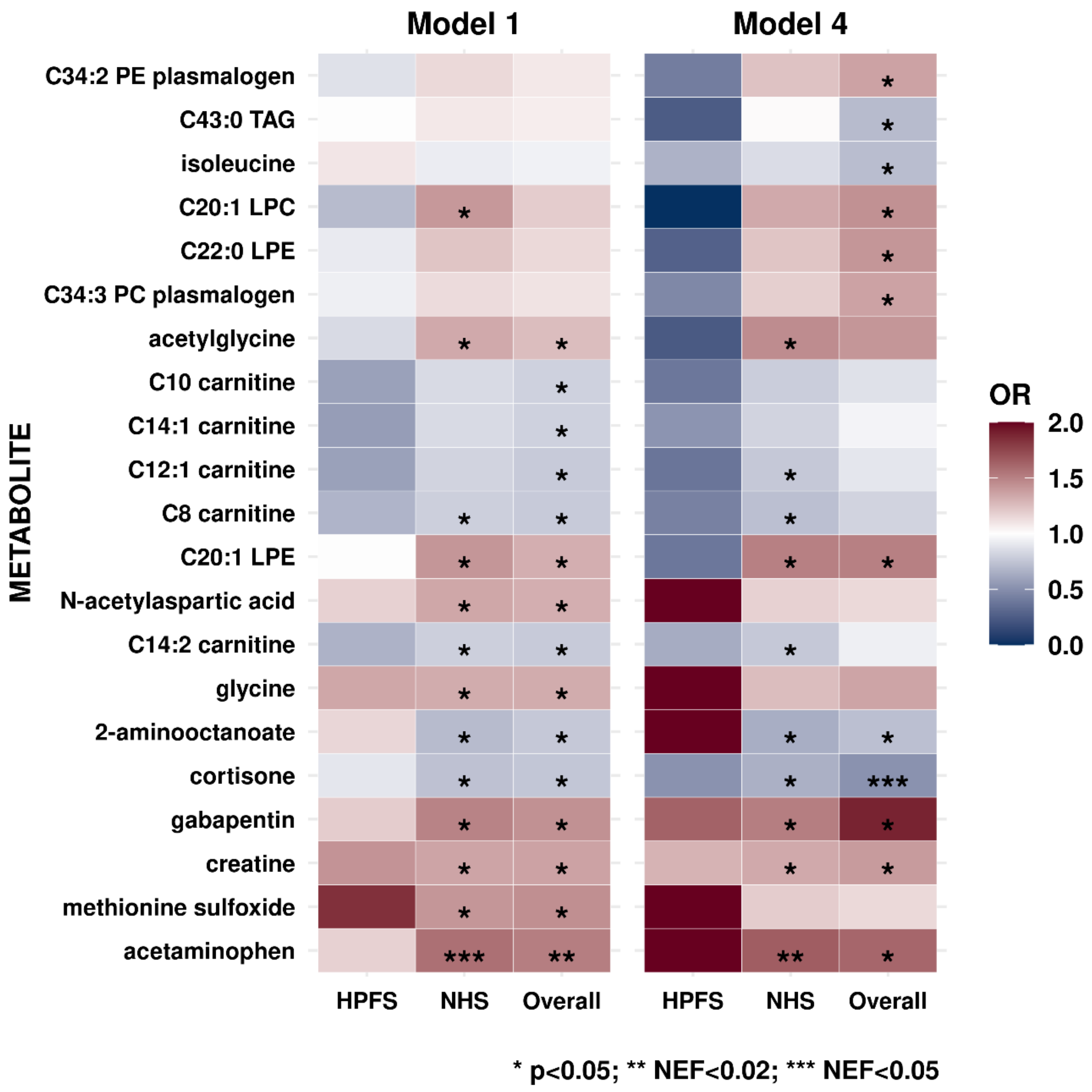


#### eFigure 2. Secondary analysis by sex (Nurses Health Study (NHS; n=348) participants are all women and Health Professionals Follow-up Study (HPFS; n=62) participants are all men).

Metabolites that are nominally significant in either Model 1 or Model 4 are plotted (Model 5 and Model 6 did not converge due to smaller sample sizes for stratified analyses). **Model 1**: basic model, adjusting for matching factors only (see Table 1); **Model 4**: Model 1 + age, smoking status, BMI, physical activity, fasting status, time of day of blood draw, month of blood draw, family history of glaucoma, type of Caucasian, time out in sunlight in the summer in youth, non-melanoma skin cancer, latitude, population density, folate intake, caffeine intake, alcohol intake, caloric intake. Models 5 and 6 did not converge due to smaller sample sizes in subgroup analysis. *p<0.05. ** Number of effective tests (NEF)<0.2; ** NEF<0.05.

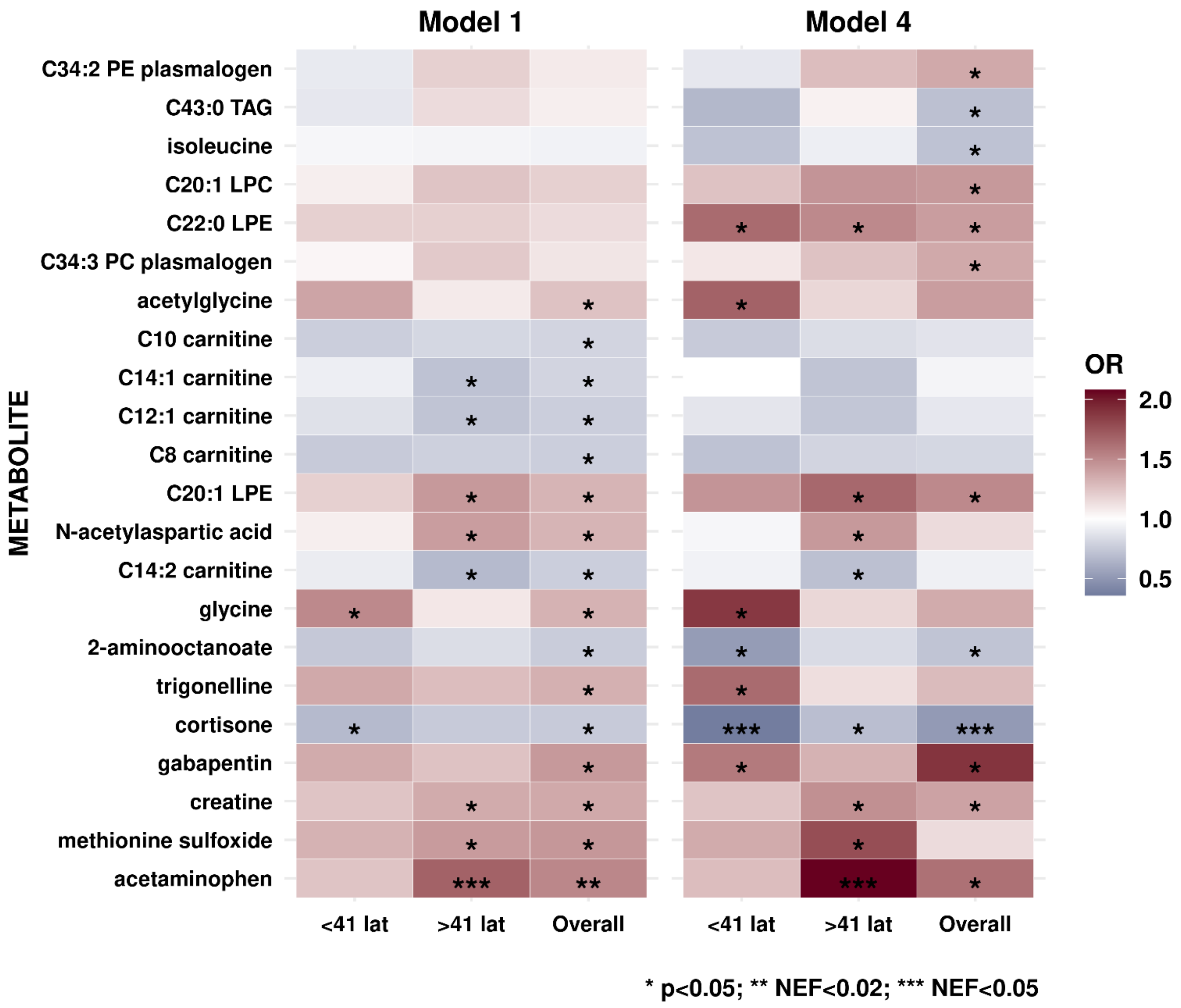


#### eFigure 3. Secondary analysis by latitude (<41°N in latitude vs. ≥41°N in latitude; n=182 vs. n=228).

Metabolites that are nominally significant in either Model 1 or Model 4 are plotted (Model 5 and Model 6 did not converge due to smaller sample sizes for stratified analyses). **Model 1**: basic model, adjusting for matching factors only (see Table 1); **Model 4**: Model 1 + age, sex, smoking status, BMI, physical activity, fasting status, time of day of blood draw, month of blood draw, family history of glaucoma, type of Caucasian, time out in sunlight in the summer in youth, non-melanoma skin cancer, population density, folate intake, caffeine intake, alcohol intake, caloric intake. Models 5 and 6 did not converge due to smaller sample sizes in subgroup analysis. *p<0.05. ** Number of effective tests (NEF)<0.2; ** NEF<0.05.

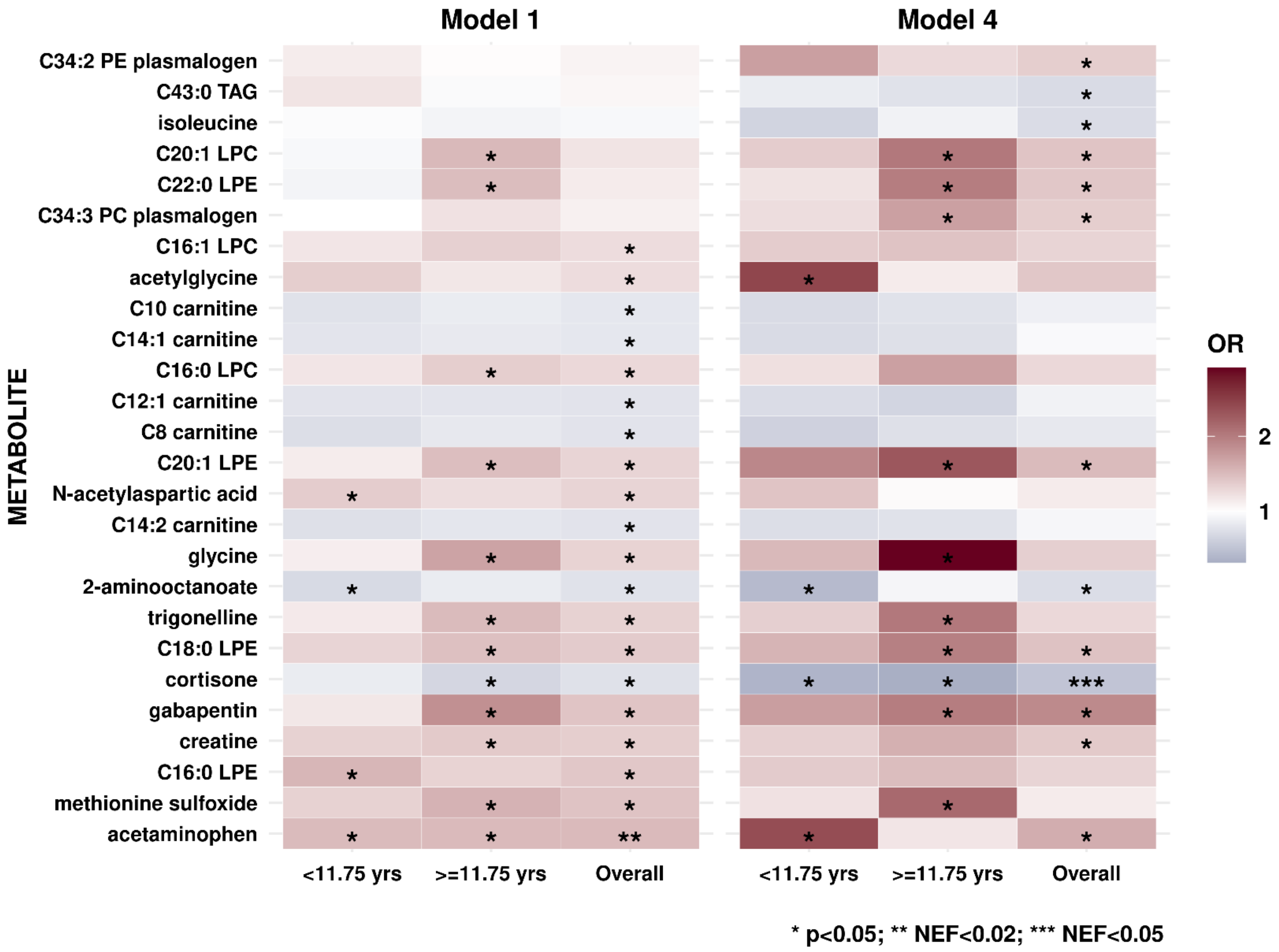


#### eFigure 4. Secondary analysis by time to diagnosis (<11.75 vs. ≥11.75 years; n=204 vs. 206).

Metabolites that are nominally significant in either Model 1 or Model 4 are plotted (Model 5 and Model 6 did not converge due to smaller sample sizes for stratified analyses). **Model 1**: basic model, adjusting for matching factors only (see Table 1); **Model 4**: Model 1 + age, sex, smoking status, BMI, physical activity, fasting status, time of day of blood draw, month of blood draw, family history of glaucoma, type of Caucasian, time out in sunlight in the summer in youth, non-melanoma skin cancer, latitude, population density, folate intake, caffeine intake, alcohol intake, caloric intake. Models 5 and 6 did not converge due to smaller sample sizes in subgroup analysis. *p<0.05. ** Number of effective tests (NEF)<0.2; ** NEF<0.05.

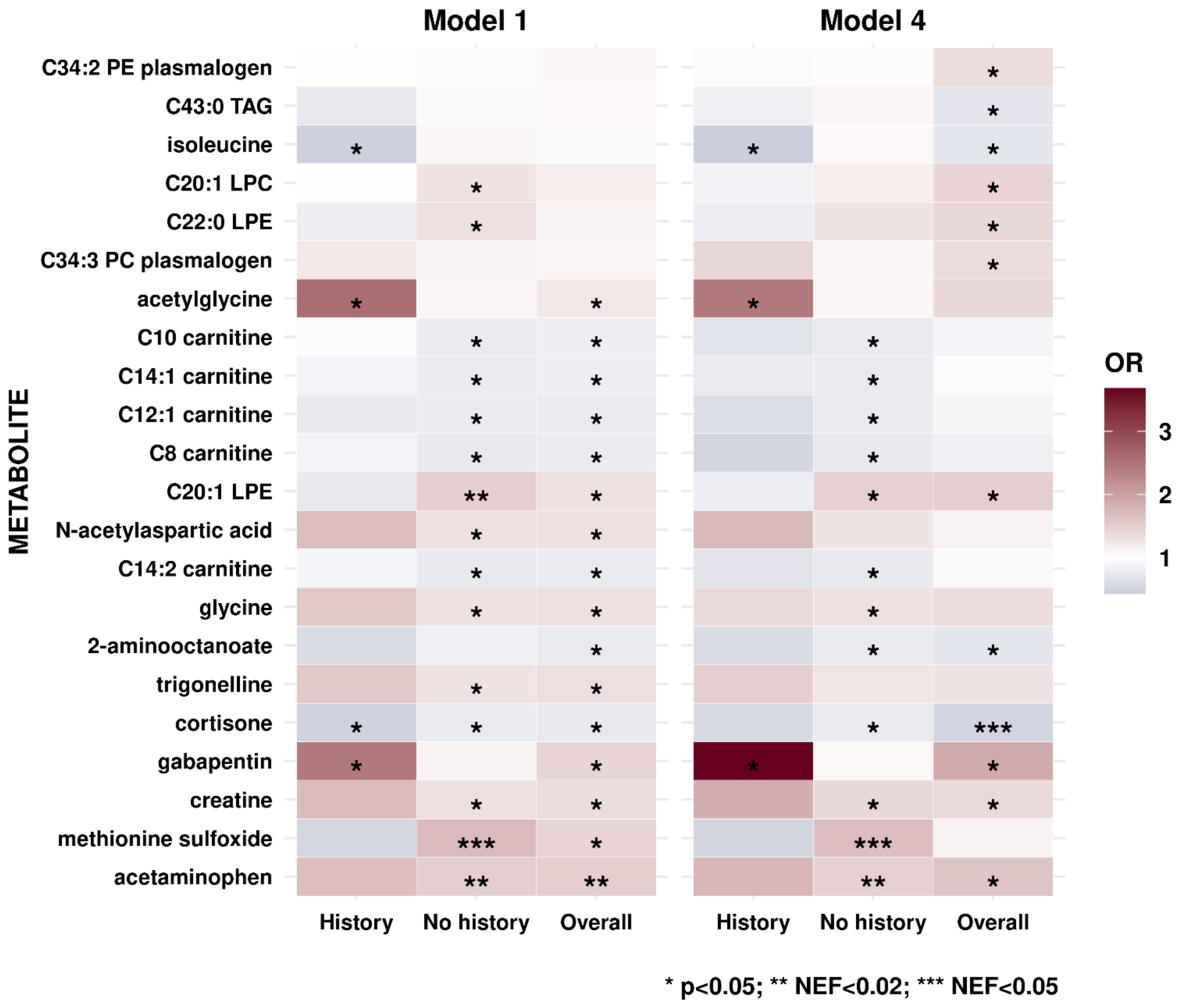


#### eFigure 5. Secondary analysis by self-reported glaucoma family history (yes vs. no; n=84 vs n=320).

Metabolites that are nominally significant in either Model 1 or Model 4 are plotted (Model 5 and Model 6 did not converge due to smaller sample sizes for stratified analyses). **Model 1**: basic model, adjusting for matching factors only (see Table 1); **Model 4**: Model 1 adjusts for age, sex, smoking status, BMI, physical activity, fasting status, time of day of blood draw, month of blood draw, type of Caucasian, time out in sunlight in the summer in youth, non-melanoma skin cancer, latitude, population density, folate intake, caffeine intake, alcohol intake, caloric intake. Models 5 and 6 did not converge due to smaller sample sizes in subgroup analysis. *p<0.05. ** Number of effective tests (NEF)<0.2; ** NEF<0.05.

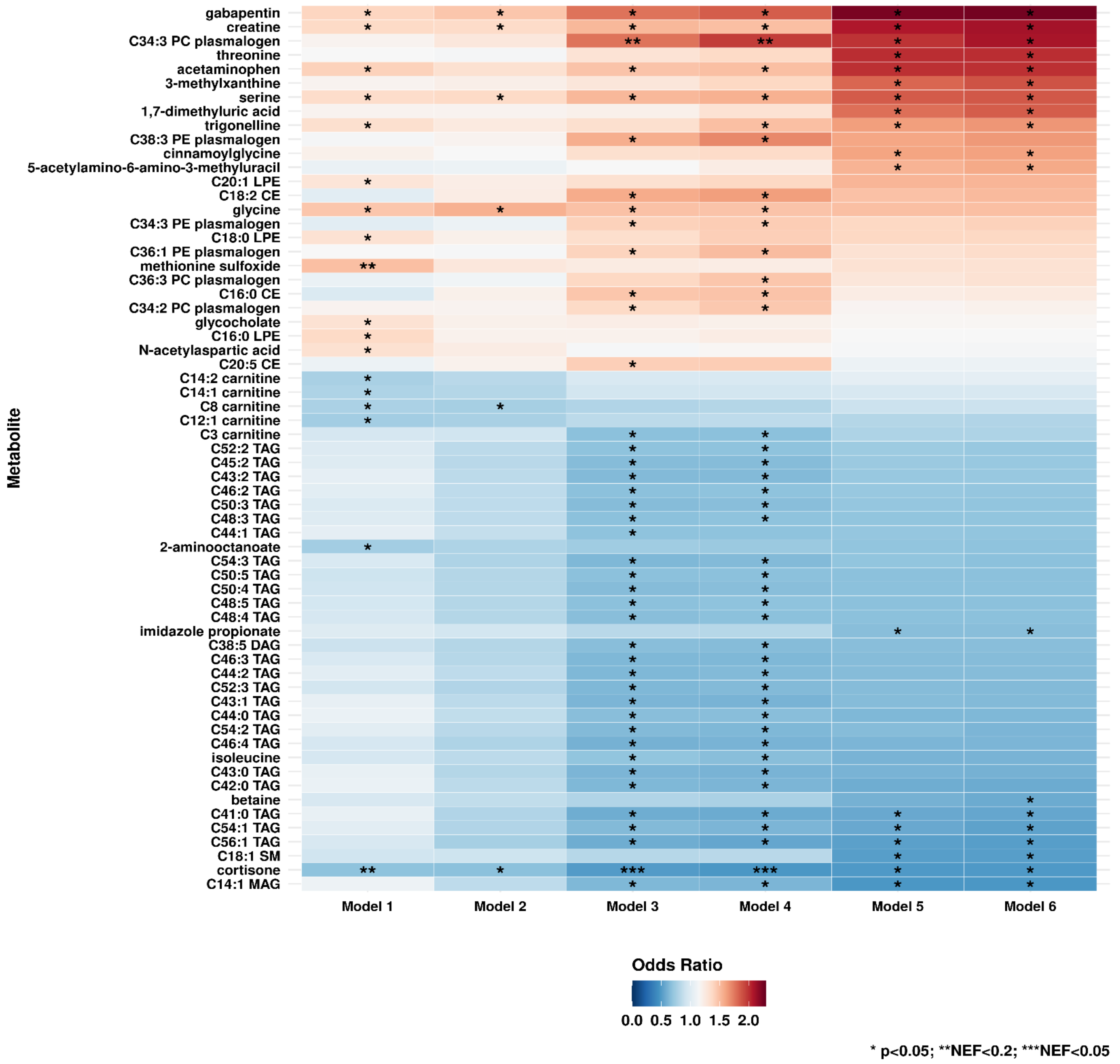


#### eFigure 6. Sensitivity analyses excluding those who were diagnosed within 5 years after blood draw (n=336).

**Model 1**: basic model, adjusting for matching factors only (see Table 1); **Model 2** (factors that affect metabolite levels): Model 1 + age, sex, smoking status, BMI, physical activity, fasting status, time of day of blood draw, month of blood draw; **Model 3** (presumed exfoliation syndrome risk factors): Model 2 + family history of glaucoma, type of Caucasian, time out in sunlight in the summer in youth, non-melanoma skin cancer, latitude, population density; **Model 4** (factors that may raise homocysteine levels): Model 3 + folate intake, caffeine intake, alcohol intake, caloric intake; **Model 5** (systemic comorbidities suggested to be associated with XFS in some studies): Model 4 + heart disease, hypertension, high cholesterol, hearing loss, diabetes, stroke, sleep duration; **Model 6** (use of drugs associated with glaucoma): Model 5 + steroid use. * Number of effective tests (NEF)<0.2; ** NEF<0.05; ** NEF<0.001.

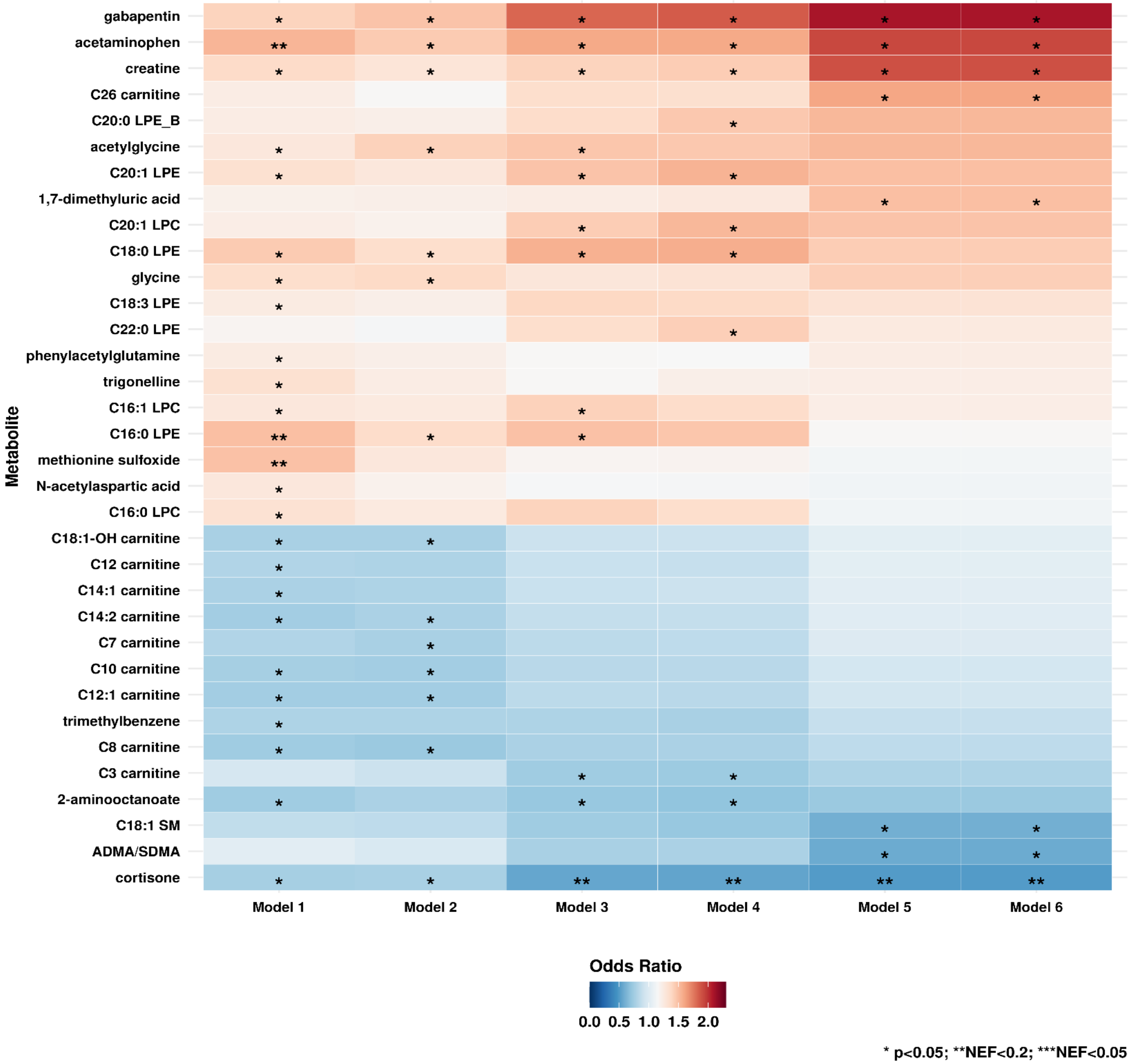


#### eFigure 7a. Sensitivity analyses excluding those who had a history of age-related macular degeneration or history of use of oral / inhaled corticosteroids (n=391)

**Model 1**: basic model, adjusting for matching factors only (see Table 1); **Model 2** (factors that affect metabolite levels): Model 1 + age, sex, smoking status, BMI, physical activity, fasting status, time of day of blood draw, month of blood draw; **Model 3** (presumed exfoliation syndrome risk factors): Model 2 + family history of glaucoma, type of Caucasian, time out in sunlight in the summer in youth, non-melanoma skin cancer, latitude, population density; **Model 4** (factors that may raise homocysteine levels): Model 3 + folate intake, caffeine intake, alcohol intake, caloric intake; **Model 5** (systemic comorbidities suggested to be associated with XFS in some studies): Model 4 + heart disease, hypertension, high cholesterol, hearing loss, diabetes, stroke, sleep duration; **Model 6** (use of drugs associated with glaucoma): Model 5 + steroid use. * Number of effective tests (NEF)<0.2; ** NEF<0.05; ** NEF<0.001.

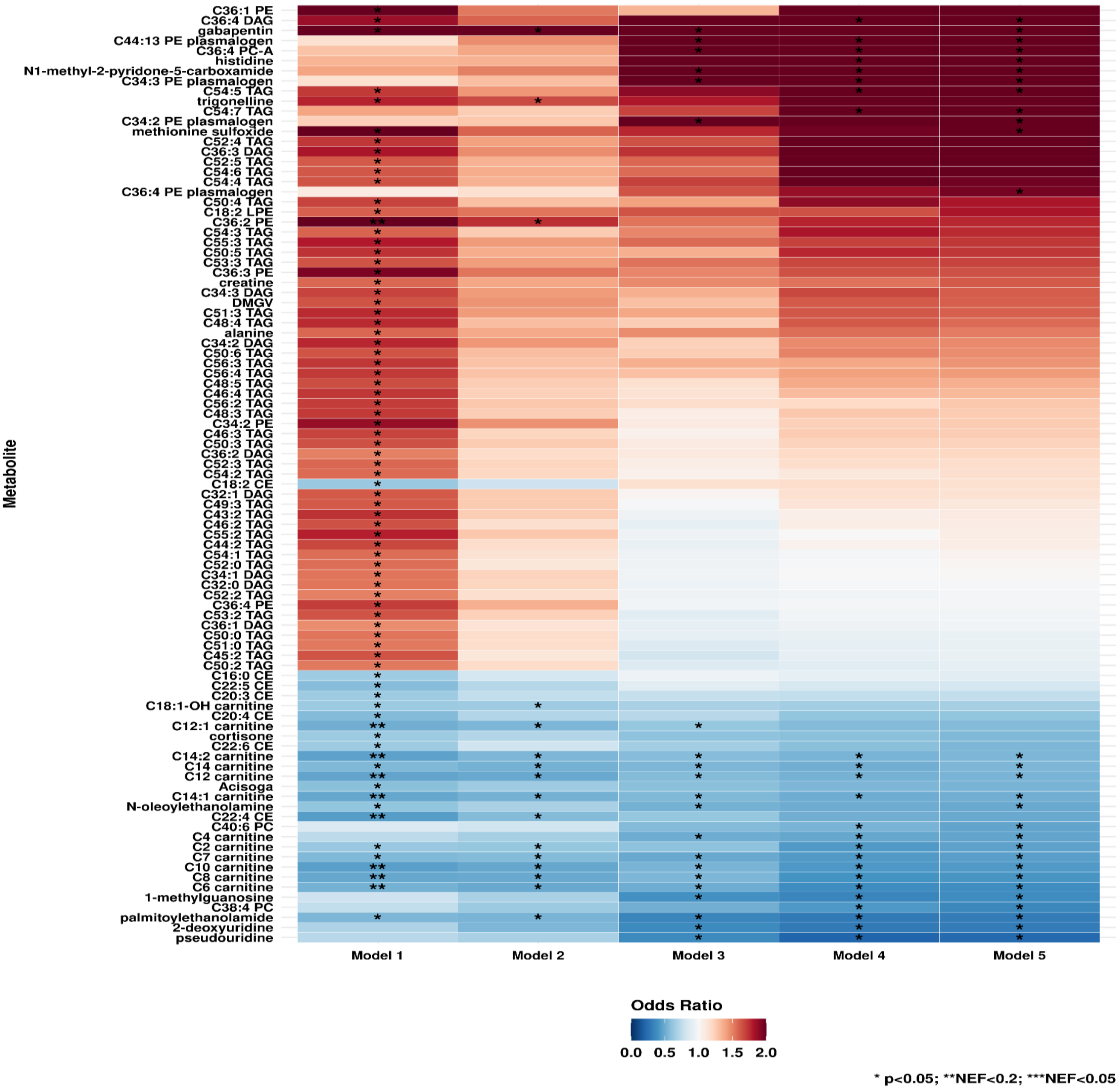


eFigure 7b. Sensitivity analyses excluding those who had a history of age-related macular degeneration or an ever history of use of oral / inhaled corticosteroids or cumulative history of any conditions (that may be indications for oral / inhaled corticosteroids (n=220))

Self-reported conditions that are indications for corticosteroid use included asthma, COPD, gout, hyperthyroidism, hypothyroidism, inflammatory bowel disease, leukemia or lymphoma, multiple sclerosis, osteoarthritis, psoriasis, rheumatoid arthritis, systemic lupus erythematosus and vitiligo.

**Model 1**: basic model, adjusting for matching factors only (see Table 1); **Model 2** (factors that affect metabolite levels): Model 1 + age, sex, smoking status, BMI, physical activity, fasting status, time of day of blood draw, month of blood draw; **Model 3** (presumed exfoliation syndrome risk factors): Model 2 + family history of glaucoma, type of Caucasian, time out in sunlight in the summer in youth, non-melanoma skin cancer, latitude, population density; **Model 4** (factors that may raise homocysteine levels): Model 3 + folate intake, caffeine intake, alcohol intake, caloric intake; **Model 5** (systemic comorbidities suggested to be associated with XFS in some studies): Model 4 + heart disease, hypertension, high cholesterol, hearing loss, diabetes, stroke, sleep duration; **Model 6** (use of drugs associated with glaucoma): Model 5 + steroid use. * Number of effective tests (NEF)<0.2; ** NEF<0.05; ** NEF<0.001.

#### eTable 1. Associations (odds ratio (95%CI)) between plasma homocysteine and XFG risk in a nested case-control study*

| | Cases (n=203) | Controls (n=203) |
|---|---|---|
| Mean homocysteine level (SD), µmol/L | 13.3 (4.1) | 13.1 (4.5) |

| METABOLITE | HMDB_ID | Metabolite class | Model 1 | Model 2 | Model 3 | Model 4 | Model 5 | Model 6 |
|---|---|---|---|---|---|---|---|---|
| | | | OR (95%CI) | OR (95%CI) | OR (95%CI) | OR (95%CI) | OR (95%CI) | OR (95%CI) |
| homocysteine | HMDB0000742 | Organic acids and derivatives | 1.05 (0.85, 1.29) | 1.16 (0.89, 1.52) | 1.18 (0.89, 1.55) | 1.21 (0.90, 1.62) | 1.23 (0.91, 1.66) | 1.22 (0.90, 1.65) |

* Homocysteine was not assessed as part of the metabolomic platform; homocysteine levels were measured in a separate specific assay, lab-batch corrected and available among a subset of n=203 cases and n=203 matched controls (matching factors are the same as those described in Table 1).

**Model 1**: basic model, adjusting for matching factors only (see Table 1); **Model 2** (factors that affect metabolite levels): Model 1 + age, sex, smoking status, BMI, physical activity, fasting status, time of day of blood draw, month of blood draw; **Model 3** (presumed exfoliation syndrome risk factors): Model 2 + family history of glaucoma, type of Caucasian, time out in sunlight in the summer in youth, non-melanoma skin cancer, latitude, population density; **Model 4** (factors that may raise homocysteine levels): Model 3 + folate intake, caffeine intake, alcohol intake, caloric intake; **Model 5** (systemic comorbidities suggested to be associated with XFS in some studies): Model 4 + heart disease, hypertension, high cholesterol, hearing loss, diabetes, stroke, sleep duration; **Model 6** (use of drugs associated with glaucoma): Model 5 + steroid use.

### eTable 2. Associations (odds ratio (95%CI)) between 379 metabolites and XFG risk in a nested case-control study

| METABOLITE | HMDB_ID | Metabolite class | Model 1 | Model 2 | Model 3 | Model 4 | | | Model 5 | Model 6 | | |
|---|---|---|---|---|---|---|---|---|---|---|---|---|
| | | | OR (95%CI) | OR (95%CI) | OR (95%CI) | OR (95%CI) | p-value | NEF | OR (95%CI) | OR (95%CI) | p-value | NEF |
| 1,7-dimethyluric acid | HMDB0011103 | Organoheterocyclic compounds | 1.21 (0.98-1.48) | 1.24 (0.98-1.56) | 1.28 (0.97-1.67) | 1.32 (0.98-1.78) | 0.07 | >0.99 | 1.61 (1.11-2.35) | 1.65 (1.12-2.42) | 0.01 | 0.76 |
| 1-methyladenosine | HMDB0003331 | NA | 1.24 (0.71-2.16) | NA | NA | NA | NA | NA | NA | NA | NA | NA |
| 1-methylguanine | HMDB0003282 | NA | 1.00 (0.80-1.25) | 0.96 (0.73-1.25) | 0.83 (0.58-1.18) | 0.81 (0.57-1.17) | 0.27 | >0.99 | 0.89 (0.58-1.36) | 0.88 (0.57-1.36) | 0.56 | >0.99 |
| 1-methylguanosine | HMDB0001563 | NA | 1.01 (0.82-1.26) | 1.01 (0.78-1.31) | 0.82 (0.60-1.13) | 0.82 (0.59-1.14) | 0.24 | >0.99 | 0.91 (0.61-1.36) | 0.91 (0.61-1.36) | 0.64 | >0.99 |
| 1-methylhistamine | HMDB0000898 | NA | 0.91 (0.75-1.11) | 0.88 (0.70-1.11) | 0.89 (0.68-1.18) | 0.90 (0.68-1.19) | 0.44 | >0.99 | 0.91 (0.65-1.27) | 0.90 (0.64-1.27) | 0.55 | >0.99 |
| 1-methylnicotinamide | HMDB0000699 | Pyridines and derivatives | 0.98 (0.80-1.20) | 1.02 (0.81-1.29) | 1.14 (0.85-1.54) | 1.13 (0.82-1.55) | 0.45 | >0.99 | 1.21 (0.81-1.81) | 1.21 (0.81-1.81) | 0.36 | >0.99 |
| 2-aminohippuric acid | NA | NA | 1.05 (0.86-1.27) | 0.99 (0.79-1.25) | 0.97 (0.74-1.27) | 0.96 (0.73-1.28) | 0.80 | >0.99 | 1.04 (0.75-1.44) | 1.04 (0.75-1.44) | 0.82 | >0.99 |
| 2-aminoisobutyric acid/GABA | HMDB0001906* | Organic acids and derivatives | 1.05 (0.85-1.29) | 1.12 (0.89-1.42) | 1.16 (0.87-1.54) | 1.15 (0.86-1.54) | 0.35 | >0.99 | 1.28 (0.90-1.83) | 1.31 (0.91-1.88) | 0.15 | >0.99 |
| 2-aminooctanoate | HMDB0000991 | Organic acids and derivatives | 0.76 (0.61-0.94) | 0.79 (0.62-1.00) | 0.74 (0.55-1.00) | 0.72 (0.52-0.97) | 0.03 | >0.99 | 0.74 (0.51-1.06) | 0.73 (0.51-1.06) | 0.10 | >0.99 |
| 2-deoxycytidine | HMDB0000014 | NA | 1.04 (0.83-1.29) | 0.99 (0.78-1.28) | 0.98 (0.73-1.32) | 0.99 (0.73-1.35) | 0.95 | >0.99 | 1.09 (0.74-1.59) | 1.09 (0.74-1.59) | 0.67 | >0.99 |
| 2-deoxyuridine | HMDB0000012 | NA | 0.92 (0.73-1.16) | 0.87 (0.66-1.15) | 0.84 (0.60-1.17) | 0.83 (0.59-1.17) | 0.28 | >0.99 | 0.83 (0.54-1.27) | 0.82 (0.54-1.26) | 0.37 | >0.99 |
| 3-(N-acetyl-L-cystein-S-yl) acetaminophen | NA | NA | 1.07 (0.54-2.11) | NA | NA | NA | NA | NA | NA | NA | NA | NA |
| 3-hydroxyanthranilic acid | HMDB0001476 | Benzenoids | 1.00 (0.82-1.23) | 0.95 (0.75-1.20) | 0.85 (0.63-1.13) | 0.85 (0.63-1.15) | 0.30 | >0.99 | 0.93 (0.65-1.34) | 0.93 (0.65-1.34) | 0.70 | >0.99 |
| 3-methylhistidine | HMDB0000479 | Organic acids and derivatives | 1.11 (0.90-1.36) | 1.12 (0.89-1.42) | 1.07 (0.82-1.40) | 1.05 (0.80-1.38) | 0.72 | >0.99 | 1.00 (0.71-1.40) | 1.00 (0.70-1.42) | >0.99 | >0.99 |
| 3-methylxanthine | HMDB0001886 | Organoheterocyclic compounds | 1.21 (0.99-1.48) | 1.27 (1.02-1.60) | 1.26 (0.96-1.65) | 1.27 (0.95-1.69) | 0.11 | >0.99 | 1.46 (1.03-2.08) | 1.49 (1.04-2.13) | 0.03 | >0.99 |
| 4-acetamidobutanoate | HMDB0003681 | Organic acids and derivatives | 1.13 (0.92-1.38) | 0.98 (0.78-1.25) | 1.00 (0.75-1.34) | 0.96 (0.71-1.31) | 0.81 | >0.99 | 0.94 (0.66-1.33) | 0.94 (0.66-1.33) | 0.72 | >0.99 |
| 4-hydroxyhippurate | HMDB0013678 | Benzenoids | 1.05 (0.86-1.30) | 1.07 (0.85-1.36) | 0.97 (0.74-1.28) | 0.96 (0.72-1.28) | 0.79 | >0.99 | 0.97 (0.70-1.36) | 0.97 (0.70-1.35) | 0.86 | >0.99 |
| 5-acetylamino-6-amino-3-methyluracil | HMDB0004400 | Organic nitrogen compounds | 1.12 (0.92-1.36) | 1.13 (0.91-1.41) | 1.20 (0.93-1.55) | 1.26 (0.94-1.68) | 0.12 | >0.99 | 1.45 (1.02-2.07) | 1.47 (1.02-2.11) | 0.04 | >0.99 |
| 5-hydroxytryptophol | HMDB0001855 | Organoheterocyclic compounds | 0.88 (0.54-1.44) | NA | NA | NA | NA | NA | NA | NA | NA | NA |
| 7-dehydrodesmosterol | HMDB0003896 | NA | 0.98 (0.77-1.24) | 0.96 (0.73-1.25) | 1.08 (0.77-1.51) | 1.03 (0.73-1.46) | 0.86 | >0.99 | 0.91 (0.60-1.39) | 0.91 (0.60-1.39) | 0.67 | >0.99 |
| 7-methylguanine | HMDB0000897 | Organoheterocyclic compounds | 1.01 (0.82-1.25) | 1.01 (0.78-1.30) | 1.02 (0.74-1.40) | 0.98 (0.70-1.36) | 0.89 | >0.99 | 1.01 (0.69-1.48) | 1.01 (0.68-1.49) | 0.97 | >0.99 |
| acetaminophen | HMDB0001859 | Phenols | 1.52 (1.19-1.94) | 1.43 (1.10-1.87) | 1.60 (1.13-2.25) | 1.62 (1.14-2.29) | 0.01 | 0.01 | 1.91 (1.22-3.00) | 1.92 (1.22-3.02) | 0.005 | 0.32 |
| acetaminophen glucuronide | HMDB0010316 | NA | 1.78 (0.85-3.75) | NA | NA | NA | NA | NA | NA | NA | NA | NA |
| acetyl-galactosamine | HMDB0000853 | NA | 1.02 (0.83-1.26) | 0.92 (0.72-1.17) | 0.83 (0.61-1.12) | 0.81 (0.59-1.10) | 0.18 | >0.99 | 0.97 (0.68-1.38) | 0.96 (0.67-1.39) | 0.84 | >0.99 |
| acetylglycine | HMDB0000532 | NA | 1.26 (1.01-1.57) | 1.35 (1.03-1.76) | 1.45 (1.02-2.05) | 1.42 (1.00-2.02) | 0.05 | >0.99 | 1.51 (0.99-2.30) | 1.51 (0.99-2.30) | 0.06 | >0.99 |
| Acisoga | HMDB0061384 | NA | 0.86 (0.69-1.06) | 0.86 (0.68-1.09) | 0.81 (0.61-1.08) | 0.82 (0.61-1.10) | 0.18 | >0.99 | 0.98 (0.69-1.38) | 0.98 (0.69-1.38) | 0.89 | >0.99 |
| ADMA/SDMA | HMDB0001539* | Organic acids and derivatives | 1.06 (0.85-1.32) | 1.01 (0.78-1.30) | 0.86 (0.63-1.18) | 0.85 (0.62-1.17) | 0.33 | >0.99 | 0.68 (0.44-1.05) | 0.68 (0.44-1.05) | 0.08 | >0.99 |
| alanine | HMDB0000161 | Carboxylic acids and derivatives | 1.05 (0.85-1.30) | 1.01 (0.79-1.28) | 0.87 (0.65-1.17) | 0.86 (0.64-1.17) | 0.33 | >0.99 | 0.88 (0.62-1.26) | 0.88 (0.62-1.26) | 0.49 | >0.99 |

| METABOLITE | HMDB_ID | Metabolite class | Model 1 | Model 2 | Model 3 | Model 4 | | | Model 5 | Model 6 | | |
|---|---|---|---|---|---|---|---|---|---|---|---|---|
| | | | OR (95%CI) | OR (95%CI) | OR (95%CI) | OR (95%CI) | p-value | NEF | OR (95%CI) | OR (95%CI) | p-value | NEF |
| allantoin | HMDB0000462 | Azoles | 1.15 (0.93-1.42) | 0.98 (0.77-1.25) | 0.91 (0.68-1.22) | 0.90 (0.67-1.21) | 0.48 | >0.99 | 0.92 (0.65-1.30) | 0.92 (0.65-1.31) | 0.65 | >0.99 |
| anserine | HMDB0000194 | NA | 0.47 (0.14-1.56) | NA | NA | NA | NA | NA | NA | NA | NA | NA |
| arginine | HMDB0000517 | NA | 1.18 (0.69-2.01) | NA | NA | NA | NA | NA | NA | NA | NA | NA |
| asparagine | HMDB0000168 | Organic acids and derivatives | 0.93 (0.75-1.15) | 0.89 (0.70-1.13) | 0.92 (0.68-1.24) | 0.90 (0.66-1.22) | 0.49 | >0.99 | 0.87 (0.59-1.29) | 0.87 (0.59-1.29) | 0.49 | >0.99 |
| atenolol | HMDB0001924 | NA | NA | NA | NA | NA | NA | NA | NA | NA | NA | NA |
| betaine | HMDB0000043 | Carboxylic acids and derivatives | 0.92 (0.73-1.16) | 0.80 (0.61-1.05) | 0.77 (0.56-1.07) | 0.79 (0.56-1.10) | 0.16 | >0.99 | 0.65 (0.43-0.99) | 0.64 (0.42-0.98) | 0.04 | >0.99 |
| bilirubin | HMDB0000054 | NA | 0.96 (0.78-1.19) | 0.98 (0.77-1.25) | 1.02 (0.76-1.36) | 1.05 (0.78-1.41) | 0.74 | >0.99 | 1.06 (0.74-1.51) | 1.06 (0.74-1.51) | 0.76 | >0.99 |
| biliverdin | HMDB0001008 | Organoheterocyclic compounds | 0.98 (0.79-1.22) | 1.03 (0.81-1.31) | 1.28 (0.94-1.75) | 1.31 (0.96-1.80) | 0.09 | >0.99 | 1.28 (0.86-1.89) | 1.28 (0.86-1.89) | 0.22 | >0.99 |
| butyrobetaine | HMDB0001161 | NA | 0.91 (0.73-1.13) | 0.81 (0.64-1.04) | 0.88 (0.66-1.19) | 0.90 (0.67-1.22) | 0.50 | >0.99 | 0.90 (0.63-1.29) | 0.90 (0.63-1.29) | 0.56 | >0.99 |
| C10 carnitine | HMDB0000651 | Carnitines | 0.80 (0.65-0.99) | 0.79 (0.62-1.01) | 0.87 (0.65-1.15) | 0.87 (0.65-1.16) | 0.35 | >0.99 | 0.98 (0.70-1.38) | 0.98 (0.69-1.38) | 0.90 | >0.99 |
| C10:2 carnitine | HMDB0013325 | Carnitines | 1.07 (0.87-1.32) | 0.97 (0.76-1.23) | 0.99 (0.74-1.33) | 1.00 (0.74-1.35) | 0.99 | >0.99 | 1.02 (0.71-1.47) | 1.02 (0.71-1.47) | 0.91 | >0.99 |
| C12 carnitine | HMDB0002250 | Carnitines | 0.83 (0.68-1.01) | 0.82 (0.65-1.04) | 0.94 (0.72-1.22) | 0.93 (0.71-1.22) | 0.60 | >0.99 | 1.05 (0.75-1.47) | 1.05 (0.75-1.47) | 0.78 | >0.99 |
| C12:1 carnitine | HMDB0013326* | Carnitines | 0.77 (0.63-0.96) | 0.79 (0.63-1.00) | 0.90 (0.68-1.19) | 0.89 (0.67-1.19) | 0.43 | >0.99 | 0.96 (0.68-1.35) | 0.96 (0.67-1.36) | 0.80 | >0.99 |
| C14 carnitine | HMDB0005066 | Carnitines | 0.93 (0.77-1.14) | 0.92 (0.74-1.15) | 1.03 (0.79-1.34) | 1.03 (0.79-1.35) | 0.81 | >0.99 | 1.25 (0.90-1.74) | 1.26 (0.89-1.77) | 0.19 | >0.99 |
| C14:0 CE | HMDB0006725 | Cholesteryl esters | 1.16 (0.94-1.44) | 1.22 (0.94-1.58) | 1.27 (0.93-1.74) | 1.25 (0.90-1.72) | 0.18 | >0.99 | 1.13 (0.73-1.73) | 1.13 (0.73-1.74) | 0.57 | >0.99 |
| C14:0 LPC | HMDB0010379 | Lysophosphatidylcholines | 1.20 (0.96-1.49) | 1.23 (0.96-1.58) | 1.25 (0.92-1.69) | 1.24 (0.90-1.69) | 0.18 | >0.99 | 1.21 (0.79-1.84) | 1.21 (0.79-1.85) | 0.37 | >0.99 |
| C14:0 SM | HMDB0012097 | Sphingomyelins | 1.20 (0.95-1.52) | 1.28 (0.98-1.68) | 1.34 (0.98-1.84) | 1.33 (0.96-1.84) | 0.09 | >0.99 | 1.30 (0.86-1.95) | 1.30 (0.86-1.96) | 0.21 | >0.99 |
| C14:1 carnitine | HMDB0002014* | Carnitines | 0.80 (0.65-0.98) | 0.82 (0.65-1.03) | 0.94 (0.71-1.24) | 0.95 (0.71-1.25) | 0.70 | >0.99 | 1.05 (0.75-1.46) | 1.04 (0.74-1.47) | 0.80 | >0.99 |
| C14:1 MAG | HMDB0011562* | NA | 1.15 (0.90-1.46) | 1.00 (0.76-1.32) | 0.96 (0.69-1.34) | 1.00 (0.71-1.40) | 0.98 | >0.99 | 0.89 (0.59-1.35) | 0.89 (0.59-1.35) | 0.59 | >0.99 |
| C14:2 carnitine | HMDB0013331* | Carnitines | 0.77 (0.63-0.95) | 0.80 (0.63-1.01) | 0.92 (0.70-1.20) | 0.93 (0.70-1.23) | 0.60 | >0.99 | 1.05 (0.75-1.47) | 1.05 (0.75-1.47) | 0.77 | >0.99 |
| C16 carnitine | HMDB0000222 | NA | 0.77 (0.44-1.35) | NA | NA | NA | NA | NA | NA | NA | NA | NA |
| C16:0 CE | HMDB0000885 | Cholesteryl esters | 0.90 (0.73-1.10) | 1.03 (0.81-1.30) | 1.16 (0.87-1.55) | 1.16 (0.87-1.55) | 0.31 | >0.99 | 1.05 (0.74-1.50) | 1.06 (0.74-1.51) | 0.77 | >0.99 |
| C16:0 Ceramide (d18:1) | HMDB0004949 | Ceramides | 1.02 (0.82-1.25) | 0.90 (0.71-1.13) | 1.00 (0.75-1.34) | 0.99 (0.73-1.33) | 0.93 | >0.99 | 1.03 (0.72-1.47) | 1.03 (0.72-1.47) | 0.89 | >0.99 |
| C16:0 LPC | HMDB0010382 | Lysophosphatidylcholines | 1.29 (1.02-1.62) | 1.21 (0.94-1.57) | 1.35 (0.98-1.85) | 1.29 (0.91-1.83) | 0.16 | >0.99 | 1.02 (0.64-1.62) | 1.02 (0.64-1.62) | 0.94 | >0.99 |
| C16:0 LPE | HMDB0011503 | Lysophosphatidylethanolamines | 1.42 (1.13-1.78) | 1.26 (0.98-1.62) | 1.33 (0.98-1.81) | 1.32 (0.92-1.90) | 0.13 | >0.99 | 1.03 (0.63-1.68) | 1.03 (0.63-1.68) | 0.91 | >0.99 |
| C16:0 SM | HMDB0010169 | Sphingomyelins | 1.07 (0.87-1.32) | 1.04 (0.82-1.31) | 1.16 (0.89-1.53) | 1.13 (0.85-1.51) | 0.40 | >0.99 | 1.02 (0.68-1.52) | 1.02 (0.68-1.52) | 0.93 | >0.99 |
| C16:1 CE | HMDB0000658* | Cholesteryl esters | 1.10 (0.91-1.34) | 1.20 (0.96-1.50) | 1.33 (1.00-1.76) | 1.28 (0.95-1.71) | 0.11 | >0.99 | 1.02 (0.73-1.43) | 1.02 (0.73-1.44) | 0.89 | >0.99 |
| C16:1 LPC | HMDB0010383* | Lysophosphatidylcholines | 1.26 (1.01-1.57) | 1.24 (0.96-1.59) | 1.37 (1.00-1.90) | 1.32 (0.94-1.84) | 0.11 | >0.99 | 1.18 (0.77-1.81) | 1.18 (0.77-1.81) | 0.46 | >0.99 |
| C16:1 LPC plasmalogen | HMDB0010407* | LPC plasmalogens | 1.13 (0.89-1.43) | 1.10 (0.84-1.44) | 1.33 (0.96-1.85) | 1.37 (0.95-1.98) | 0.09 | >0.99 | 1.33 (0.86-2.06) | 1.33 (0.86-2.06) | 0.20 | >0.99 |
| C16:1 MAG | HMDB0011565* | NA | 1.07 (0.61-1.90) | NA | NA | NA | NA | NA | NA | NA | NA | NA |
| C16:1 SM | HMDB0029216 | Sphingomyelins | 1.08 (0.85-1.37) | 1.11 (0.84-1.46) | 1.15 (0.83-1.60) | 1.12 (0.79-1.58) | 0.52 | >0.99 | 0.89 (0.54-1.47) | 0.89 (0.54-1.47) | 0.65 | >0.99 |

| METABOLITE | HMDB_ID | Metabolite class | Model 1 | Model 2 | Model 3 | Model 4 | | | Model 5 | Model 6 | | |
|---|---|---|---|---|---|---|---|---|---|---|---|---|
| | | | OR (95%CI) | OR (95%CI) | OR (95%CI) | OR (95%CI) | p-value | NEF | OR (95%CI) | OR (95%CI) | p-value | NEF |
| C18 carnitine | HMDB0000848 | NA | 0.88 (0.51-1.53) | NA | NA | NA | NA | NA | NA | NA | NA | NA |
| C18:0 CE | HMDB0010368 | Cholesteryl esters | 0.93 (0.75-1.14) | 0.92 (0.73-1.16) | 0.92 (0.69-1.24) | 0.93 (0.69-1.26) | 0.65 | >0.99 | 0.79 (0.55-1.15) | 0.79 (0.55-1.15) | 0.22 | >0.99 |
| C18:0 LPC | HMDB0010384 | Lysophosphatidylcholines | 1.21 (0.95-1.55) | 1.15 (0.87-1.52) | 1.39 (0.98-1.97) | 1.37 (0.95-1.96) | 0.09 | >0.99 | 1.15 (0.72-1.83) | 1.15 (0.72-1.83) | 0.56 | >0.99 |
| C18:0 LPE | HMDB0011130 | Lysophosphatidylethanolamines | 1.39 (1.10-1.76) | 1.28 (0.98-1.67) | 1.45 (1.04-2.01) | 1.46 (1.01-2.12) | 0.04 | >0.99 | 1.30 (0.81-2.08) | 1.30 (0.81-2.08) | 0.28 | >0.99 |
| C18:0 LPE_B | HMDB0011130* | Lysophosphatidylethanolamines | 0.95 (0.57-1.56) | NA | NA | NA | NA | NA | NA | NA | NA | NA |
| C18:0 MAG | HMDB0011131 | NA | 0.85 (0.42-1.71) | NA | NA | NA | NA | NA | NA | NA | NA | NA |
| C18:0 SM | HMDB0001348 | Sphingomyelins | 0.99 (0.80-1.22) | 1.02 (0.81-1.29) | 1.03 (0.78-1.37) | 0.99 (0.73-1.32) | 0.92 | >0.99 | 0.95 (0.66-1.37) | 0.95 (0.66-1.37) | 0.80 | >0.99 |
| C18:1 carnitine | HMDB0005065* | NA | 0.79 (0.48-1.32) | NA | NA | NA | NA | NA | NA | NA | NA | NA |
| C18:1 CE | HMDB0000918* | Cholesteryl esters | 1.00 (0.81-1.24) | 1.12 (0.87-1.43) | 1.19 (0.88-1.61) | 1.14 (0.83-1.56) | 0.43 | >0.99 | 0.83 (0.52-1.32) | 0.79 (0.49-1.29) | 0.35 | >0.99 |
| C18:1 LPC | HMDB0002815* | Lysophosphatidylcholines | 1.15 (0.91-1.45) | 1.12 (0.86-1.45) | 1.29 (0.94-1.78) | 1.24 (0.88-1.74) | 0.22 | >0.99 | 1.00 (0.66-1.51) | 0.99 (0.65-1.50) | 0.97 | >0.99 |
| C18:1 LPC plasmalogen_A | HMDB0013122* | NA | 0.75 (0.43-1.30) | NA | NA | NA | NA | NA | NA | NA | NA | NA |
| C18:1 LPC plasmalogen_B | HMDB0013122* | NA | 0.64 (0.37-1.11) | NA | NA | NA | NA | NA | NA | NA | NA | NA |
| C18:1 LPE | HMDB0011506* | Lysophosphatidylethanolamines | 1.19 (0.97-1.47) | 1.07 (0.85-1.34) | 1.10 (0.84-1.45) | 1.06 (0.79-1.42) | 0.70 | >0.99 | 0.90 (0.63-1.29) | 0.90 (0.63-1.29) | 0.57 | >0.99 |
| C18:1 SM | HMDB0012101* | Sphingomyelins | 0.92 (0.73-1.15) | 0.93 (0.71-1.20) | 0.85 (0.61-1.19) | 0.81 (0.57-1.15) | 0.23 | >0.99 | 0.66 (0.41-1.06) | 0.66 (0.41-1.06) | 0.09 | >0.99 |
| C18:1-OH carnitine | HMDB0013339 | NA | 0.82 (0.67-1.01) | 0.84 (0.67-1.05) | 0.98 (0.75-1.28) | 0.97 (0.74-1.28) | 0.83 | >0.99 | 1.09 (0.78-1.52) | 1.09 (0.77-1.54) | 0.63 | >0.99 |
| C18:2 carnitine | HMDB0006469* | NA | 0.89 (0.54-1.48) | NA | NA | NA | NA | NA | NA | NA | NA | NA |
| C18:2 CE | HMDB0000610* | Cholesteryl esters | 0.94 (0.76-1.15) | 1.06 (0.83-1.35) | 1.19 (0.89-1.59) | 1.19 (0.89-1.59) | 0.25 | >0.99 | 1.19 (0.82-1.71) | 1.19 (0.82-1.71) | 0.36 | >0.99 |
| C18:2 LPC | HMDB0010386* | Lysophosphatidylcholines | 1.04 (0.85-1.28) | 0.99 (0.78-1.26) | 1.06 (0.79-1.43) | 1.02 (0.74-1.41) | 0.90 | >0.99 | 0.92 (0.62-1.36) | 0.92 (0.62-1.36) | 0.68 | >0.99 |
| C18:2 LPE | HMDB0011507* | Lysophosphatidylethanolamines | 1.16 (0.95-1.42) | 1.05 (0.83-1.32) | 1.08 (0.80-1.44) | 1.03 (0.75-1.41) | 0.85 | >0.99 | 0.88 (0.60-1.29) | 0.88 (0.60-1.29) | 0.51 | >0.99 |
| C18:3 CE | HMDB0010370* | Cholesteryl esters | 1.02 (0.84-1.23) | 1.10 (0.88-1.38) | 1.16 (0.89-1.49) | 1.13 (0.87-1.46) | 0.38 | >0.99 | 0.91 (0.66-1.26) | 0.91 (0.66-1.26) | 0.58 | >0.99 |
| C18:3 LPC | HMDB0010387* | Lysophosphatidylcholines | 1.16 (0.94-1.44) | 1.09 (0.86-1.38) | 1.20 (0.90-1.59) | 1.14 (0.84-1.56) | 0.40 | >0.99 | 1.00 (0.68-1.48) | 1.00 (0.68-1.48) | 0.99 | >0.99 |
| C18:3 LPE | HMDB0011478* | Lysophosphatidylethanolamines | 1.21 (0.98-1.49) | 1.15 (0.90-1.47) | 1.24 (0.91-1.70) | 1.22 (0.87-1.69) | 0.24 | >0.99 | 1.12 (0.74-1.70) | 1.12 (0.74-1.71) | 0.58 | >0.99 |
| C2 carnitine | HMDB0000201 | Carnitines | 0.87 (0.71-1.07) | 0.84 (0.67-1.05) | 0.88 (0.67-1.17) | 0.86 (0.64-1.14) | 0.29 | >0.99 | 0.95 (0.69-1.31) | 0.95 (0.69-1.31) | 0.74 | >0.99 |
| C20 carnitine | HMDB0006460 | NA | 1.04 (0.84-1.29) | 0.96 (0.75-1.23) | 1.12 (0.82-1.53) | 1.09 (0.79-1.49) | 0.61 | >0.99 | 1.16 (0.77-1.75) | 1.16 (0.77-1.75) | 0.47 | >0.99 |
| C20:0 LPE_A | HMDB0011481 | Lysophosphatidylethanolamines | 1.15 (0.92-1.43) | 1.11 (0.87-1.42) | 1.23 (0.91-1.65) | 1.26 (0.91-1.73) | 0.16 | >0.99 | 1.21 (0.82-1.78) | 1.21 (0.82-1.78) | 0.34 | >0.99 |
| C20:0 LPE_B | HMDB0011511 | Lysophosphatidylethanolamines | 1.19 (0.95-1.49) | 1.17 (0.91-1.51) | 1.32 (0.96-1.80) | 1.36 (0.97-1.90) | 0.07 | >0.99 | 1.49 (0.97-2.28) | 1.49 (0.97-2.28) | 0.07 | >0.99 |
| C20:0 SM | HMDB0012102 | Sphingomyelins | 1.05 (0.84-1.31) | 1.09 (0.85-1.39) | 1.18 (0.88-1.58) | 1.15 (0.85-1.56) | 0.36 | >0.99 | 1.09 (0.76-1.55) | 1.09 (0.76-1.55) | 0.64 | >0.99 |
| C20:1 LPC | HMDB0010391* | Lysophosphatidylcholines | 1.20 (0.95-1.50) | 1.15 (0.88-1.50) | 1.41 (1.01-1.96) | 1.44 (1.02-2.05) | 0.04 | >0.99 | 1.47 (0.97-2.23) | 1.47 (0.97-2.24) | 0.07 | >0.99 |
| C20:1 LPE | HMDB0011512* | Lysophosphatidylethanolamines | 1.32 (1.05-1.65) | 1.28 (1.00-1.63) | 1.46 (1.07-2.00) | 1.51 (1.08-2.11) | 0.02 | >0.99 | 1.51 (1.00-2.30) | 1.52 (1.00-2.31) | 0.05 | >0.99 |
| C20:3 CE | HMDB0006736* | Cholesteryl esters | 1.03 (0.84-1.26) | 1.16 (0.92-1.46) | 1.21 (0.91-1.61) | 1.20 (0.90-1.60) | 0.22 | >0.99 | 1.10 (0.78-1.55) | 1.10 (0.78-1.55) | 0.59 | >0.99 |
| C20:3 LPC | HMDB0010393* | Lysophosphatidylcholines | 1.19 (0.96-1.46) | 1.13 (0.89-1.43) | 1.22 (0.91-1.64) | 1.20 (0.89-1.62) | 0.23 | >0.99 | 1.26 (0.85-1.87) | 1.26 (0.85-1.86) | 0.24 | >0.99 |

| METABOLITE | HMDB_ID | Metabolite class | Model 1 | Model 2 | Model 3 | Model 4 | | | Model 5 | Model 6 | | |
|---|---|---|---|---|---|---|---|---|---|---|---|---|
| | | | OR (95%CI) | OR (95%CI) | OR (95%CI) | OR (95%CI) | p-value | NEF | OR (95%CI) | OR (95%CI) | p-value | NEF |
| C20:4 carnitine | HMDB0006455 | NA | 1.11 (0.89-1.38) | 1.13 (0.88-1.46) | 1.23 (0.90-1.69) | 1.23 (0.89-1.71) | 0.21 | >0.99 | 1.38 (0.94-2.03) | 1.38 (0.94-2.03) | 0.10 | >0.99 |
| C20:4 CE | HMDB0006726 | Cholesteryl esters | 0.92 (0.75-1.13) | 1.04 (0.82-1.32) | 1.08 (0.81-1.43) | 1.06 (0.79-1.42) | 0.72 | >0.99 | 1.01 (0.70-1.45) | 1.01 (0.70-1.45) | 0.96 | >0.99 |
| C20:4 LPC | HMDB0010395 | Lysophosphatidylcholines | 1.07 (0.86-1.33) | 1.10 (0.86-1.41) | 1.22 (0.89-1.66) | 1.17 (0.84-1.64) | 0.34 | >0.99 | 1.05 (0.67-1.64) | 1.04 (0.67-1.62) | 0.86 | >0.99 |
| C20:4 LPE | HMDB0011517 | Lysophosphatidylethanolamines | 1.18 (0.97-1.44) | 1.10 (0.88-1.37) | 1.16 (0.88-1.54) | 1.13 (0.84-1.52) | 0.43 | >0.99 | 0.97 (0.67-1.40) | 0.97 (0.67-1.41) | 0.87 | >0.99 |
| C20:5 CE | HMDB0006731 | Cholesteryl esters | 1.06 (0.86-1.30) | 1.17 (0.91-1.51) | 1.30 (0.96-1.76) | 1.25 (0.91-1.71) | 0.16 | >0.99 | 1.10 (0.74-1.63) | 1.10 (0.74-1.64) | 0.64 | >0.99 |
| C20:5 LPC | HMDB0010397 | Lysophosphatidylcholines | 1.22 (0.98-1.52) | 1.19 (0.92-1.53) | 1.28 (0.94-1.73) | 1.24 (0.91-1.70) | 0.17 | >0.99 | 1.18 (0.78-1.77) | 1.18 (0.78-1.78) | 0.43 | >0.99 |
| C22:0 Ceramide (d18:1) | HMDB0004952 | Ceramides | 0.98 (0.80-1.21) | 0.95 (0.75-1.20) | 0.98 (0.73-1.31) | 0.96 (0.70-1.31) | 0.79 | >0.99 | 0.91 (0.64-1.29) | 0.91 (0.64-1.29) | 0.60 | >0.99 |
| C22:0 LPE | HMDB0011520 | Lysophosphatidylethanolamines | 1.15 (0.93-1.43) | 1.12 (0.87-1.43) | 1.36 (1.00-1.85) | 1.42 (1.02-1.97) | 0.04 | >0.99 | 1.28 (0.85-1.93) | 1.28 (0.85-1.93) | 0.24 | >0.99 |
| C22:0 SM | HMDB0012103 | Sphingomyelins | 0.83 (0.48-1.44) | NA | NA | NA | NA | NA | NA | NA | NA | NA |
| C22:4 CE | HMDB0006729* | Cholesteryl esters | 0.86 (0.70-1.06) | 0.95 (0.75-1.21) | 1.05 (0.78-1.42) | 1.08 (0.79-1.47) | 0.63 | >0.99 | 0.99 (0.67-1.46) | 0.99 (0.67-1.46) | 0.96 | >0.99 |
| C22:4 LPC | HMDB0010401* | Lysophosphatidylcholines | 1.03 (0.83-1.26) | 1.03 (0.81-1.31) | 1.12 (0.83-1.51) | 1.10 (0.81-1.48) | 0.56 | >0.99 | 1.08 (0.73-1.61) | 1.08 (0.73-1.61) | 0.69 | >0.99 |
| C22:5 CE | HMDB0010375* | Cholesteryl esters | 0.91 (0.74-1.12) | 1.02 (0.80-1.29) | 1.15 (0.86-1.54) | 1.18 (0.87-1.59) | 0.29 | >0.99 | 1.09 (0.76-1.56) | 1.09 (0.76-1.56) | 0.64 | >0.99 |
| C22:5 LPC | HMDB0010403* | Lysophosphatidylcholines | 1.09 (0.88-1.35) | 1.10 (0.86-1.40) | 1.27 (0.92-1.75) | 1.26 (0.90-1.76) | 0.18 | >0.99 | 1.21 (0.79-1.86) | 1.21 (0.79-1.86) | 0.37 | >0.99 |
| C22:6 CE | HMDB0006733 | Cholesteryl esters | 0.96 (0.78-1.18) | 1.01 (0.79-1.29) | 1.04 (0.78-1.39) | 1.01 (0.75-1.35) | 0.95 | >0.99 | 1.00 (0.70-1.44) | 1.00 (0.70-1.44) | 0.98 | >0.99 |
| C22:6 LPC | HMDB0010404 | Lysophosphatidylcholines | 1.15 (0.92-1.43) | 1.12 (0.87-1.44) | 1.17 (0.87-1.59) | 1.09 (0.78-1.50) | 0.62 | >0.99 | 0.98 (0.63-1.52) | 0.97 (0.62-1.51) | 0.89 | >0.99 |
| C22:6 LPE | HMDB0011526 | Lysophosphatidylethanolamines | 1.21 (0.98-1.49) | 1.16 (0.91-1.48) | 1.29 (0.96-1.74) | 1.26 (0.92-1.71) | 0.15 | >0.99 | 1.30 (0.88-1.93) | 1.31 (0.88-1.95) | 0.18 | >0.99 |
| C24:0 Ceramide (d18:1) | HMDB0004956 | Ceramides | 1.02 (0.83-1.24) | 0.99 (0.79-1.24) | 1.02 (0.77-1.34) | 0.98 (0.73-1.32) | 0.90 | >0.99 | 0.91 (0.65-1.29) | 0.91 (0.65-1.29) | 0.61 | >0.99 |
| C24:0 PC | NA | Phosphatidylcholines | 1.01 (0.79-1.29) | 0.91 (0.69-1.20) | 1.00 (0.72-1.39) | 0.94 (0.66-1.32) | 0.71 | >0.99 | 0.85 (0.57-1.28) | 0.85 (0.57-1.28) | 0.44 | >0.99 |
| C24:1 Ceramide (d18:1) | HMDB0004953* | Ceramides | 1.11 (0.89-1.38) | 1.02 (0.79-1.31) | 1.14 (0.84-1.55) | 1.13 (0.82-1.54) | 0.46 | >0.99 | 1.04 (0.71-1.51) | 1.04 (0.71-1.51) | 0.85 | >0.99 |
| C24:1 SM | HMDB0012107* | Sphingomyelins | 1.12 (0.90-1.38) | 1.11 (0.87-1.41) | 1.21 (0.91-1.61) | 1.18 (0.88-1.59) | 0.27 | >0.99 | 1.11 (0.76-1.63) | 1.11 (0.76-1.64) | 0.58 | >0.99 |
| C26 carnitine | HMDB0006347 | Carnitines | 1.25 (1.00-1.57) | 1.19 (0.92-1.54) | 1.37 (0.99-1.90) | 1.34 (0.96-1.88) | 0.09 | >0.99 | 1.57 (1.03-2.40) | 1.59 (1.04-2.43) | 0.03 | >0.99 |
| C3 carnitine | HMDB0000824 | Carnitines | 0.97 (0.79-1.19) | 0.93 (0.73-1.17) | 0.76 (0.57-1.02) | 0.75 (0.56-1.01) | 0.06 | >0.99 | 0.83 (0.58-1.19) | 0.83 (0.58-1.19) | 0.32 | >0.99 |
| C30:0 DAG | HMDB0007011* | Diglycerides | 0.88 (0.68-1.12) | 0.81 (0.61-1.09) | 0.73 (0.49-1.08) | 0.70 (0.47-1.06) | 0.09 | >0.99 | 0.67 (0.41-1.10) | NA | NA | NA |
| C30:0 PC | HMDB0007869* | Phosphatidylcholines | 1.09 (0.88-1.35) | 1.08 (0.84-1.38) | 1.10 (0.82-1.47) | 1.05 (0.78-1.42) | 0.75 | >0.99 | 0.93 (0.64-1.36) | 0.93 (0.64-1.37) | 0.72 | >0.99 |
| C30:1 PC | HMDB0007870* | Phosphatidylcholines | 1.09 (0.88-1.35) | 1.09 (0.85-1.39) | 1.07 (0.80-1.42) | 1.03 (0.77-1.39) | 0.82 | >0.99 | 0.92 (0.64-1.33) | 0.92 (0.64-1.33) | 0.67 | >0.99 |
| C32:0 DAG | HMDB0007098* | Diglycerides | 1.14 (0.93-1.41) | 1.05 (0.83-1.34) | 0.98 (0.74-1.31) | 0.95 (0.71-1.28) | 0.74 | >0.99 | 0.92 (0.64-1.32) | 0.92 (0.64-1.32) | 0.64 | >0.99 |
| C32:0 PC | HMDB0007871* | Phosphatidylcholines | 1.13 (0.92-1.39) | 1.06 (0.84-1.33) | 1.17 (0.89-1.54) | 1.11 (0.83-1.50) | 0.48 | >0.99 | 0.90 (0.62-1.32) | 0.90 (0.62-1.32) | 0.60 | >0.99 |
| C32:0 PE | HMDB0008923* | Phosphatidylethanolamines | 1.15 (0.91-1.44) | 1.18 (0.90-1.53) | 1.13 (0.83-1.55) | 1.16 (0.84-1.60) | 0.36 | >0.99 | 1.21 (0.79-1.86) | 1.21 (0.79-1.87) | 0.37 | >0.99 |
| C32:1 DAG | HMDB0007099* | Diglycerides | 1.14 (0.92-1.41) | 1.06 (0.84-1.35) | 0.97 (0.72-1.30) | 0.96 (0.71-1.30) | 0.80 | >0.99 | 0.99 (0.69-1.41) | 0.98 (0.68-1.41) | 0.93 | >0.99 |
| C32:1 PC | HMDB0007873* | Phosphatidylcholines | 1.13 (0.91-1.39) | 1.11 (0.87-1.41) | 1.13 (0.86-1.49) | 1.08 (0.80-1.44) | 0.61 | >0.99 | 0.92 (0.65-1.31) | 0.92 (0.65-1.32) | 0.65 | >0.99 |
| C32:2 PC | HMDB0007874* | Phosphatidylcholines | 1.00 (0.81-1.23) | 1.03 (0.81-1.32) | 1.05 (0.78-1.40) | 1.04 (0.77-1.40) | 0.81 | >0.99 | 1.03 (0.71-1.51) | 1.03 (0.70-1.51) | 0.87 | >0.99 |

| METABOLITE | HMDB_ID | Metabolite class | Model 1 | Model 2 | Model 3 | Model 4 | | | Model 5 | Model 6 | | |
|---|---|---|---|---|---|---|---|---|---|---|---|---|
| | | | OR (95%CI) | OR (95%CI) | OR (95%CI) | OR (95%CI) | p-value | NEF | OR (95%CI) | OR (95%CI) | p-value | NEF |
| C34:0 DAG | HMDB0007100* | Diglycerides | 1.06 (0.86-1.31) | 0.95 (0.75-1.20) | 0.89 (0.67-1.18) | 0.85 (0.64-1.14) | 0.28 | >0.99 | 0.80 (0.56-1.14) | 0.79 (0.55-1.14) | 0.21 | >0.99 |
| C34:0 PC | HMDB0007970* | Phosphatidylcholines | 0.68 (0.38-1.21) | NA | NA | NA | NA | NA | NA | NA | NA | NA |
| C34:0 PE | HMDB0008925* | Phosphatidylethanolamines | 1.14 (0.92-1.40) | 1.12 (0.88-1.42) | 1.13 (0.86-1.50) | 1.14 (0.86-1.51) | 0.38 | >0.99 | 1.04 (0.74-1.45) | 1.04 (0.74-1.46) | 0.81 | >0.99 |
| C34:0 PS | HMDB0012356* | Phosphatidylserines | 1.18 (0.95-1.46) | 1.20 (0.93-1.55) | 1.35 (0.99-1.85) | 1.30 (0.94-1.80) | 0.11 | >0.99 | 1.21 (0.79-1.87) | 1.22 (0.79-1.88) | 0.38 | >0.99 |
| C34:1 DAG | HMDB0007102* | Diglycerides | 1.10 (0.90-1.36) | 1.03 (0.81-1.30) | 0.96 (0.72-1.28) | 0.95 (0.71-1.26) | 0.70 | >0.99 | 0.97 (0.68-1.39) | 0.97 (0.67-1.40) | 0.87 | >0.99 |
| C34:1 PC | HMDB0007972* | Phosphatidylcholines | 1.18 (0.95-1.45) | 1.13 (0.89-1.43) | 1.18 (0.90-1.55) | 1.12 (0.84-1.50) | 0.45 | >0.99 | 0.90 (0.63-1.28) | 0.90 (0.63-1.28) | 0.56 | >0.99 |
| C34:1 PC plasmalogen-A | HMDB0011208* | Phosphatidylcholine plasmalogens | 1.07 (0.88-1.32) | 0.96 (0.76-1.22) | 1.17 (0.89-1.55) | 1.17 (0.88-1.56) | 0.29 | >0.99 | 1.05 (0.73-1.49) | 1.05 (0.73-1.49) | 0.81 | >0.99 |
| C34:1 PC plasmalogen-B | HMDB0011239* | Phosphatidylcholine plasmalogens | 1.02 (0.84-1.25) | 0.99 (0.78-1.24) | 1.09 (0.83-1.43) | 1.07 (0.80-1.42) | 0.66 | >0.99 | 0.96 (0.67-1.37) | 0.96 (0.67-1.38) | 0.83 | >0.99 |
| C34:2 DAG | HMDB0007103* | Diglycerides | 1.09 (0.88-1.35) | 1.00 (0.79-1.28) | 0.92 (0.68-1.25) | 0.92 (0.68-1.25) | 0.60 | >0.99 | 0.95 (0.66-1.38) | 0.95 (0.65-1.38) | 0.79 | >0.99 |
| C34:2 PC | HMDB0007973* | Phosphatidylcholines | 1.11 (0.88-1.40) | 1.08 (0.83-1.41) | 1.10 (0.81-1.50) | 1.07 (0.76-1.50) | 0.71 | >0.99 | 0.96 (0.61-1.52) | 0.96 (0.61-1.52) | 0.88 | >0.99 |
| C34:2 PC plasmalogen | HMDB0011210* | Phosphatidylcholine plasmalogens | 1.14 (0.93-1.38) | 1.12 (0.89-1.41) | 1.30 (0.98-1.72) | 1.29 (0.95-1.75) | 0.10 | >0.99 | 1.12 (0.76-1.65) | 1.12 (0.76-1.66) | 0.55 | >0.99 |
| C34:2 PE | HMDB0008928* | Phosphatidylethanolamines | 1.18 (0.96-1.46) | 1.09 (0.86-1.39) | 1.07 (0.80-1.43) | 1.04 (0.78-1.40) | 0.77 | >0.99 | 0.99 (0.70-1.40) | 0.99 (0.70-1.40) | 0.95 | >0.99 |
| C34:2 PE plasmalogen | HMDB0008952* | Phosphatidylethanolamine plasmalogens | 1.09 (0.88-1.34) | 1.12 (0.89-1.41) | 1.39 (1.03-1.88) | 1.37 (1.00-1.87) | 0.05 | >0.99 | 1.35 (0.95-1.93) | 1.37 (0.95-1.96) | 0.09 | >0.99 |
| C34:3 DAG | HMDB0007132* | Diglycerides | 1.06 (0.86-1.32) | 0.97 (0.76-1.24) | 0.93 (0.68-1.28) | 0.97 (0.70-1.34) | 0.84 | >0.99 | 1.05 (0.71-1.54) | 1.05 (0.71-1.54) | 0.82 | >0.99 |
| C34:3 PC | HMDB0008006* | Phosphatidylcholines | 1.03 (0.83-1.27) | 1.03 (0.81-1.31) | 1.06 (0.80-1.42) | 1.05 (0.78-1.41) | 0.74 | >0.99 | 0.90 (0.61-1.33) | 0.90 (0.61-1.33) | 0.60 | >0.99 |
| C34:3 PC plasmalogen | HMDB0011211* | Phosphatidylcholine plasmalogens | 1.11 (0.92-1.34) | 1.13 (0.91-1.41) | 1.35 (1.02-1.78) | 1.37 (1.02-1.84) | 0.03 | >0.99 | 1.39 (0.95-2.02) | 1.41 (0.96-2.08) | 0.08 | >0.99 |
| C34:3 PE plasmalogen | HMDB0011343* | Phosphatidylethanolamine plasmalogens | 1.01 (0.83-1.24) | 1.04 (0.83-1.30) | 1.19 (0.90-1.59) | 1.18 (0.89-1.58) | 0.25 | >0.99 | 1.29 (0.91-1.82) | 1.30 (0.92-1.84) | 0.14 | >0.99 |
| C34:4 PC | HMDB0007883* | Phosphatidylcholines | 0.99 (0.80-1.23) | 1.03 (0.80-1.31) | 1.03 (0.77-1.38) | 1.02 (0.75-1.37) | 0.92 | >0.99 | 0.96 (0.65-1.41) | 0.96 (0.65-1.42) | 0.84 | >0.99 |
| C34:4 PC plasmalogen | HMDB0011212* | Phosphatidylcholine plasmalogens | 0.95 (0.78-1.16) | 0.95 (0.76-1.20) | 1.01 (0.77-1.33) | 0.99 (0.74-1.32) | 0.94 | >0.99 | 0.91 (0.64-1.30) | 0.91 (0.64-1.31) | 0.62 | >0.99 |
| C34:5 PC plasmalogen | HMDB0011214* | Phosphatidylcholine plasmalogens | 0.92 (0.75-1.12) | 0.95 (0.76-1.18) | 1.08 (0.82-1.42) | 1.07 (0.80-1.42) | 0.66 | >0.99 | 1.11 (0.79-1.56) | 1.12 (0.79-1.58) | 0.52 | >0.99 |
| C36:0 DAG | HMDB0007158* | Diglycerides | 0.82 (0.45-1.48) | NA | NA | NA | NA | NA | NA | NA | NA | NA |
| C36:0 PC | HMDB0008036* | Phosphatidylcholines | 0.73 (0.39-1.34) | NA | NA | NA | NA | NA | NA | NA | NA | NA |
| C36:0 PE | HMDB0008991* | Phosphatidylethanolamines | 1.20 (0.96-1.51) | 1.18 (0.91-1.52) | 1.20 (0.89-1.61) | 1.19 (0.87-1.62) | 0.28 | >0.99 | 1.11 (0.74-1.67) | 1.10 (0.73-1.66) | 0.63 | >0.99 |
| C36:1 DAG | HMDB0007216* | Diglycerides | 1.08 (0.88-1.33) | 0.99 (0.78-1.25) | 0.91 (0.68-1.21) | 0.88 (0.66-1.18) | 0.39 | >0.99 | 0.91 (0.63-1.30) | 0.90 (0.62-1.30) | 0.57 | >0.99 |
| C36:1 PC | HMDB0008038* | Phosphatidylcholines | 1.09 (0.88-1.35) | 1.03 (0.81-1.31) | 1.15 (0.86-1.54) | 1.11 (0.82-1.50) | 0.50 | >0.99 | 0.89 (0.62-1.29) | 0.89 (0.62-1.29) | 0.54 | >0.99 |
| C36:1 PC plasmalogen | HMDB0011241* | Phosphatidylcholine plasmalogens | 0.74 (0.41-1.33) | NA | NA | NA | NA | NA | NA | NA | NA | NA |
| C36:1 PE | HMDB0008993* | Phosphatidylethanolamines | 1.15 (0.92-1.45) | 1.09 (0.84-1.42) | 1.10 (0.81-1.49) | 1.09 (0.79-1.51) | 0.59 | >0.99 | 1.05 (0.68-1.61) | 1.06 (0.69-1.63) | 0.80 | >0.99 |
| C36:1 PE plasmalogen | HMDB0009016* | Phosphatidylethanolamine plasmalogens | 1.05 (0.85-1.29) | 1.04 (0.82-1.31) | 1.15 (0.87-1.51) | 1.14 (0.86-1.52) | 0.36 | >0.99 | 1.06 (0.76-1.48) | 1.06 (0.76-1.49) | 0.72 | >0.99 |
| C36:2 DAG | HMDB0007218* | Diglycerides | 1.09 (0.89-1.34) | 1.00 (0.79-1.26) | 0.94 (0.71-1.26) | 0.93 (0.70-1.25) | 0.64 | >0.99 | 0.98 (0.69-1.40) | 0.98 (0.69-1.40) | 0.92 | >0.99 |
| C36:2 PC | HMDB0008039* | Phosphatidylcholines | 1.04 (0.83-1.29) | 0.96 (0.75-1.23) | 1.08 (0.79-1.47) | 1.06 (0.77-1.45) | 0.74 | >0.99 | 1.00 (0.66-1.53) | 1.00 (0.66-1.53) | 0.98 | >0.99 |
| C36:2 PC plasmalogen | HMDB0011243* | Phosphatidylcholine plasmalogens | 0.99 (0.81-1.22) | 0.94 (0.74-1.19) | 1.10 (0.83-1.46) | 1.12 (0.83-1.50) | 0.46 | >0.99 | 1.03 (0.70-1.51) | 1.03 (0.70-1.51) | 0.89 | >0.99 |

| METABOLITE | HMDB_ID | Metabolite class | Model 1 | Model 2 | Model 3 | Model 4 | | | Model 5 | Model 6 | | |
|---|---|---|---|---|---|---|---|---|---|---|---|---|
| | | | OR (95%CI) | OR (95%CI) | OR (95%CI) | OR (95%CI) | p-value | NEF | OR (95%CI) | OR (95%CI) | p-value | NEF |
| C36:2 PE | HMDB0008994* | Phosphatidylethanolamines | 1.14 (0.93-1.41) | 1.03 (0.81-1.30) | 0.99 (0.74-1.32) | 0.96 (0.71-1.28) | 0.77 | >0.99 | 0.99 (0.69-1.42) | 0.99 (0.69-1.42) | 0.95 | >0.99 |
| C36:2 PE plasmalogen | HMDB0009082* | Phosphatidylethanolamine plasmalogens | 0.99 (0.81-1.21) | 1.01 (0.80-1.27) | 1.18 (0.89-1.58) | 1.15 (0.86-1.55) | 0.34 | >0.99 | 1.15 (0.81-1.64) | 1.16 (0.81-1.65) | 0.42 | >0.99 |
| C36:2 PS plasmalogen | NA | Phosphatidylserine plasmalogens | 0.94 (0.75-1.17) | 0.97 (0.76-1.24) | 1.00 (0.74-1.35) | 0.99 (0.72-1.35) | 0.93 | >0.99 | 1.04 (0.70-1.55) | 1.04 (0.70-1.55) | 0.84 | >0.99 |
| C36:3 DAG | HMDB0007219* | Diglycerides | 0.99 (0.80-1.22) | 0.89 (0.69-1.14) | 0.85 (0.63-1.16) | 0.87 (0.64-1.19) | 0.39 | >0.99 | 0.95 (0.64-1.39) | 0.94 (0.64-1.39) | 0.77 | >0.99 |
| C36:3 PC | HMDB0008105* | Phosphatidylcholines | 0.99 (0.81-1.23) | 0.97 (0.76-1.22) | 1.01 (0.76-1.34) | 1.00 (0.74-1.34) | 0.97 | >0.99 | 0.86 (0.59-1.25) | 0.85 (0.59-1.24) | 0.41 | >0.99 |
| C36:3 PC plasmalogen | HMDB0011244* | Phosphatidylcholine plasmalogens | 0.99 (0.82-1.21) | 1.01 (0.80-1.27) | 1.17 (0.89-1.54) | 1.18 (0.88-1.59) | 0.26 | >0.99 | 1.13 (0.77-1.65) | 1.13 (0.77-1.67) | 0.52 | >0.99 |
| C36:3 PE | HMDB0009060* | Phosphatidylethanolamines | 1.06 (0.87-1.30) | 0.94 (0.74-1.20) | 0.92 (0.68-1.23) | 0.90 (0.67-1.21) | 0.48 | >0.99 | 0.85 (0.59-1.21) | 0.85 (0.59-1.21) | 0.37 | >0.99 |
| C36:3 PE plasmalogen | HMDB0011441* | Phosphatidylethanolamine plasmalogens | 0.96 (0.79-1.18) | 0.98 (0.78-1.24) | 1.14 (0.85-1.52) | 1.11 (0.83-1.50) | 0.48 | >0.99 | 1.11 (0.78-1.58) | 1.11 (0.78-1.59) | 0.56 | >0.99 |
| C36:3 PS plasmalogen | NA | Phosphatidylserine plasmalogens | 1.09 (0.89-1.33) | 1.10 (0.88-1.38) | 1.11 (0.84-1.46) | 1.07 (0.81-1.43) | 0.63 | >0.99 | 1.13 (0.79-1.62) | 1.13 (0.79-1.62) | 0.49 | >0.99 |
| C36:4 DAG | HMDB0007248* | Diglycerides | 0.91 (0.74-1.13) | 0.83 (0.64-1.06) | 0.80 (0.59-1.10) | 0.84 (0.60-1.17) | 0.31 | >0.99 | 1.02 (0.69-1.52) | 1.02 (0.68-1.52) | 0.92 | >0.99 |
| C36:4 PC plasmalogen | HMDB0011310* | Phosphatidylcholine plasmalogens | 0.98 (0.79-1.20) | 0.97 (0.77-1.22) | 1.16 (0.88-1.53) | 1.17 (0.88-1.56) | 0.28 | >0.99 | 1.15 (0.82-1.63) | 1.16 (0.82-1.64) | 0.41 | >0.99 |
| C36:4 PC-A | HMDB0007983* | Phosphatidylcholines | 0.99 (0.80-1.21) | 0.99 (0.78-1.26) | 1.15 (0.85-1.57) | 1.13 (0.83-1.54) | 0.45 | >0.99 | 0.96 (0.64-1.44) | 0.96 (0.64-1.44) | 0.84 | >0.99 |
| C36:4 PC-B | HMDB0008138* | Phosphatidylcholines | 1.01 (0.82-1.25) | 1.03 (0.81-1.31) | 1.04 (0.78-1.38) | 1.00 (0.74-1.34) | 0.99 | >0.99 | 0.93 (0.64-1.36) | 0.93 (0.64-1.36) | 0.71 | >0.99 |
| C36:4 PE | HMDB0008937* | Phosphatidylethanolamines | 1.20 (0.97-1.47) | 1.13 (0.90-1.42) | 1.15 (0.88-1.51) | 1.11 (0.84-1.47) | 0.45 | >0.99 | 1.05 (0.75-1.46) | 1.05 (0.75-1.46) | 0.78 | >0.99 |
| C36:4 PE plasmalogen | HMDB0011442* | Phosphatidylethanolamine plasmalogens | 1.00 (0.82-1.22) | 1.02 (0.81-1.27) | 1.13 (0.86-1.50) | 1.13 (0.85-1.51) | 0.39 | >0.99 | 1.19 (0.85-1.68) | 1.20 (0.85-1.70) | 0.29 | >0.99 |
| C36:4 PI | HMDB0009789* | NA | 0.99 (0.55-1.78) | NA | NA | NA | NA | NA | NA | NA | NA | NA |
| C36:5 PC plasmalogen-B | HMDB0011220* | Phosphatidylcholine plasmalogens | 0.95 (0.78-1.17) | 0.97 (0.77-1.23) | 1.14 (0.86-1.50) | 1.12 (0.84-1.50) | 0.43 | >0.99 | 1.14 (0.82-1.59) | 1.15 (0.82-1.63) | 0.42 | >0.99 |
| C36:5 PE plasmalogen | HMDB0011410* | Phosphatidylethanolamine plasmalogens | 0.99 (0.81-1.21) | 0.99 (0.79-1.24) | 1.11 (0.85-1.46) | 1.09 (0.82-1.44) | 0.55 | >0.99 | 1.21 (0.87-1.70) | 1.23 (0.87-1.75) | 0.24 | >0.99 |
| C38:2 PC | HMDB0008270* | Phosphatidylcholines | 1.07 (0.86-1.32) | 1.00 (0.78-1.26) | 1.18 (0.87-1.59) | 1.17 (0.87-1.59) | 0.30 | >0.99 | 1.09 (0.75-1.60) | 1.09 (0.75-1.60) | 0.64 | >0.99 |
| C38:2 PE | HMDB0008942* | Phosphatidylethanolamines | 1.11 (0.92-1.35) | 1.10 (0.87-1.38) | 1.19 (0.89-1.58) | 1.22 (0.90-1.64) | 0.20 | >0.99 | 1.23 (0.86-1.76) | 1.24 (0.87-1.77) | 0.24 | >0.99 |
| C38:3 PC | HMDB0008047* | Phosphatidylcholines | 1.08 (0.88-1.33) | 1.02 (0.81-1.29) | 1.04 (0.79-1.38) | 1.04 (0.78-1.38) | 0.78 | >0.99 | 1.00 (0.71-1.43) | 1.00 (0.70-1.43) | 0.99 | >0.99 |
| C38:3 PE plasmalogen | HMDB0011384* | Phosphatidylethanolamine plasmalogens | 1.03 (0.85-1.25) | 1.08 (0.86-1.35) | 1.25 (0.94-1.64) | 1.29 (0.96-1.74) | 0.09 | >0.99 | 1.24 (0.87-1.78) | 1.26 (0.87-1.81) | 0.22 | >0.99 |
| C38:4 DAG | HMDB0007170* | Diglycerides | 0.90 (0.45-1.77) | NA | NA | NA | NA | NA | NA | NA | NA | NA |
| C38:4 PC | HMDB0008048* | Phosphatidylcholines | 0.92 (0.75-1.13) | 0.92 (0.73-1.17) | 0.93 (0.71-1.23) | 0.93 (0.70-1.23) | 0.61 | >0.99 | 0.91 (0.63-1.32) | 0.91 (0.63-1.32) | 0.61 | >0.99 |
| C38:4 PC plasmalogen | HMDB0011252* | Phosphatidylcholine plasmalogens | 0.97 (0.78-1.19) | 0.91 (0.71-1.16) | 1.03 (0.78-1.37) | 1.03 (0.77-1.38) | 0.83 | >0.99 | 0.98 (0.69-1.38) | 0.98 (0.69-1.38) | 0.90 | >0.99 |
| C38:4 PE | HMDB0009003* | Phosphatidylethanolamines | 1.12 (0.91-1.37) | 1.06 (0.84-1.33) | 1.04 (0.79-1.37) | 1.01 (0.77-1.33) | 0.94 | >0.99 | 1.04 (0.74-1.47) | 1.04 (0.74-1.47) | 0.80 | >0.99 |
| C38:5 DAG | HMDB0007199* | Diglycerides | 1.01 (0.81-1.24) | 0.95 (0.75-1.21) | 0.90 (0.67-1.22) | 0.88 (0.65-1.19) | 0.39 | >0.99 | 0.81 (0.55-1.19) | 0.81 (0.55-1.19) | 0.29 | >0.99 |
| C38:5 PE | HMDB0009069* | Phosphatidylethanolamines | 1.08 (0.88-1.33) | 1.01 (0.80-1.28) | 1.07 (0.80-1.43) | 1.06 (0.79-1.42) | 0.72 | >0.99 | 0.97 (0.69-1.37) | 0.97 (0.69-1.37) | 0.88 | >0.99 |
| C38:5 PE plasmalogen | HMDB0011386* | Phosphatidylethanolamine plasmalogens | 0.90 (0.73-1.11) | 0.91 (0.72-1.15) | 1.06 (0.80-1.41) | 1.04 (0.77-1.40) | 0.78 | >0.99 | 1.13 (0.79-1.62) | 1.14 (0.79-1.65) | 0.48 | >0.99 |
| C38:6 PC | HMDB0007991* | Phosphatidylcholines | 1.10 (0.89-1.37) | 1.05 (0.82-1.35) | 1.13 (0.84-1.52) | 1.08 (0.79-1.48) | 0.64 | >0.99 | 1.03 (0.68-1.56) | 1.03 (0.68-1.56) | 0.89 | >0.99 |
| C38:6 PC plasmalogen | HMDB0011319* | Phosphatidylcholine plasmalogens | 0.77 (0.43-1.36) | NA | NA | NA | NA | NA | NA | NA | NA | NA |

| METABOLITE | HMDB_ID | Metabolite class | Model 1 | Model 2 | Model 3 | Model 4 | | | Model 5 | Model 6 | | |
|---|---|---|---|---|---|---|---|---|---|---|---|---|
| | | | OR (95%CI) | OR (95%CI) | OR (95%CI) | OR (95%CI) | p-value | NEF | OR (95%CI) | OR (95%CI) | p-value | NEF |
| C38:6 PE | HMDB0009102* | Phosphatidylethanolamines | 1.15 (0.94-1.42) | 1.10 (0.87-1.39) | 1.16 (0.87-1.54) | 1.12 (0.84-1.50) | 0.44 | >0.99 | 1.17 (0.81-1.69) | 1.18 (0.81-1.70) | 0.39 | >0.99 |
| C38:6 PE plasmalogen | HMDB0011387* | Phosphatidylethanolamine plasmalogens | 1.01 (0.83-1.23) | 1.02 (0.82-1.28) | 1.20 (0.91-1.58) | 1.20 (0.90-1.59) | 0.21 | >0.99 | 1.25 (0.89-1.75) | 1.27 (0.89-1.80) | 0.18 | >0.99 |
| C38:6 PS | HMDB0012362* | Phosphatidylserines | 1.16 (0.94-1.42) | 1.15 (0.91-1.46) | 1.12 (0.85-1.48) | 1.11 (0.84-1.48) | 0.46 | >0.99 | 0.99 (0.70-1.39) | 0.99 (0.70-1.39) | 0.94 | >0.99 |
| C38:7 PC plasmalogen | HMDB0011229* | Phosphatidylcholine plasmalogens | 1.07 (0.87-1.31) | 1.04 (0.82-1.32) | 1.22 (0.91-1.64) | 1.19 (0.87-1.61) | 0.28 | >0.99 | 1.27 (0.86-1.88) | 1.30 (0.86-1.95) | 0.21 | >0.99 |
| C38:7 PE plasmalogen | HMDB0011420* | Phosphatidylethanolamine plasmalogens | 1.11 (0.91-1.35) | 1.09 (0.87-1.37) | 1.25 (0.95-1.65) | 1.22 (0.91-1.63) | 0.19 | >0.99 | 1.36 (0.95-1.95) | 1.40 (0.96-2.04) | 0.08 | >0.99 |
| C3-DC-CH3 carnitine | HMDB0013133 | NA | 0.79 (0.45-1.41) | NA | NA | NA | NA | NA | NA | NA | NA | NA |
| C4 carnitine | HMDB0002013 | Carnitines | 0.93 (0.76-1.14) | 0.96 (0.77-1.21) | 0.84 (0.64-1.11) | 0.85 (0.65-1.12) | 0.26 | >0.99 | 0.89 (0.63-1.26) | 0.89 (0.63-1.26) | 0.51 | >0.99 |
| C40:10 PC | HMDB0008511* | Phosphatidylcholines | 1.15 (0.93-1.42) | 1.10 (0.86-1.42) | 1.21 (0.89-1.64) | 1.18 (0.86-1.62) | 0.30 | >0.99 | 1.06 (0.70-1.61) | 1.06 (0.70-1.62) | 0.77 | >0.99 |
| C40:6 PC | HMDB0008057* | Phosphatidylcholines | 1.02 (0.83-1.26) | 0.96 (0.76-1.22) | 1.01 (0.75-1.35) | 0.99 (0.73-1.33) | 0.94 | >0.99 | 0.95 (0.64-1.41) | 0.95 (0.64-1.41) | 0.81 | >0.99 |
| C40:6 PE | HMDB0009012* | Phosphatidylethanolamines | 1.11 (0.91-1.37) | 1.06 (0.84-1.34) | 1.03 (0.78-1.37) | 1.01 (0.75-1.35) | 0.96 | >0.99 | 1.15 (0.78-1.70) | 1.15 (0.78-1.70) | 0.47 | >0.99 |
| C40:7 PC plasmalogen | HMDB0011294* | Phosphatidylcholine plasmalogens | 0.68 (0.35-1.31) | NA | NA | NA | NA | NA | NA | NA | NA | NA |
| C40:7 PE plasmalogen | HMDB0011394* | Phosphatidylethanolamine plasmalogens | 0.99 (0.82-1.21) | 0.98 (0.78-1.23) | 1.14 (0.86-1.50) | 1.11 (0.83-1.49) | 0.47 | >0.99 | 1.27 (0.88-1.81) | 1.28 (0.89-1.84) | 0.19 | >0.99 |
| C40:9 PC | HMDB0008731* | Phosphatidylcholines | 1.09 (0.88-1.35) | 1.04 (0.81-1.34) | 1.11 (0.83-1.50) | 1.07 (0.78-1.46) | 0.69 | >0.99 | 1.02 (0.68-1.54) | 1.02 (0.68-1.54) | 0.91 | >0.99 |
| C41:0 TAG | NA | Triglycerides | 1.11 (0.89-1.39) | 0.89 (0.69-1.16) | 0.73 (0.52-1.02) | 0.73 (0.52-1.02) | 0.07 | >0.99 | 0.70 (0.46-1.06) | 0.70 (0.46-1.06) | 0.09 | >0.99 |
| C42:0 TAG | HMDB0072780* | Triglycerides | 1.12 (0.90-1.40) | 0.96 (0.75-1.24) | 0.82 (0.59-1.13) | 0.80 (0.58-1.11) | 0.18 | >0.99 | 0.76 (0.50-1.16) | 0.76 (0.49-1.16) | 0.20 | >0.99 |
| C43:0 TAG | HMDB0042062* | Triglycerides | 1.07 (0.86-1.34) | 0.87 (0.67-1.14) | 0.70 (0.50-1.00) | 0.70 (0.49-0.99) | 0.05 | >0.99 | 0.71 (0.46-1.09) | 0.70 (0.46-1.08) | 0.11 | >0.99 |
| C43:1 TAG | HMDB0042098* | Triglycerides | 1.10 (0.88-1.37) | 0.94 (0.72-1.22) | 0.79 (0.56-1.11) | 0.78 (0.55-1.10) | 0.15 | >0.99 | 0.76 (0.50-1.17) | 0.76 (0.50-1.17) | 0.21 | >0.99 |
| C43:2 TAG | HMDB0043169* | Triglycerides | 1.10 (0.88-1.37) | 0.95 (0.73-1.23) | 0.83 (0.59-1.15) | 0.80 (0.57-1.12) | 0.20 | >0.99 | 0.80 (0.53-1.22) | 0.80 (0.53-1.22) | 0.30 | >0.99 |
| C44:0 TAG | HMDB0042063* | Triglycerides | 1.13 (0.91-1.40) | 1.00 (0.77-1.28) | 0.87 (0.63-1.19) | 0.84 (0.61-1.16) | 0.30 | >0.99 | 0.77 (0.51-1.16) | 0.77 (0.50-1.16) | 0.21 | >0.99 |
| C44:1 TAG | HMDB0042301* | Triglycerides | 1.12 (0.90-1.39) | 1.00 (0.78-1.28) | 0.89 (0.65-1.21) | 0.87 (0.64-1.19) | 0.38 | >0.99 | 0.84 (0.56-1.25) | 0.84 (0.56-1.25) | 0.38 | >0.99 |
| C44:13 PE plasmalogen | NA | Phosphatidylethanolamine plasmalogens | 0.94 (0.76-1.15) | 0.97 (0.77-1.23) | 1.14 (0.84-1.56) | 1.15 (0.84-1.58) | 0.38 | >0.99 | 1.10 (0.74-1.64) | 1.10 (0.74-1.64) | 0.63 | >0.99 |
| C44:2 TAG | HMDB0042279* | Triglycerides | 1.10 (0.89-1.37) | 0.98 (0.76-1.25) | 0.84 (0.62-1.15) | 0.83 (0.61-1.14) | 0.26 | >0.99 | 0.80 (0.53-1.21) | 0.80 (0.53-1.21) | 0.29 | >0.99 |
| C45:0 TAG | HMDB0042093* | Triglycerides | 1.05 (0.84-1.30) | 0.93 (0.72-1.20) | 0.78 (0.56-1.08) | 0.77 (0.55-1.07) | 0.12 | >0.99 | 0.76 (0.50-1.15) | 0.76 (0.50-1.15) | 0.20 | >0.99 |
| C45:1 TAG | HMDB0042099* | Triglycerides | 1.08 (0.87-1.34) | 0.96 (0.74-1.24) | 0.84 (0.61-1.16) | 0.83 (0.60-1.15) | 0.25 | >0.99 | 0.78 (0.52-1.17) | 0.77 (0.51-1.17) | 0.22 | >0.99 |
| C45:2 TAG | HMDB0043170* | Triglycerides | 1.07 (0.86-1.33) | 0.96 (0.74-1.24) | 0.85 (0.61-1.18) | 0.84 (0.60-1.17) | 0.30 | >0.99 | 0.83 (0.54-1.27) | 0.83 (0.54-1.27) | 0.39 | >0.99 |
| C45:3 TAG | NA | Triglycerides | 1.05 (0.85-1.30) | 1.01 (0.79-1.29) | 0.95 (0.70-1.28) | 0.92 (0.67-1.26) | 0.60 | >0.99 | 0.94 (0.64-1.39) | 0.94 (0.64-1.39) | 0.76 | >0.99 |
| C46:0 TAG | HMDB0010411* | Triglycerides | 1.10 (0.89-1.37) | 1.01 (0.78-1.30) | 0.91 (0.67-1.23) | 0.86 (0.62-1.18) | 0.34 | >0.99 | 0.76 (0.51-1.13) | 0.76 (0.51-1.13) | 0.17 | >0.99 |
| C46:1 TAG | HMDB0010412* | Triglycerides | 1.11 (0.90-1.38) | 1.02 (0.80-1.30) | 0.92 (0.68-1.25) | 0.90 (0.66-1.22) | 0.50 | >0.99 | 0.86 (0.59-1.26) | 0.86 (0.58-1.26) | 0.43 | >0.99 |
| C46:2 TAG | HMDB0010419* | Triglycerides | 1.10 (0.89-1.37) | 0.99 (0.78-1.27) | 0.89 (0.65-1.20) | 0.88 (0.65-1.20) | 0.43 | >0.99 | 0.87 (0.58-1.28) | 0.86 (0.58-1.28) | 0.47 | >0.99 |
| C46:3 TAG | HMDB0042751* | Triglycerides | 1.06 (0.86-1.31) | 0.95 (0.74-1.21) | 0.84 (0.62-1.14) | 0.84 (0.62-1.15) | 0.28 | >0.99 | 0.82 (0.55-1.21) | 0.82 (0.55-1.21) | 0.31 | >0.99 |
| C46:4 TAG | HMDB0042548* | Triglycerides | 1.04 (0.84-1.29) | 0.93 (0.72-1.19) | 0.83 (0.60-1.13) | 0.82 (0.60-1.13) | 0.23 | >0.99 | 0.80 (0.53-1.20) | 0.80 (0.53-1.20) | 0.28 | >0.99 |

| METABOLITE | HMDB_ID | Metabolite class | Model 1 | Model 2 | Model 3 | Model 4 | | | Model 5 | Model 6 | | |
|---|---|---|---|---|---|---|---|---|---|---|---|---|
| | | | OR (95%CI) | OR (95%CI) | OR (95%CI) | OR (95%CI) | p-value | NEF | OR (95%CI) | OR (95%CI) | p-value | NEF |
| C47:0 TAG | HMDB0042094* | Triglycerides | 1.07 (0.86-1.33) | 0.93 (0.72-1.21) | 0.81 (0.59-1.13) | 0.81 (0.58-1.13) | 0.22 | >0.99 | 0.84 (0.57-1.25) | 0.84 (0.57-1.24) | 0.39 | >0.99 |
| C47:1 TAG | HMDB0042100* | Triglycerides | 1.12 (0.91-1.39) | 1.04 (0.81-1.33) | 0.93 (0.68-1.26) | 0.92 (0.67-1.25) | 0.58 | >0.99 | 0.87 (0.59-1.27) | 0.87 (0.59-1.27) | 0.46 | >0.99 |
| C47:2 TAG | HMDB0042076* | Triglycerides | 1.12 (0.91-1.39) | 1.03 (0.81-1.32) | 0.93 (0.69-1.26) | 0.93 (0.68-1.26) | 0.64 | >0.99 | 0.90 (0.62-1.32) | 0.90 (0.61-1.32) | 0.58 | >0.99 |
| C48:0 TAG | HMDB0005356* | Triglycerides | 1.12 (0.90-1.39) | 1.03 (0.80-1.32) | 0.94 (0.70-1.26) | 0.88 (0.64-1.20) | 0.42 | >0.99 | 0.78 (0.53-1.14) | 0.78 (0.53-1.14) | 0.20 | >0.99 |
| C48:1 TAG | HMDB0005359* | Triglycerides | 1.13 (0.92-1.40) | 1.05 (0.82-1.34) | 0.94 (0.70-1.27) | 0.91 (0.67-1.23) | 0.52 | >0.99 | 0.85 (0.59-1.24) | 0.85 (0.58-1.24) | 0.40 | >0.99 |
| C48:2 TAG | HMDB0005376* | Triglycerides | 1.12 (0.91-1.39) | 1.04 (0.82-1.33) | 0.94 (0.70-1.27) | 0.92 (0.68-1.24) | 0.57 | >0.99 | 0.88 (0.61-1.28) | 0.88 (0.60-1.28) | 0.50 | >0.99 |
| C48:3 TAG | HMDB0005432* | Triglycerides | 1.10 (0.89-1.35) | 0.99 (0.78-1.26) | 0.90 (0.67-1.21) | 0.90 (0.67-1.22) | 0.51 | >0.99 | 0.88 (0.61-1.27) | 0.88 (0.60-1.27) | 0.48 | >0.99 |
| C48:4 TAG | HMDB0042811* | Triglycerides | 1.04 (0.84-1.28) | 0.93 (0.74-1.18) | 0.85 (0.63-1.15) | 0.87 (0.64-1.19) | 0.39 | >0.99 | 0.86 (0.59-1.26) | 0.86 (0.59-1.26) | 0.44 | >0.99 |
| C48:5 TAG | HMDB0042789* | Triglycerides | 1.03 (0.84-1.28) | 0.94 (0.74-1.19) | 0.86 (0.64-1.16) | 0.87 (0.64-1.19) | 0.39 | >0.99 | 0.85 (0.58-1.25) | 0.85 (0.58-1.25) | 0.40 | >0.99 |
| C49:0 TAG | HMDB0042095* | Triglycerides | 1.09 (0.88-1.36) | 0.99 (0.77-1.27) | 0.86 (0.64-1.18) | 0.86 (0.63-1.17) | 0.33 | >0.99 | 0.83 (0.57-1.21) | 0.83 (0.57-1.21) | 0.32 | >0.99 |
| C49:1 TAG | HMDB0011705* | Triglycerides | 1.15 (0.93-1.43) | 1.09 (0.85-1.39) | 0.95 (0.71-1.29) | 0.95 (0.70-1.28) | 0.71 | >0.99 | 0.86 (0.59-1.26) | 0.86 (0.59-1.26) | 0.44 | >0.99 |
| C49:2 TAG | HMDB0011706* | Triglycerides | 1.17 (0.95-1.44) | 1.11 (0.87-1.41) | 1.00 (0.75-1.34) | 1.00 (0.75-1.34) | >0.99 | >0.99 | 0.96 (0.68-1.37) | 0.96 (0.68-1.37) | 0.83 | >0.99 |
| C49:3 TAG | HMDB0042103* | Triglycerides | 1.12 (0.91-1.38) | 1.05 (0.83-1.33) | 0.96 (0.72-1.29) | 0.98 (0.73-1.32) | 0.89 | >0.99 | 0.97 (0.68-1.38) | 0.97 (0.68-1.38) | 0.86 | >0.99 |
| C4-OH carnitine | HMDB0013127 | Carnitines | 0.94 (0.77-1.15) | 0.93 (0.74-1.17) | 1.02 (0.77-1.37) | 0.99 (0.73-1.33) | 0.94 | >0.99 | 1.09 (0.78-1.53) | 1.09 (0.77-1.53) | 0.63 | >0.99 |
| C5 carnitine | HMDB0000688 | Carnitines | 1.01 (0.81-1.26) | 1.02 (0.80-1.32) | 0.98 (0.72-1.33) | 0.99 (0.72-1.35) | 0.95 | >0.99 | 1.10 (0.77-1.58) | 1.11 (0.77-1.59) | 0.58 | >0.99 |
| C5:1 carnitine | HMDB0002366 | Carnitines | 1.02 (0.83-1.27) | 1.02 (0.81-1.30) | 1.12 (0.84-1.49) | 1.11 (0.84-1.49) | 0.46 | >0.99 | 1.21 (0.86-1.71) | 1.24 (0.86-1.78) | 0.25 | >0.99 |
| C50:0 TAG | HMDB0005357* | Triglycerides | 1.12 (0.90-1.39) | 1.01 (0.79-1.30) | 0.94 (0.71-1.26) | 0.88 (0.65-1.20) | 0.43 | >0.99 | 0.80 (0.55-1.16) | 0.80 (0.55-1.16) | 0.24 | >0.99 |
| C50:1 TAG | HMDB0005360* | Triglycerides | 1.10 (0.89-1.35) | 1.02 (0.80-1.30) | 0.92 (0.69-1.24) | 0.88 (0.65-1.19) | 0.41 | >0.99 | 0.83 (0.57-1.21) | 0.83 (0.56-1.21) | 0.33 | >0.99 |
| C50:2 TAG | HMDB0005377* | Triglycerides | 1.13 (0.92-1.40) | 1.04 (0.81-1.32) | 0.93 (0.69-1.26) | 0.91 (0.67-1.23) | 0.53 | >0.99 | 0.88 (0.61-1.29) | 0.88 (0.60-1.29) | 0.51 | >0.99 |
| C50:3 TAG | HMDB0005433* | Triglycerides | 1.07 (0.87-1.33) | 0.98 (0.77-1.26) | 0.92 (0.67-1.24) | 0.91 (0.67-1.24) | 0.56 | >0.99 | 0.90 (0.62-1.30) | 0.90 (0.62-1.30) | 0.57 | >0.99 |
| C50:4 TAG | HMDB0005435* | Triglycerides | 1.00 (0.81-1.24) | 0.91 (0.72-1.17) | 0.86 (0.63-1.18) | 0.89 (0.65-1.22) | 0.45 | >0.99 | 0.88 (0.61-1.29) | 0.88 (0.61-1.29) | 0.52 | >0.99 |
| C50:5 TAG | HMDB0010471* | Triglycerides | 1.00 (0.81-1.24) | 0.92 (0.72-1.17) | 0.85 (0.63-1.16) | 0.88 (0.64-1.20) | 0.41 | >0.99 | 0.84 (0.57-1.22) | 0.84 (0.57-1.22) | 0.35 | >0.99 |
| C50:6 TAG | HMDB0010497* | Triglycerides | 1.01 (0.82-1.25) | 0.94 (0.74-1.19) | 0.89 (0.66-1.19) | 0.90 (0.67-1.22) | 0.51 | >0.99 | 0.85 (0.58-1.23) | 0.85 (0.58-1.23) | 0.39 | >0.99 |
| C51:0 TAG | HMDB0031106* | Triglycerides | 1.16 (0.93-1.44) | 1.03 (0.80-1.33) | 0.94 (0.69-1.27) | 0.90 (0.66-1.23) | 0.51 | >0.99 | 0.85 (0.59-1.25) | 0.85 (0.58-1.25) | 0.41 | >0.99 |
| C51:1 TAG | HMDB0042104* | Triglycerides | 1.14 (0.92-1.41) | 1.07 (0.84-1.37) | 0.96 (0.71-1.30) | 0.94 (0.69-1.28) | 0.71 | >0.99 | 0.92 (0.63-1.35) | 0.92 (0.63-1.35) | 0.68 | >0.99 |
| C51:3 TAG | HMDB0011701* | Triglycerides | 1.12 (0.92-1.37) | 1.04 (0.83-1.30) | 0.99 (0.74-1.32) | 1.04 (0.77-1.39) | 0.82 | >0.99 | 1.11 (0.78-1.58) | 1.11 (0.78-1.58) | 0.57 | >0.99 |
| C52:0 TAG | HMDB0005365* | Triglycerides | 1.11 (0.90-1.38) | 0.96 (0.75-1.24) | 0.90 (0.67-1.21) | 0.85 (0.63-1.15) | 0.30 | >0.99 | 0.79 (0.54-1.16) | 0.79 (0.54-1.16) | 0.23 | >0.99 |
| C52:1 TAG | HMDB0005367* | Triglycerides | 1.09 (0.88-1.34) | 0.98 (0.77-1.25) | 0.90 (0.67-1.20) | 0.84 (0.61-1.14) | 0.26 | >0.99 | 0.80 (0.55-1.17) | 0.79 (0.54-1.16) | 0.24 | >0.99 |
| C52:2 TAG | HMDB0005369* | Triglycerides | 1.08 (0.88-1.33) | 0.99 (0.78-1.26) | 0.89 (0.66-1.19) | 0.87 (0.64-1.17) | 0.35 | >0.99 | 0.88 (0.62-1.27) | 0.88 (0.61-1.27) | 0.49 | >0.99 |
| C52:3 TAG | HMDB0005384* | Triglycerides | 1.00 (0.81-1.23) | 0.89 (0.70-1.14) | 0.82 (0.60-1.11) | 0.82 (0.60-1.12) | 0.22 | >0.99 | 0.83 (0.57-1.22) | 0.83 (0.56-1.22) | 0.34 | >0.99 |
| C52:4 TAG | HMDB0005363* | Triglycerides | 0.93 (0.75-1.15) | 0.83 (0.65-1.07) | 0.82 (0.60-1.11) | 0.84 (0.61-1.16) | 0.30 | >0.99 | 0.92 (0.62-1.36) | 0.92 (0.62-1.36) | 0.67 | >0.99 |

| METABOLITE | HMDB_ID | Metabolite class | Model 1 | Model 2 | Model 3 | Model 4 | | | Model 5 | Model 6 | | |
|---|---|---|---|---|---|---|---|---|---|---|---|---|
| | | | OR (95%CI) | OR (95%CI) | OR (95%CI) | OR (95%CI) | p-value | NEF | OR (95%CI) | OR (95%CI) | p-value | NEF |
| C52:5 TAG | HMDB0005380* | Triglycerides | 0.92 (0.74-1.15) | 0.86 (0.67-1.11) | 0.84 (0.61-1.16) | 0.88 (0.63-1.23) | 0.45 | >0.99 | 0.90 (0.59-1.37) | 0.90 (0.59-1.37) | 0.63 | >0.99 |
| C52:6 TAG | HMDB0005436* | Triglycerides | 0.95 (0.76-1.17) | 0.89 (0.70-1.13) | 0.86 (0.63-1.17) | 0.88 (0.64-1.22) | 0.45 | >0.99 | 0.84 (0.57-1.25) | 0.84 (0.57-1.25) | 0.39 | >0.99 |
| C52:7 TAG | HMDB0010517* | Triglycerides | 0.97 (0.79-1.19) | 0.92 (0.73-1.16) | 0.90 (0.67-1.21) | 0.92 (0.68-1.24) | 0.58 | >0.99 | 0.85 (0.59-1.23) | 0.85 (0.58-1.24) | 0.40 | >0.99 |
| C53:2 TAG | HMDB0042196* | Triglycerides | 1.18 (0.96-1.46) | 1.11 (0.88-1.41) | 1.04 (0.77-1.41) | 1.05 (0.77-1.42) | 0.76 | >0.99 | 1.11 (0.78-1.60) | 1.11 (0.78-1.60) | 0.56 | >0.99 |
| C53:3 TAG | HMDB0043058* | Triglycerides | 1.07 (0.87-1.30) | 1.02 (0.81-1.29) | 1.01 (0.76-1.36) | 1.07 (0.79-1.44) | 0.68 | >0.99 | 1.19 (0.83-1.71) | 1.19 (0.83-1.71) | 0.34 | >0.99 |
| C54:1 TAG | HMDB0005395* | Triglycerides | 1.08 (0.87-1.33) | 0.95 (0.74-1.21) | 0.86 (0.64-1.15) | 0.80 (0.58-1.09) | 0.16 | >0.99 | 0.77 (0.52-1.14) | 0.76 (0.51-1.13) | 0.18 | >0.99 |
| C54:10 TAG | NA | Triglycerides | 0.94 (0.75-1.17) | 1.02 (0.79-1.31) | 1.00 (0.73-1.35) | 0.98 (0.72-1.35) | 0.92 | >0.99 | 1.05 (0.73-1.50) | 1.05 (0.73-1.50) | 0.80 | >0.99 |
| C54:2 TAG | HMDB0005403* | Triglycerides | 1.09 (0.89-1.34) | 0.98 (0.77-1.25) | 0.89 (0.66-1.20) | 0.85 (0.63-1.15) | 0.30 | >0.99 | 0.89 (0.62-1.28) | 0.88 (0.61-1.28) | 0.51 | >0.99 |
| C54:3 TAG | HMDB0005405* | Triglycerides | 1.04 (0.85-1.27) | 0.91 (0.71-1.16) | 0.86 (0.64-1.17) | 0.86 (0.64-1.17) | 0.34 | >0.99 | 0.95 (0.66-1.35) | 0.94 (0.66-1.35) | 0.76 | >0.99 |
| C54:4 TAG | HMDB0005370* | Triglycerides | 0.97 (0.79-1.20) | 0.86 (0.66-1.10) | 0.85 (0.62-1.16) | 0.88 (0.64-1.21) | 0.43 | >0.99 | 0.98 (0.67-1.44) | 0.98 (0.67-1.44) | 0.91 | >0.99 |
| C54:5 TAG | HMDB0005385* | Triglycerides | 0.94 (0.77-1.16) | 0.83 (0.65-1.06) | 0.83 (0.62-1.12) | 0.87 (0.64-1.19) | 0.38 | >0.99 | 0.99 (0.66-1.48) | 0.99 (0.66-1.48) | 0.96 | >0.99 |
| C54:6 TAG | HMDB0005391* | Triglycerides | 0.92 (0.75-1.14) | 0.83 (0.65-1.06) | 0.83 (0.61-1.12) | 0.86 (0.63-1.18) | 0.36 | >0.99 | 0.95 (0.62-1.45) | 0.95 (0.62-1.46) | 0.81 | >0.99 |
| C54:7 TAG | HMDB0005447* | Triglycerides | 0.88 (0.72-1.09) | 0.83 (0.65-1.06) | 0.89 (0.66-1.20) | 0.95 (0.69-1.31) | 0.76 | >0.99 | 1.06 (0.70-1.61) | 1.07 (0.70-1.63) | 0.77 | >0.99 |
| C54:8 TAG | HMDB0010518* | Triglycerides | 0.94 (0.76-1.15) | 0.90 (0.72-1.14) | 0.93 (0.70-1.26) | 0.96 (0.71-1.30) | 0.78 | >0.99 | 0.93 (0.63-1.36) | 0.92 (0.62-1.37) | 0.70 | >0.99 |
| C54:9 TAG | HMDB0010498* | Triglycerides | 0.93 (0.75-1.14) | 0.94 (0.74-1.19) | 0.99 (0.74-1.31) | 1.00 (0.75-1.34) | >0.99 | >0.99 | 0.92 (0.65-1.31) | 0.92 (0.64-1.32) | 0.65 | >0.99 |
| C55:2 TAG | HMDB0042226* | Triglycerides | 1.20 (0.97-1.50) | 1.09 (0.85-1.40) | 1.04 (0.76-1.42) | 1.01 (0.74-1.39) | 0.95 | >0.99 | 1.07 (0.73-1.56) | 1.07 (0.73-1.56) | 0.73 | >0.99 |
| C55:3 TAG | HMDB0042466* | Triglycerides | 1.17 (0.96-1.43) | 1.10 (0.87-1.38) | 1.17 (0.87-1.57) | 1.21 (0.89-1.63) | 0.22 | >0.99 | 1.36 (0.96-1.94) | 1.36 (0.96-1.94) | 0.09 | >0.99 |
| C56:1 TAG | HMDB0005396* | Triglycerides | 1.03 (0.83-1.27) | 0.89 (0.70-1.15) | 0.80 (0.59-1.09) | 0.74 (0.54-1.02) | 0.07 | >0.99 | 0.66 (0.44-0.99) | 0.66 (0.44-0.99) | 0.04 | >0.99 |
| C56:10 TAG | HMDB0010513* | Triglycerides | 0.94 (0.74-1.18) | 0.94 (0.72-1.22) | 1.03 (0.75-1.41) | 1.02 (0.74-1.42) | 0.89 | >0.99 | 0.88 (0.57-1.34) | 0.85 (0.55-1.32) | 0.47 | >0.99 |
| C56:2 TAG | HMDB0005404* | Triglycerides | 1.09 (0.88-1.36) | 0.98 (0.76-1.27) | 0.96 (0.71-1.30) | 0.90 (0.66-1.24) | 0.54 | >0.99 | 0.90 (0.62-1.31) | 0.90 (0.62-1.31) | 0.59 | >0.99 |
| C56:3 TAG | HMDB0005410* | Triglycerides | 1.13 (0.92-1.40) | 1.02 (0.80-1.31) | 1.04 (0.78-1.39) | 1.01 (0.75-1.36) | 0.96 | >0.99 | 1.06 (0.75-1.49) | 1.06 (0.75-1.49) | 0.74 | >0.99 |
| C56:4 TAG | HMDB0005398* | Triglycerides | 1.11 (0.89-1.38) | 1.01 (0.78-1.30) | 1.05 (0.77-1.43) | 1.05 (0.76-1.44) | 0.77 | >0.99 | 1.14 (0.78-1.65) | 1.14 (0.78-1.65) | 0.51 | >0.99 |
| C56:5 TAG | HMDB0005406* | Triglycerides | 1.10 (0.89-1.35) | 1.08 (0.85-1.38) | 1.07 (0.80-1.44) | 1.05 (0.78-1.41) | 0.75 | >0.99 | 1.12 (0.78-1.62) | 1.12 (0.78-1.62) | 0.53 | >0.99 |
| C56:6 TAG | HMDB0005456* | Triglycerides | 1.01 (0.82-1.25) | 0.99 (0.77-1.26) | 1.00 (0.75-1.34) | 1.00 (0.74-1.34) | 0.98 | >0.99 | 0.98 (0.68-1.40) | 0.98 (0.68-1.41) | 0.91 | >0.99 |
| C56:7 TAG | HMDB0005462* | Triglycerides | 0.96 (0.78-1.19) | 0.91 (0.72-1.16) | 0.95 (0.71-1.28) | 0.95 (0.71-1.29) | 0.76 | >0.99 | 0.97 (0.66-1.41) | 0.97 (0.66-1.42) | 0.87 | >0.99 |
| C56:8 TAG | HMDB0005392* | Triglycerides | 0.95 (0.77-1.17) | 0.92 (0.72-1.17) | 0.96 (0.72-1.29) | 0.96 (0.71-1.29) | 0.78 | >0.99 | 1.00 (0.69-1.46) | 1.01 (0.68-1.48) | 0.98 | >0.99 |
| C56:9 TAG | HMDB0005448* | Triglycerides | 0.94 (0.76-1.16) | 0.91 (0.72-1.16) | 0.97 (0.72-1.30) | 0.97 (0.72-1.31) | 0.85 | >0.99 | 0.95 (0.65-1.38) | 0.95 (0.64-1.40) | 0.79 | >0.99 |
| C58:10 TAG | HMDB0005476* | Triglycerides | 0.93 (0.76-1.16) | 0.93 (0.73-1.19) | 0.99 (0.74-1.32) | 0.98 (0.73-1.31) | 0.88 | >0.99 | 0.99 (0.69-1.40) | 0.99 (0.68-1.42) | 0.94 | >0.99 |
| C58:11 TAG | HMDB0010531* | Triglycerides | 0.97 (0.79-1.20) | 0.97 (0.76-1.24) | 1.05 (0.79-1.40) | 1.04 (0.78-1.39) | 0.79 | >0.99 | 1.03 (0.73-1.48) | 1.04 (0.72-1.50) | 0.83 | >0.99 |
| C58:6 TAG | HMDB0005458* | Triglycerides | 1.03 (0.83-1.29) | 1.02 (0.80-1.31) | 1.03 (0.77-1.37) | 1.01 (0.75-1.35) | 0.96 | >0.99 | 1.02 (0.71-1.47) | 1.02 (0.71-1.48) | 0.90 | >0.99 |
| C58:7 TAG | HMDB0005471* | Triglycerides | 1.03 (0.83-1.28) | 1.04 (0.81-1.33) | 1.09 (0.81-1.47) | 1.09 (0.81-1.47) | 0.57 | >0.99 | 1.13 (0.79-1.62) | 1.14 (0.79-1.64) | 0.50 | >0.99 |

| METABOLITE | HMDB_ID | Metabolite class | Model 1 | Model 2 | Model 3 | Model 4 | | | Model 5 | Model 6 | | |
|---|---|---|---|---|---|---|---|---|---|---|---|---|
| | | | OR (95%CI) | OR (95%CI) | OR (95%CI) | OR (95%CI) | p-value | NEF | OR (95%CI) | OR (95%CI) | p-value | NEF |
| C58:8 TAG | HMDB0005413* | Triglycerides | 0.99 (0.80-1.23) | 0.98 (0.77-1.25) | 1.08 (0.81-1.45) | 1.09 (0.81-1.47) | 0.58 | >0.99 | 1.15 (0.80-1.65) | 1.16 (0.79-1.68) | 0.45 | >0.99 |
| C58:9 TAG | HMDB0005463* | Triglycerides | 0.98 (0.79-1.21) | 0.98 (0.77-1.25) | 1.07 (0.80-1.43) | 1.06 (0.79-1.42) | 0.69 | >0.99 | 1.11 (0.78-1.57) | 1.12 (0.78-1.61) | 0.55 | >0.99 |
| C5-DC carnitine | HMDB0013130 | Carnitines | 1.02 (0.84-1.25) | 1.02 (0.82-1.28) | 1.09 (0.83-1.43) | 1.11 (0.83-1.47) | 0.48 | >0.99 | 1.21 (0.86-1.71) | 1.23 (0.87-1.75) | 0.25 | >0.99 |
| C6 carnitine | HMDB0000705 | Carnitines | 0.85 (0.69-1.05) | 0.85 (0.67-1.06) | 0.93 (0.71-1.22) | 0.93 (0.70-1.23) | 0.61 | >0.99 | 1.02 (0.73-1.41) | 1.02 (0.73-1.41) | 0.93 | >0.99 |
| C60:12 TAG | HMDB0005478* | Triglycerides | 0.99 (0.80-1.22) | 1.01 (0.78-1.29) | 1.07 (0.80-1.43) | 1.04 (0.77-1.40) | 0.80 | >0.99 | 1.05 (0.73-1.50) | 1.05 (0.73-1.52) | 0.78 | >0.99 |
| C7 carnitine | HMDB0013238 | Carnitines | 0.84 (0.67-1.04) | 0.81 (0.64-1.04) | 0.87 (0.66-1.16) | 0.88 (0.66-1.17) | 0.38 | >0.99 | 1.03 (0.73-1.45) | 1.03 (0.73-1.45) | 0.86 | >0.99 |
| C8 carnitine | HMDB0000791 | Carnitines | 0.77 (0.63-0.95) | 0.75 (0.59-0.96) | 0.81 (0.61-1.07) | 0.81 (0.61-1.09) | 0.16 | >0.99 | 0.89 (0.63-1.25) | 0.89 (0.63-1.25) | 0.50 | >0.99 |
| caffeine | HMDB0001847 | Imidazopyrimidines | 1.22 (0.98-1.50) | 1.18 (0.93-1.49) | 1.21 (0.91-1.60) | 1.21 (0.89-1.64) | 0.23 | >0.99 | 1.40 (0.97-2.02) | 1.41 (0.97-2.04) | 0.07 | >0.99 |
| campesterol | HMDB0002869 | NA | 1.03 (0.82-1.28) | 1.00 (0.78-1.28) | 1.09 (0.81-1.48) | 1.04 (0.75-1.43) | 0.81 | >0.99 | 0.92 (0.61-1.40) | 0.92 (0.61-1.40) | 0.71 | >0.99 |
| carnitine | HMDB0000062 | Carnitines | 0.89 (0.71-1.13) | 0.86 (0.66-1.12) | 0.92 (0.67-1.25) | 0.90 (0.66-1.24) | 0.52 | >0.99 | 1.01 (0.69-1.46) | 1.01 (0.70-1.46) | 0.96 | >0.99 |
| cholesterol | HMDB0000067 | NA | 1.02 (0.82-1.27) | 1.00 (0.79-1.27) | 1.06 (0.79-1.43) | 1.01 (0.74-1.39) | 0.93 | >0.99 | 0.92 (0.61-1.38) | 0.92 (0.61-1.38) | 0.68 | >0.99 |
| cinnamoylglycine | HMDB0011621 | Organic acids and derivatives | 1.18 (0.96-1.44) | 1.12 (0.89-1.41) | 1.25 (0.95-1.66) | 1.29 (0.96-1.73) | 0.10 | >0.99 | 1.46 (1.02-2.10) | 1.46 (1.02-2.10) | 0.04 | >0.99 |
| citrulline | HMDB0000904 | Organic acids and derivatives | 1.09 (0.89-1.34) | 1.04 (0.81-1.33) | 1.02 (0.74-1.41) | 1.05 (0.76-1.47) | 0.75 | >0.99 | 1.10 (0.72-1.66) | 1.09 (0.72-1.66) | 0.67 | >0.99 |
| cortisol | HMDB0000063 | Steroids and steroid derivatives | 0.88 (0.72-1.09) | 0.94 (0.73-1.22) | 0.79 (0.57-1.09) | 0.76 (0.54-1.07) | 0.11 | >0.99 | 0.75 (0.49-1.15) | 0.75 (0.49-1.15) | 0.19 | >0.99 |
| cortisone | HMDB0002802 | Steroids and steroid derivatives | 0.75 (0.61-0.92) | 0.75 (0.59-0.95) | 0.53 (0.38-0.74) | 0.51 (0.36-0.73) | 0.0002 | 0.00 | 0.49 (0.32-0.74) | 0.49 (0.32-0.74) | 0.001 | 0.05 |
| cotinine | HMDB0001046 | Organoheterocyclic compounds | 1.00 (0.80-1.24) | 0.85 (0.64-1.13) | 0.82 (0.57-1.19) | 0.78 (0.53-1.15) | 0.21 | >0.99 | 0.89 (0.56-1.42) | 0.89 (0.56-1.42) | 0.62 | >0.99 |
| cotinine N-oxide | HMDB0001411 | NA | 1.01 (0.36-2.82) | NA | NA | NA | NA | NA | NA | NA | NA | NA |
| creatine | HMDB0000064 | Organic acids and derivatives | 1.37 (1.10-1.71) | 1.33 (1.04-1.69) | 1.40 (1.04-1.89) | 1.40 (1.04-1.90) | 0.03 | >0.99 | 1.73 (1.20-2.50) | 1.73 (1.20-2.51) | 0.004 | 0.25 |
| creatinine | HMDB0000562 | Carboxylic acids and derivatives | 1.05 (0.86-1.29) | 1.00 (0.79-1.27) | 0.98 (0.74-1.30) | 0.99 (0.74-1.32) | 0.94 | >0.99 | 1.08 (0.76-1.54) | 1.08 (0.76-1.55) | 0.66 | >0.99 |
| cyclohexylamine | HMDB0031404 | NA | 1.06 (0.84-1.33) | 1.06 (0.81-1.38) | 1.18 (0.85-1.65) | 1.19 (0.85-1.67) | 0.31 | >0.99 | 1.20 (0.80-1.78) | 1.20 (0.80-1.78) | 0.37 | >0.99 |
| cystine | HMDB0000192 | NA | 1.04 (0.81-1.34) | 1.07 (0.81-1.42) | 0.91 (0.64-1.29) | 0.91 (0.64-1.31) | 0.62 | >0.99 | 0.86 (0.57-1.30) | 0.86 (0.57-1.30) | 0.47 | >0.99 |
| diacetylspermine | HMDB0002172 | Organic acids and derivatives | 1.09 (0.88-1.34) | 1.03 (0.82-1.29) | 1.07 (0.80-1.41) | 1.05 (0.79-1.40) | 0.73 | >0.99 | 1.12 (0.79-1.60) | 1.12 (0.79-1.60) | 0.53 | >0.99 |
| dimethylglycine | HMDB0000092 | Carboxylic acids and derivatives | 0.94 (0.76-1.16) | 0.92 (0.72-1.17) | 0.84 (0.62-1.14) | 0.86 (0.62-1.19) | 0.36 | >0.99 | 0.75 (0.51-1.11) | 0.75 (0.50-1.11) | 0.15 | >0.99 |
| DMGV | HMDB0240212 | NA | 1.12 (0.90-1.39) | 1.09 (0.84-1.41) | 0.82 (0.60-1.13) | 0.83 (0.60-1.15) | 0.26 | >0.99 | 0.93 (0.64-1.35) | 0.93 (0.64-1.35) | 0.71 | >0.99 |
| ectoine | NA | NA | 1.11 (0.92-1.35) | 1.16 (0.92-1.45) | 1.26 (0.95-1.68) | 1.23 (0.92-1.66) | 0.16 | >0.99 | 1.27 (0.89-1.81) | 1.27 (0.89-1.81) | 0.19 | >0.99 |
| gabapentin | HMDB0005015 | NA | 1.44 (1.11-1.87) | 1.54 (1.15-2.06) | 1.89 (1.27-2.82) | 1.90 (1.26-2.87) | 0.002 | 0.00 | 2.20 (1.31-3.72) | 2.21 (1.31-3.73) | 0.003 | 0.21 |
| glutamine | HMDB0000641 | Organic acids and derivatives | 0.85 (0.68-1.05) | 0.81 (0.63-1.04) | 0.74 (0.53-1.03) | 0.76 (0.54-1.06) | 0.10 | >0.99 | 0.73 (0.48-1.13) | 0.73 (0.48-1.13) | 0.16 | >0.99 |
| glycine | HMDB0000123 | Carboxylic acids and derivatives | 1.33 (1.07-1.66) | 1.36 (1.06-1.76) | 1.36 (0.98-1.88) | 1.36 (0.98-1.90) | 0.07 | >0.99 | 1.55 (1.02-2.33) | 1.55 (1.02-2.34) | 0.04 | >0.99 |
| glycocholate | HMDB0000138 | Steroids and steroid derivatives | 1.14 (0.93-1.39) | 1.05 (0.84-1.32) | 1.00 (0.76-1.32) | 1.03 (0.77-1.38) | 0.86 | >0.99 | 0.95 (0.68-1.33) | 0.95 (0.67-1.34) | 0.76 | >0.99 |
| glycodeoxycholate/glycochenodeoxycholate | HMDB0000631* | Steroids and steroid derivatives | 1.00 (0.81-1.23) | 0.93 (0.74-1.17) | 0.84 (0.62-1.12) | 0.85 (0.63-1.14) | 0.27 | >0.99 | 0.79 (0.55-1.12) | 0.78 (0.54-1.11) | 0.16 | >0.99 |
| guanidinoacetic acid | HMDB0000128 | Carboxylic acids and derivatives | 1.21 (0.97-1.51) | 1.11 (0.86-1.43) | 1.03 (0.76-1.39) | 1.05 (0.77-1.42) | 0.77 | >0.99 | 1.09 (0.77-1.56) | 1.09 (0.76-1.57) | 0.62 | >0.99 |

| METABOLITE | HMDB_ID | Metabolite class | Model 1 | Model 2 | Model 3 | Model 4 | | | Model 5 | Model 6 | | |
|---|---|---|---|---|---|---|---|---|---|---|---|---|
| | | | OR (95%CI) | OR (95%CI) | OR (95%CI) | OR (95%CI) | p-value | NEF | OR (95%CI) | OR (95%CI) | p-value | NEF |
| guanine | HMDB0000132 | NA | 0.84 (0.45-1.57) | NA | NA | NA | NA | NA | NA | NA | NA | NA |
| guanosine | HMDB0000133 | NA | 0.90 (0.73-1.11) | 0.98 (0.77-1.23) | 0.90 (0.68-1.19) | 0.93 (0.70-1.25) | 0.64 | >0.99 | 1.06 (0.76-1.49) | 1.07 (0.76-1.50) | 0.71 | >0.99 |
| hippurate | HMDB0000714 | Benzene and substituted derivatives | 1.09 (0.90-1.33) | 1.01 (0.81-1.25) | 0.95 (0.73-1.25) | 0.96 (0.73-1.26) | 0.76 | >0.99 | 1.10 (0.80-1.52) | 1.10 (0.80-1.52) | 0.56 | >0.99 |
| histidine | HMDB0000177 | Carboxylic acids and derivatives | 0.89 (0.71-1.13) | 0.84 (0.65-1.10) | 0.88 (0.63-1.23) | 0.86 (0.61-1.22) | 0.40 | >0.99 | 0.90 (0.59-1.38) | 0.90 (0.59-1.37) | 0.62 | >0.99 |
| homoarginine | HMDB0000670 | Organic acids and derivatives | 0.81 (0.64-1.02) | 0.79 (0.61-1.02) | 0.80 (0.58-1.11) | 0.80 (0.58-1.13) | 0.20 | >0.99 | 0.90 (0.61-1.32) | 0.90 (0.61-1.32) | 0.59 | >0.99 |
| homocitrulline | HMDB0000679 | Organic acids and derivatives | 1.11 (0.89-1.37) | 1.11 (0.87-1.43) | 1.06 (0.78-1.43) | 1.08 (0.79-1.47) | 0.64 | >0.99 | 1.05 (0.73-1.51) | 1.05 (0.73-1.52) | 0.78 | >0.99 |
| hydroxycotinine | HMDB0001390 | Organoheterocyclic compounds | 1.10 (0.89-1.36) | 0.96 (0.73-1.26) | 1.05 (0.75-1.46) | 1.00 (0.71-1.43) | 0.98 | >0.99 | 1.05 (0.70-1.59) | 1.05 (0.70-1.59) | 0.81 | >0.99 |
| hydroxyectoine | NA | NA | 1.08 (0.88-1.32) | 1.03 (0.81-1.32) | 1.03 (0.75-1.42) | 1.06 (0.77-1.47) | 0.72 | >0.99 | 1.10 (0.73-1.65) | 1.10 (0.73-1.65) | 0.65 | >0.99 |
| hydroxyproline | HMDB0000725 | Carboxylic acids and derivatives | 1.07 (0.87-1.31) | 1.07 (0.84-1.35) | 1.14 (0.86-1.52) | 1.13 (0.84-1.51) | 0.42 | >0.99 | 1.05 (0.74-1.48) | 1.07 (0.73-1.55) | 0.74 | >0.99 |
| hypoxanthine | HMDB0000157 | NA | 1.05 (0.62-1.79) | NA | NA | NA | NA | NA | NA | NA | NA | NA |
| imidazole propionate | HMDB0002271 | Organoheterocyclic compounds | 1.02 (0.83-1.27) | 0.98 (0.77-1.25) | 0.90 (0.67-1.20) | 0.91 (0.67-1.23) | 0.53 | >0.99 | 0.73 (0.51-1.04) | 0.73 (0.51-1.04) | 0.08 | >0.99 |
| inosine | HMDB0000195 | NA | 1.02 (0.62-1.70) | NA | NA | NA | NA | NA | NA | NA | NA | NA |
| isoleucine | HMDB0000172 | Organic acids and derivatives | 0.94 (0.77-1.16) | 0.86 (0.67-1.11) | 0.74 (0.55-1.00) | 0.71 (0.51-0.99) | 0.04 | >0.99 | 0.67 (0.44-1.02) | 0.67 (0.44-1.02) | 0.06 | >0.99 |
| kynurenic acid | HMDB0000715 | NA | 1.01 (0.80-1.27) | 0.99 (0.75-1.30) | 0.80 (0.57-1.12) | 0.77 (0.54-1.10) | 0.15 | >0.99 | 0.87 (0.58-1.31) | 0.87 (0.58-1.31) | 0.51 | >0.99 |
| L-alpha-glutamyl-L-Lysine | HMDB0004207 | NA | 1.00 (0.80-1.25) | 0.88 (0.68-1.13) | 0.84 (0.62-1.15) | 0.87 (0.63-1.19) | 0.38 | >0.99 | 0.96 (0.66-1.40) | 0.96 (0.66-1.39) | 0.82 | >0.99 |
| leucine | HMDB0000687 | Carboxylic acids and derivatives | 0.92 (0.75-1.13) | 0.85 (0.66-1.09) | 0.80 (0.59-1.08) | 0.78 (0.56-1.07) | 0.13 | >0.99 | 0.76 (0.50-1.14) | 0.75 (0.50-1.14) | 0.18 | >0.99 |
| lysine | HMDB0000182 | Carboxylic acids and derivatives | 0.97 (0.78-1.20) | 0.89 (0.70-1.13) | 0.90 (0.67-1.21) | 0.92 (0.68-1.25) | 0.59 | >0.99 | 1.01 (0.70-1.45) | 1.01 (0.70-1.45) | 0.96 | >0.99 |
| methionine | HMDB0000696 | Organic acids and derivatives | 0.89 (0.73-1.09) | 0.85 (0.67-1.08) | 0.82 (0.61-1.10) | 0.82 (0.61-1.11) | 0.20 | >0.99 | 0.87 (0.61-1.23) | 0.87 (0.61-1.23) | 0.43 | >0.99 |
| methionine sulfoxide | HMDB0002005 | NA | 1.45 (1.14-1.85) | 1.23 (0.94-1.62) | 1.15 (0.84-1.59) | 1.15 (0.83-1.58) | 0.40 | >0.99 | 1.13 (0.76-1.66) | 1.13 (0.76-1.67) | 0.55 | >0.99 |
| methylimidazoleacetic acid | HMDB0002820 | Organoheterocyclic compounds | 1.15 (0.94-1.40) | 1.15 (0.92-1.44) | 1.12 (0.86-1.47) | 1.11 (0.84-1.46) | 0.48 | >0.99 | 1.05 (0.75-1.48) | 1.06 (0.75-1.49) | 0.75 | >0.99 |
| metronidazole | HMDB0015052 | NA | 0.97 (0.74-1.26) | 0.99 (0.72-1.36) | 1.00 (0.68-1.47) | 0.96 (0.64-1.44) | 0.84 | >0.99 | 0.83 (0.50-1.40) | 0.83 (0.49-1.41) | 0.49 | >0.99 |
| myristoleate | HMDB0002000 | NA | 0.92 (0.74-1.15) | 0.88 (0.68-1.13) | 0.92 (0.68-1.25) | 0.92 (0.68-1.26) | 0.61 | >0.99 | 1.00 (0.69-1.45) | 1.00 (0.69-1.45) | 0.99 | >0.99 |
| N1-acetylspermidine | HMDB0001276 | Organic acids and derivatives | 1.02 (0.82-1.28) | 1.06 (0.81-1.37) | 1.08 (0.78-1.49) | 1.11 (0.80-1.55) | 0.52 | >0.99 | 1.39 (0.92-2.09) | 1.39 (0.92-2.10) | 0.12 | >0.99 |
| N1-methyl-2-pyridone-5-carboxamide | HMDB0004193 | Organoheterocyclic compounds | 1.09 (0.87-1.35) | 1.06 (0.84-1.35) | 1.20 (0.89-1.61) | 1.23 (0.90-1.70) | 0.20 | >0.99 | 1.34 (0.92-1.96) | 1.34 (0.92-1.97) | 0.13 | >0.99 |
| N2,N2-dimethylguanosine | HMDB0004824 | Nucleosides, nucleotides, and analogues | 1.18 (0.96-1.45) | 1.17 (0.92-1.48) | 1.15 (0.86-1.53) | 1.14 (0.85-1.53) | 0.37 | >0.99 | 1.29 (0.93-1.80) | 1.29 (0.92-1.80) | 0.13 | >0.99 |
| N4-acetylcytidine | HMDB0005923 | Nucleosides, nucleotides, and analogues | 0.97 (0.79-1.20) | 0.99 (0.78-1.26) | 0.90 (0.68-1.21) | 0.87 (0.64-1.19) | 0.38 | >0.99 | 0.96 (0.65-1.41) | 0.96 (0.65-1.41) | 0.82 | >0.99 |
| N6,N6,N6-trimethyllysine | HMDB0001325 | Organic acids and derivatives | 1.06 (0.86-1.31) | 1.05 (0.83-1.34) | 1.03 (0.76-1.39) | 1.02 (0.75-1.39) | 0.88 | >0.99 | 1.09 (0.75-1.59) | 1.10 (0.75-1.61) | 0.62 | >0.99 |
| N6,N6-dimethyllysine | HMDB0013287 | Organic acids and derivatives | 1.12 (0.92-1.38) | 1.06 (0.85-1.34) | 0.92 (0.69-1.23) | 0.90 (0.67-1.21) | 0.49 | >0.99 | 0.79 (0.54-1.15) | 0.79 (0.54-1.15) | 0.22 | >0.99 |
| N6-acetyllysine | HMDB0000206 | Organic acids and derivatives | 1.12 (0.92-1.37) | 1.08 (0.86-1.36) | 1.00 (0.75-1.31) | 0.98 (0.74-1.30) | 0.88 | >0.99 | 0.94 (0.67-1.32) | 0.94 (0.66-1.32) | 0.71 | >0.99 |
| N6-methyllysine | HMDB0002038 | NA | 0.97 (0.79-1.19) | 0.93 (0.73-1.17) | 0.85 (0.64-1.13) | 0.83 (0.62-1.11) | 0.21 | >0.99 | 0.76 (0.53-1.09) | 0.76 (0.53-1.09) | 0.14 | >0.99 |
| N-acetylaspartic acid | HMDB0000812 | Organic acids and derivatives | 1.32 (1.05-1.65) | 1.22 (0.95-1.58) | 1.16 (0.86-1.58) | 1.15 (0.84-1.58) | 0.37 | >0.99 | 1.10 (0.75-1.62) | 1.10 (0.74-1.62) | 0.64 | >0.99 |

| METABOLITE | HMDB_ID | Metabolite class | Model 1 | Model 2 | Model 3 | Model 4 | | | Model 5 | Model 6 | | |
|---|---|---|---|---|---|---|---|---|---|---|---|---|
| | | | OR (95%CI) | OR (95%CI) | OR (95%CI) | OR (95%CI) | p-value | NEF | OR (95%CI) | OR (95%CI) | p-value | NEF |
| N-acetylornithine | HMDB0003357 | Organic acids and derivatives | 0.93 (0.77-1.13) | 0.88 (0.70-1.09) | 0.91 (0.70-1.17) | 0.88 (0.67-1.15) | 0.34 | >0.99 | 0.89 (0.64-1.24) | 0.89 (0.64-1.24) | 0.50 | >0.99 |
| N-acetylputrescine | HMDB0002064 | Organic acids and derivatives | 0.96 (0.77-1.20) | 0.92 (0.72-1.18) | 0.94 (0.70-1.27) | 0.93 (0.69-1.27) | 0.66 | >0.99 | 0.85 (0.58-1.24) | 0.85 (0.58-1.25) | 0.41 | >0.99 |
| N-acetyltryptophan | HMDB0013713 | Organic acids and derivatives | 1.17 (0.95-1.44) | 1.04 (0.81-1.33) | 0.92 (0.68-1.26) | 0.91 (0.66-1.25) | 0.56 | >0.99 | 0.83 (0.57-1.21) | 0.83 (0.57-1.21) | 0.34 | >0.99 |
| N-alpha-acetylarginine | HMDB0004620 | Organic acids and derivatives | 1.07 (0.86-1.33) | 1.07 (0.83-1.37) | 1.16 (0.86-1.56) | 1.19 (0.88-1.63) | 0.26 | >0.99 | 1.11 (0.77-1.58) | 1.11 (0.77-1.58) | 0.58 | >0.99 |
| N-carbamoyl-beta-alanine | HMDB0000026 | Organic acids and derivatives | 0.84 (0.67-1.04) | 0.80 (0.62-1.03) | 0.86 (0.63-1.18) | 0.84 (0.61-1.16) | 0.30 | >0.99 | 0.99 (0.66-1.49) | 0.99 (0.66-1.49) | 0.96 | >0.99 |
| N-methylproline | NA | NA | 1.02 (0.84-1.24) | 1.00 (0.79-1.25) | 0.96 (0.73-1.25) | 0.95 (0.72-1.26) | 0.72 | >0.99 | 0.92 (0.66-1.28) | 0.92 (0.66-1.28) | 0.61 | >0.99 |
| NMMA | HMDB0029416 | Organic acids and derivatives | 1.00 (0.79-1.26) | 0.97 (0.74-1.27) | 0.94 (0.68-1.30) | 0.92 (0.65-1.31) | 0.65 | >0.99 | 0.83 (0.54-1.29) | 0.84 (0.54-1.30) | 0.43 | >0.99 |
| N-oleoylethanolamine | HMDB0002088 | NA | 0.88 (0.72-1.09) | 0.92 (0.73-1.16) | 0.91 (0.69-1.21) | 0.91 (0.68-1.22) | 0.54 | >0.99 | 1.07 (0.75-1.52) | 1.07 (0.75-1.53) | 0.71 | >0.99 |
| palmitoylethanolamide | HMDB0002100 | NA | 0.87 (0.68-1.12) | 0.87 (0.66-1.14) | 0.87 (0.62-1.22) | 0.88 (0.62-1.26) | 0.48 | >0.99 | 0.85 (0.55-1.31) | 0.85 (0.55-1.31) | 0.46 | >0.99 |
| pantothenate | HMDB0000210 | Carboxylic acids and derivatives | 1.00 (0.81-1.22) | 0.98 (0.78-1.22) | 1.01 (0.76-1.34) | 0.99 (0.70-1.39) | 0.96 | >0.99 | 1.07 (0.71-1.60) | 1.07 (0.71-1.60) | 0.75 | >0.99 |
| phenylacetylglutamine | HMDB0006344 | Organic acids and derivatives | 1.22 (1.00-1.48) | 1.17 (0.94-1.45) | 1.10 (0.85-1.44) | 1.10 (0.84-1.44) | 0.49 | >0.99 | 1.15 (0.84-1.58) | 1.16 (0.85-1.58) | 0.36 | >0.99 |
| phenylalanine | HMDB0000159 | Organic acids and derivatives | 0.99 (0.82-1.21) | 0.95 (0.76-1.19) | 0.98 (0.75-1.28) | 1.00 (0.76-1.32) | 0.97 | >0.99 | 1.11 (0.78-1.58) | 1.11 (0.78-1.59) | 0.56 | >0.99 |
| phenylalanine-d8 | NA | NA | 1.06 (0.84-1.32) | 1.13 (0.87-1.46) | 1.12 (0.82-1.53) | 1.17 (0.85-1.60) | 0.35 | >0.99 | 1.36 (0.88-2.12) | 1.37 (0.88-2.12) | 0.17 | >0.99 |
| pipecolic acid | HMDB0000716 | Organic acids and derivatives | 0.95 (0.78-1.16) | 0.99 (0.79-1.23) | 1.06 (0.81-1.39) | 1.06 (0.80-1.40) | 0.68 | >0.99 | 1.16 (0.82-1.64) | 1.16 (0.82-1.65) | 0.40 | >0.99 |
| piperine | HMDB0029377 | Alkaloids and derivatives | 0.92 (0.76-1.12) | 0.96 (0.76-1.20) | 1.05 (0.79-1.39) | 1.01 (0.76-1.36) | 0.93 | >0.99 | 1.16 (0.83-1.64) | 1.17 (0.83-1.65) | 0.37 | >0.99 |
| proline | HMDB0000162 | Carboxylic acids and derivatives | 1.04 (0.84-1.28) | 0.97 (0.76-1.24) | 0.89 (0.66-1.20) | 0.90 (0.66-1.23) | 0.50 | >0.99 | 0.99 (0.68-1.44) | 0.99 (0.68-1.44) | 0.96 | >0.99 |
| proline-betaine | HMDB0004827 | Organic acids and derivatives | 1.00 (0.83-1.22) | 0.96 (0.77-1.20) | 0.93 (0.71-1.21) | 0.92 (0.70-1.21) | 0.56 | >0.99 | 0.92 (0.66-1.28) | 0.92 (0.66-1.28) | 0.61 | >0.99 |
| pseudouridine | HMDB0000767 | Nucleosides, nucleotides, and analogues | 0.99 (0.79-1.24) | 1.01 (0.78-1.30) | 0.89 (0.64-1.24) | 0.88 (0.62-1.23) | 0.45 | >0.99 | 0.96 (0.64-1.46) | 0.96 (0.63-1.46) | 0.85 | >0.99 |
| pyridoxamine | HMDB0001431 | Pyridines and derivatives | 0.99 (0.80-1.23) | 0.92 (0.72-1.17) | 0.94 (0.70-1.27) | 0.95 (0.70-1.30) | 0.77 | >0.99 | 1.07 (0.74-1.54) | 1.07 (0.74-1.54) | 0.72 | >0.99 |
| quinine | HMDB0014611 | Alkaloids and derivatives | 0.96 (0.51-1.83) | NA | NA | NA | NA | NA | NA | NA | NA | NA |
| ribothymidine | HMDB0000884 | Nucleosides, nucleotides, and analogues | 1.18 (0.94-1.47) | 1.16 (0.90-1.50) | 0.99 (0.71-1.37) | 0.98 (0.70-1.37) | 0.92 | >0.99 | 0.85 (0.57-1.26) | 0.84 (0.56-1.26) | 0.40 | >0.99 |
| serine | HMDB0000187 | Carboxylic acids and derivatives | 1.18 (0.95-1.47) | 1.22 (0.95-1.56) | 1.31 (0.98-1.77) | 1.32 (0.97-1.78) | 0.07 | >0.99 | 1.39 (0.96-2.02) | 1.40 (0.96-2.05) | 0.08 | >0.99 |
| serotonin | HMDB0000259 | NA | 1.08 (0.86-1.37) | 1.00 (0.76-1.31) | 0.96 (0.70-1.32) | 0.93 (0.67-1.29) | 0.66 | >0.99 | 1.12 (0.71-1.75) | 1.11 (0.71-1.75) | 0.64 | >0.99 |
| sphinganine | HMDB0000269 | NA | 0.87 (0.46-1.63) | NA | NA | NA | NA | NA | NA | NA | NA | NA |
| sulfamethoxazole | HMDB0015150 | Benzene and substituted derivatives | NA | NA | NA | NA | NA | NA | NA | NA | NA | NA |
| sulfapyridine | HMDB0015028 | NA | NA | NA | NA | NA | NA | NA | NA | NA | NA | NA |
| thiamine | HMDB0000235 | NA | 1.22 (0.98-1.52) | 1.17 (0.91-1.50) | 1.20 (0.89-1.62) | 1.28 (0.91-1.79) | 0.15 | >0.99 | 1.22 (0.82-1.82) | 1.22 (0.82-1.82) | 0.33 | >0.99 |
| threonine | HMDB0000167 | Organic acids and derivatives | 1.03 (0.83-1.27) | 1.00 (0.78-1.27) | 1.07 (0.79-1.44) | 1.07 (0.79-1.46) | 0.65 | >0.99 | 1.13 (0.78-1.65) | 1.13 (0.78-1.65) | 0.51 | >0.99 |
| thyroxine | HMDB0000248 | Carboxylic acids and derivatives | 0.98 (0.80-1.20) | 0.92 (0.73-1.16) | 0.85 (0.64-1.13) | 0.85 (0.63-1.15) | 0.29 | >0.99 | 0.99 (0.70-1.39) | 0.98 (0.69-1.39) | 0.93 | >0.99 |
| trigonelline | HMDB0000875 | Alkaloids and derivatives | 1.34 (1.08-1.66) | 1.24 (0.97-1.60) | 1.22 (0.91-1.64) | 1.29 (0.93-1.79) | 0.12 | >0.99 | 1.34 (0.91-1.97) | 1.35 (0.91-1.99) | 0.13 | >0.99 |
| trimethylamine-N-oxide | HMDB0000925 | Organonitrogen compounds | 1.01 (0.83-1.24) | 0.96 (0.77-1.21) | 0.85 (0.64-1.12) | 0.82 (0.62-1.10) | 0.18 | >0.99 | 0.95 (0.67-1.34) | 0.95 (0.67-1.34) | 0.78 | >0.99 |

| METABOLITE | HMDB_ID | Metabolite class | Model 1 | Model 2 | Model 3 | Model 4 | | | Model 5 | Model 6 | | |
|---|---|---|---|---|---|---|---|---|---|---|---|---|
| | | | OR (95%CI) | OR (95%CI) | OR (95%CI) | OR (95%CI) | p-value | NEF | OR (95%CI) | OR (95%CI) | p-value | NEF |
| trimethylbenzene | HMDB0013733 | Benzene and substituted derivatives | 0.84 (0.68-1.04) | 0.86 (0.67-1.09) | 0.86 (0.64-1.14) | 0.85 (0.63-1.13) | 0.26 | >0.99 | 0.91 (0.64-1.28) | 0.91 (0.64-1.29) | 0.59 | >0.99 |
| tryptophan | HMDB0000929 | Organoheterocyclic compounds | 0.97 (0.79-1.19) | 0.98 (0.77-1.24) | 0.92 (0.69-1.23) | 0.94 (0.70-1.26) | 0.67 | >0.99 | 0.98 (0.71-1.37) | 0.98 (0.71-1.37) | 0.92 | >0.99 |
| tyrosine | HMDB0000158 | Carboxylic acids and derivatives | 0.87 (0.71-1.05) | 0.80 (0.63-1.01) | 0.76 (0.57-1.01) | 0.76 (0.56-1.03) | 0.08 | >0.99 | 0.83 (0.58-1.20) | 0.83 (0.57-1.19) | 0.31 | >0.99 |
| urate | HMDB0000289 | Imidazopyrimidines | 0.94 (0.77-1.14) | 0.99 (0.78-1.26) | 0.84 (0.62-1.14) | 0.81 (0.59-1.11) | 0.19 | >0.99 | 0.85 (0.58-1.23) | 0.85 (0.58-1.24) | 0.39 | >0.99 |
| valine | HMDB0000883 | Carboxylic acids and derivatives | 0.88 (0.72-1.08) | 0.83 (0.65-1.05) | 0.76 (0.57-1.01) | 0.75 (0.55-1.01) | 0.06 | >0.99 | 0.71 (0.48-1.04) | 0.71 (0.48-1.04) | 0.08 | >0.99 |
| valine-d8 | NA | NA | 1.15 (0.90-1.47) | 1.23 (0.92-1.64) | 1.25 (0.88-1.79) | 1.27 (0.89-1.82) | 0.19 | >0.99 | 1.42 (0.89-2.25) | 1.43 (0.90-2.27) | 0.13 | >0.99 |
| valsartan | HMDB0014323 | NA | 21.52 (0.02-20759.26) | NA | NA | NA | NA | NA | NA | NA | NA | NA |
| verapamil | HMDB0001850 | NA | NA | NA | NA | NA | NA | NA | NA | NA | NA | NA |
| warfarin | HMDB0001935 | NA | NA | NA | NA | NA | NA | NA | NA | NA | NA | NA |
| xanthine | HMDB0000292 | NA | 0.71 (0.38-1.33) | NA | NA | NA | NA | NA | NA | NA | NA | NA |

Abbreviations: NA=not available (as model did not converge); NEF=number of effective tests; OR=odds ratio; XFG=exfoliation glaucoma

**Model 1**: basic model, adjusting for matching factors only (see Table 1); **Model 2** (factors that affect metabolite levels): Model 1 + age, sex, smoking status, BMI, physical activity, fasting status, time of day of blood draw, month of blood draw; **Model 3** (presumed exfoliation syndrome risk factors): Model 2 + family history of glaucoma, type of Caucasian, time out in sunlight in the summer in youth, non-melanoma skin cancer, latitude, population density; **Model 4** (factors that may raise homocysteine levels): Model 3 + folate intake, caffeine intake, alcohol intake, caloric intake; **Model 5** (systemic comorbidities suggested to be associated with XFS in some studies): Model 4 + heart disease, hypertension, high cholesterol, hearing loss, diabetes, stroke, sleep duration; **Model 6** (use of drugs associated with glaucoma): Model 5 + steroid use.
